# Criteria for Detection of Married or Cohabiting Couples in the UK Biobank

**DOI:** 10.1101/2025.07.29.25332337

**Authors:** Kristen M. Kelly, Tanya B. Horwitz, Matthew C. Keller

## Abstract

**Introduction:** The UK Biobank (UKB) dataset contains a large number of spouse/partner pairs (hereafter, “couples”), making it a valuable resource for research on human mating. UKB data does not report relationships between participants, and UKB’s preliminary participant colocation data (based on participants sharing the same street address) is not available for new researchers to request. This has led to different criteria for identification of spouse/partner pairs and highly discrepant sample sizes across papers, potentially contributing to heterogeneity in results. To address this, we developed and validated a standardized method for identifying couples that maximizes sample size while minimizing selection bias.

**Methods:** We evaluated combinations of geographically-derived variables for identifying colocated UKB participants and selected six variables that performed well when compared to with the original UKB colocation data. These variables, such as “distance to coast,” do not reveal participants’ precise locations but are unlikely to match for non-colocated individuals. We then established additional criteria to confirm that colocated participants shared a household and were likely couples. These criteria were designed to be compatible with either geographic colocation or the original UKB colocation data.

**Results:** The geographically-derived variables identified 92,510 putative couples, compared to 89,278 detected using the UKB colocation data. Further analyses suggested that the additional pairs identified in our geographic sample were likely valid pairs not captured by the UKB colocation data. We also assessed certain criteria used in previous studies—such as age, income, and duration of residence—and demonstrated how they could result in biased or non-representative samples.

**Discussion:** Our approach produced a large, robust sample of couples while minimizing false positives. The criteria are flexible and can be applied using either geographic or UKB colocation data. To facilitate further research, we have made the R code for implementing these criteria publicly available at https://github.com/kkellysci/UKBCouples/

## Introduction

Research on romantic, marital, and sexual relationships has broad applications, in part because such relationships are closely linked to a wide array of important life outcomes. These include physical and mental health [1,2], health behaviors [3,4], substance use [5,6], financial security [7,8], stress and social support [9,10], sleep patterns [11], time spent on household work [12], and exposures such as second-hand smoke and infectious disease [13–15]. These and other factors may be influenced by an individual’s partner or more generally by relationship status itself, but these factors may also impact partner choice.

There is widespread evidence for positive correlations between the traits of mates, spouses, and romantic couples (hereafter, “couples”) [16], which can arise through various mechanisms. Assortative mating (AM) may occur directly, as couples choose one another based on similar levels of a specific trait (phenotypic homogamy), or indirectly, through selection based on correlated traits or social factors, such as shared educational, socioeconomic, or professional environments. Convergence, a separate process, occurs when partners grow more similar over time due to mutual environmental influences or reciprocal behavior, such as shared health practices. These mechanisms are not mutually exclusive and may act in combination, influencing traits such as socioeconomic status, health behaviors, and genetic similarity, while also violating the “random mating” assumption in genetic studies and potentially biasing results [17–19].

Several papers have studied couples using the UK Biobank (UKB), a large dataset with extensive information on approximately half a million people ages 37-73 living in the United Kingdom. While relationships between participants are not directly reported in this sample, UKB’s comprehensive data has allowed researchers to identify putative couples in the cohort who are likely to be married or romantically cohabitating. Estimates of the number of couples in this sample have varied widely, ranging from under 19,000 to over 105,000 [20,21].

Studies examining couples in the UKB have typically relied on demographic, geographic, and household-related variables to infer couple status, but the rationale for selecting these variables is often unclear and the criteria differ across studies.

This inconsistency in couple detection criteria limits the comparability of results across studies. Moreover, certain criteria may introduce unintended biases. For example, one study required that individuals’ smoking status match their partner’s response to a household smoking question, but the household smokers question was only asked of non-smokers due to a skip pattern, leading to accidental exclusion of couples who smoked [16,20]. Similarly, criteria that limit age differences between couples [22–24] can artificially inflate similarities in age-related traits, while requiring equal durations of residence at a current address [20,23,25] may disproportionately exclude newly-formed relationships, remarriages, or couples where one partner moved into the other’s home.

Another important source of differences between existing papers lies in their methods of determining participant colocation. Several years ago, the UKB produced a colocation file which used anonymous group IDs to indicate groups of participants who appeared to reside at the same address (without revealing the addresses). The colocation data was provided to some researchers in a preliminary form, but a final version was never produced, and the original preliminary colocation data is no longer available for other researchers to request. Some researchers have also used other geographic variables to attempt to detect colocated couples,[24,29] but these approaches have not been compared to UKB’s colocation data and often require the use of sensitive data such as grid coordinates that risk identifying participant locations.

This paper aims to establish criteria for identifying couples in the UKB sample that (a) can be applied consistently across studies, (b) maximize sample size, and (c) minimize false positives, false negatives, and selection bias. We conduct analyses to identify the most effective variables and criteria for distinguishing true couples from mismatched pairs and to assess the likelihood of false couples in the final sample. We identify a set of geographic variables that can be used for accurate participant colocation, then propose and validate criteria for verifying couple status which is suitable for use with either geographic colocation or the UKB colocation file. We have released R code for implementing these criteria, enabling researchers—including those without access to the UKB grid coordinates or colocation data—to apply them in their studies.

## Methods

### Overview

Our criteria for identifying likely couples consist of 9-10 steps, outlined in Box 1, and are divided into three stages:

1. Identifying colocated groups (Steps 0–2): Determine which individuals belong to groups of colocated participants.
2. Filtering to identify pairs (Steps 3–5): Narrow down individuals and groups to form pairs that could plausibly be couples.
3. Selecting likely couples (Steps 6–9): Apply additional filters to identify pairs most likely to be genuine couples.

An optional Step 10 can be used with UKB-colocated samples to further verify colocation by incorporating geographic variables.

### Colocation steps

We began by identifying groups of colocated individuals, using two different approaches. The first approach uses the UK Biobank preliminary colocation data. This file consists of two columns, participant ID and group, and does not contain any addresses or locations. Each group indicates a set of participants who matched on address. The second approach identifies groups of individuals who match on several geographically-derived variables (described in a later section) and are therefore likely to reside in the same location. We used both approaches to create two samples of colocated groups, then applied our other eligibility steps to both samples to create a “UKB-colocated sample” and a “geographically-colocated sample”.

Steps 1 and 2 then split colocated individuals who reported different household sizes into separate groups. These steps help to address the possibility that multiple households residing at the same address (such as families living in different flats within the same building) could be combined into a single colocation group. This is possible with UKB colocation as well as geographic colocation because the UKB colocation data was created by matching participants on street addresses only (without considering flat numbers). Including household size at this early stage allows these households to be separated and retained in the data rather than kept together in an incorrectly-combined group that would be excluded at a later stage. Incorrectly-combined households not separated by this step are detected and excluded in other steps, particularly Steps 3, 5, 8, and 9.

### Filtering of individuals and groups

Steps 3-5 refine colocated groups to identify pairs of individuals who may be couples. Step 3 excludes groups where members’ self-reported household size is smaller than the group size, as this likely indicates multiple households of the same size that were mistakenly combined during colocation. Step 4 narrows the sample to individuals who report living with a spouse or partner, removing other household members (e.g., children, roommates) from the groups. Finally, Step 5 selects groups of exactly two individuals, excluding groups where multiple couples may be present. While Steps 3 and 5 reduce false positives, they may also increase false negatives by removing households where multiple couples reside at the same address.

### Filtering of pairs

Steps 6-9 further filter pairs to ensure they are likely to be couples. Step 6 restricts the sample to opposite-sex pairs. An expanded version of our criteria that address additional considerations for detecting same-sex couples is presented in Horwitz, 2025 [26]. Step 7 excludes related individuals to avoid misclassifying relatives as couples. For example, when two related UKB participants (e.g., siblings) live together with their non-UKB-participant spouses, step 7 prevents them from being classified as a couple. Relatedness was determined using UKB genetic data, which is based on genetic markers that do not show large differences between ancestry groups [27], thus making it possible to include cross-ancestry pairs in our sample.

During development of Step 7, we observed a small number of pairs (UKB: 106; geographic: 117) with kinship coefficients suggesting third-degree relationships, potentially reflecting cousin marriages, which are common in some immigrant communities [28]. Many such pairs (UKB: 51; geographic: 60) had at least one partner born outside the UK; these pairs also had small age differences (median: 4 years; 95th percentile: 10 years). Based on this, we excluded second-degree relatives (kinship coefficient of 2^−7/2^ [29]) but retained third-degree relatives. Researchers concerned about consanguinity can adjust this threshold to exclude third-degree relatives. In addition, a small number of pairs (UKB: 3975, geographic: 4110) could not be relatedness-checked because a member of the pair either lacked genetic data or failed genetic quality control. We retained these pairs due to the very low rates of relatedness found among pairs that could be checked.

Steps 8 and 9 apply additional checks to exclude mismatched pairs. Step 8 retains only pairs with compatible self-reported household member types and household members consistent with there being only one couple in the household, providing another layer of protection against misclassified relatives. Step 9 retains pairs that match on either (a) Assessment Center and Date of Visit or (b) self-reported number of vehicles, home ownership status (own vs. rent), and gas hob/cooker usage. The justification for these steps and their impact on the sample of putative couples is described below.

Step 10 is an optional step to verify geographic colocation when the UKB colocation file is used in Step 1. This step matches pairs on geographic variables derived from participant locations (see Supplemental Table 1). It is optional because both the UKB colocation file and the geographic variables are derived from UK Biobank’s internal data on participant addresses (for some geographic variables, they are derived with additional intermediary steps, as described in Supplemental Table 1). When the UKB colocation file and the geographic variables do not agree, it is unclear which source of data is incorrect, particularly for variables that were derived from older versions of UKB’s 100m and 1km grid coordinates (which have known issues[30]). For researchers working with large samples, Step 10 is recommended to ensure the highest-quality data. However, researchers focusing on specific subsets of couples such as ethnic minorities or particular age groups may choose to skip Step 10 to maximize sample size.

### Selection of variables for geographic colocation

To select variables for geographic colocation, we began by reviewing location-derived variables and chose those with less than 2% missing data and fewer than 25% of values equal to the mode (most common value). This process identified 15 qualifying variables (detailed in Supplemental Table 1), of which 13 were selected for further analysis. We excluded north/south grid coordinates due to their restricted status and potential to reveal participant locations. Assessment center was also excluded from the geographic colocation criteria, as true couples may differ on this variable. For example, common mismatches such as “Manchester / Bury,” “Manchester / Stockport,” and “Barts / Croydon” involved assessment centers within the same metropolitan areas, likely reflecting true couples who attended different centers for reasons such as workplace location or scheduling convenience.

We tested all 8100 possible combinations of 3 to 13 variables for their effectiveness in identifying colocated participants. Each combination was evaluated against UKB colocation data, 1 km grid coordinate variables, and geographic variables not included in the combination. Further details on the testing process and variable selection are provided in Supplemental Note 1.

### Selection of variables for Steps 8 and 9 to exclude mismatched pairs

To select variables for confirming likely couples, we analyzed pairs remaining after Step 7. Potential match-confirming variables are described in Supplemental Table 3. For each variable, we calculated the match rate among Step 7 pairs, a stringently-filtered subset of Step 7 pairs, and pseudo-couples consisting of individuals drawn from the same assessment center who reported living with a spouse/partner. Further details of this analysis are provided in Supplemental Note 2.

### Estimating the sensitivity of our full criteria

To evaluate the sensitivity of our criteria, we used a set of “ground truth couples” who had a UKB-participating child genetically related to both parents. We began by identifying parent-offspring pairs in the UKB genetic data, defined as pairs having a kinship coefficient consistent with first-degree relatives (2^−5/2^ to 2^−3/2^)[29], an estimated proportion of IBS0 loci <= 0.0012,[27] and an age difference of at least 14 years. We excluded parent-offspring pairs that were ambiguous due to the parent having a monozygotic twin in the data, then merged mother-offspring and father-offspring pairs to create parent-offspring trios. We detected 1061 such trios, containing 1024 unique mother-father pairings (some couples had more than one offspring in the UKB). We then restricted these pairings to those who were colocated according to the UKB colocation file OR who matched on all 15 geographic variables listed in Supplemental Table 1, leaving 973 pairs.

Finally, we required that both members of the pair report living with a spouse/partner, producing a final set of 895 ground truth couples.

We estimated the sensitivity of our criteria as the proportion of ground truth couples correctly identified as couples. We also performed additional analyses to examine which steps of our criteria were responsible for excluding ground truth couples that did not meet our full criteria.

Our sample of ground truth couples relies on the following assumptions: 1) If the two colocation methods disagree on whether two individuals with a genetically-related child are colocated, the true answer is that they are colocated. 2) No couple is incorrectly classified as “not colocated” by both colocation methods. 3) Couples with a child who is a UK Biobank participant have the same probability of failing to meet criteria as other couples in the sample. Assumptions 2 and 3 are very unlikely to be true, so our estimates of sensitivity are likely to be influenced by these assumptions. Of particular note, the target age range for recruitment to UKB (40-69 years) means couples with a child old enough to participate in UKB will tend to be older when compared to our full sample of putative couples.

### Estimating the specificity of our full criteria

To evaluate the probability of incorrectly-matched pairs passing our full eligibility criteria, we used a sample of 489,718 UK Biobank participants with non-missing data the six geographic variables. We generated “pseudo-couples” by randomly pairing individuals within either “assessment center” or “1 km grid square”, excluding one person at random from strata with an odd number of participants. We excluded pairs that matched in the UKB colocation data (to avoid pairing individuals with their original partners), but did not exclude pairs that matched on geographic variables because the geographic variables were part of the criteria tested in this analysis. If the same pseudo-couple was created more than once in different repetitions, duplicate instances were excluded. We repeated this procedure 30 times, creating more than 5 million unique pseudo-couples.

Within each repetition, we applied an abbreviated version of our criteria (Steps 0, 1, 2, 4, 6, 7, 8, 9, and 10). We did not include Steps 3 and 5 because they were not applicable to the pseudo-couples. In this analysis, missing data was handled according to the rules normally used by each step, so pseudo-couples with missing data on a variable necessary for a step would be counted as excluded by that step unless that step had a mechanism to allow for missing data (eg. Steps 9 and 10).

Estimates of specificity from this analysis are subject to the following assumptions: 1) Exclusion of UKB-colocated pairs guarantees that the pseudo-couples contain no true couples. 2) Individuals who are incorrectly identified as colocated by either colocation method are not more likely to pass our criteria than our randomly-drawn pseudo-couples. Assumption 1 is unlikely to be true because our other analyses suggest that some geographically-colocated but not UKB-colocated pairs are true pairs, so it is likely this analysis under-estimates the specificity of geographic colocation. Assumption 2 is unlikely to be true because factors that lead to individuals incorrectly being classified as colocated (such as living in the same apartment building) may also increase their probability of matching on other criteria (such as having/not having a gas stove). This is likely to lead to over-estimation of specificity.

### Creation and comparison of final eligible samples

We applied all eligibility steps (shown in Box 1) to create two samples: one based on the UKB colocation data (“UKB-colocated sample”) and one using our geographic colocation method (“geographically-colocated sample”). We then examined pairs that appeared in one sample but not the other to determine potential reasons for the discrepancy.

For both the UKB-colocated and geographically-colocated couples, we examined partner correlations for the 80 traits listed in Supplemental Table 10. We used tetrachoric correlations for binary traits, Pearson correlations for most continuous traits, and Spearman correlations for continuous traits that were not normally distributed. For normally-distributed continuous traits, we removed outliers who were more than 4 standard deviations above or below the mean for their sex, and for non-normally distributed counts, we eliminated participant responses above the 99.5^th^ percentile for their sex. For non-normal count variables with a small number of possible values (such as number of days per week that a participant did something), we did not apply outlier exclusion.

### Examination of potential bias from certain match-checking variables used in previous studies

After finalizing our eligible samples, we analyzed subsets of couples that would have been excluded by specific match-checking variables employed in previous UKB assortative mating studies. Each analysis focused on the effects of a single match criterion such as household income, length of time at current residence, or within-pair age differences rather than re-implementing the full criteria used by existing papers. To evaluate the impact of various match criteria, we used our final UKB-colocated sample of 89,278 pairs and compared pairs that would meet each additional match criterion to pairs that would have been excluded if we had used that criterion. Although we initially planned to evaluate pairs with an age difference of ≥ 20 years, the sample size (N=143) was too small for analysis. Instead, we examined pairs with an age difference of ≥ 10 years to assess the broader effects of an age-based criterion. Additional details about the sizes of these subsets and the eligible pairs used for comparison are provided in Table 3 and Supplemental Table 12.

For each of the 80 traits shown in Supplemental Table 10, we tested for differences in partner trait correlations and trait means between the included and excluded subgroups. Details of the statistical analysis are available in Supplemental Note 3. All tests used a Bonferroni-corrected alpha of 7.81 * 10^−5^ to account for 640 comparisons (2 tests (mean and correlation) * 80 traits * 4 subgroups). This analysis did not adjust for covariates because the goal was to show all traits for which a given criterion had the potential to influence sample composition. If the relationship between a criterion and trait occurred by way of a commonly-used covariate (such as between-subset differences in blood pressure being related to age), that indirect relationship is still a relevant result.

The number of pairs excluded by a criterion also affects its potential to bias results. For example, excluding thousands of pairs may have a larger impact than excluding a few hundred pairs. To examine this aspect, we created three additional samples consisting of all UKB-colocated pairs meeting our main criteria plus one additional criterion (income, age, or time at address) in addition to our full UKB-colocated and geographically-colocated samples. We used bootstrap sampling to examine differences in partner correlations between the samples while accounting for sample overlap. Details of the bootstrap analysis are available in Supplemental Note 3. All tests used a Bonferroni-corrected alpha of 1.56 * 10^−4^ to account for 320 comparisons (80 traits * 4 samples compared to the UKB-colocated sample).

### Post-hoc examination of pairs discordant for missing data on a trait

While implementing our main analyses, we observed that trait means for individuals excluded due to their partner’s missing data on a trait sometimes appeared to differ from trait means based on pairs where both members had non-missing data. To further examine this pattern, we used a version of our UKB-colocated sample in which outliers were winsorized rather than excluded, so all missing values would reflect data that was missing in the original UKB variables. We selected 53 variables that had at least 1% missing data and conducted a series of t-tests comparing values for individuals whose partner had complete data on a trait to individuals whose partner had missing data. We performed the tests separately for each sex so differences in missingness rates by sex would not lead to sex-related differences in the compared groups. All tests used a Bonferroni-corrected alpha of 4.72 * 10^−4^ to account for 106 comparisons (2 sexes * 53 traits).

## Results

### Variables for geographic colocation

Details for the testing and selection of geographic colocation variables are provided in Supplemental Note 1 and Supplemental Table 2. The performance of variable combinations—evaluated against the UKB colocation file and other geographic variables not included in the combinations—improved notably with each additional variable up to six. Beyond six variables, performance gains were minimal. Based on these findings, we selected the following six-variable combination for use for geographic colocation: Nitrogen dioxide, Nitrogen oxides, Inverse distance to major road, Evening noise, Coast distance, and Townsend Deprivation Index (TDI) [31–34].

Only a single group identified using the geographic colocation variables had non-matching 1 km grid coordinates. However, this group was also listed as colocated in the UKB colocation data, matched on all 7 other geographic variables not used for geographic colocation, and met the full eligibility criteria. This suggests that this group is truly colocated but may have been affected by inaccuracies known to exist in older versions of the grid coordinate data[30]. Additionally, four geographically-colocated groups had mismatches on at least one of the seven geographic variables not included in the final combination. All four were excluded during later criteria steps: three by Step 3 and one by Step 6.

### Selection of match-confirming variables

Details for the testing and selection of potential match-confirming variables are provided in Supplemental Note 2 and Supplemental Table 4. The check for compatible household member types (described in Box 2) produced a high match rate (97.8% of pairs that passed steps 1-7) but a moderate rate of spurious matches (49.7%). This check was incorporated as Step 8, separate from the match-confirming variable combination applied in Step 9. The Step 9 match confirming variables produced a match rate of 98.1% and a spurious match rate of 11.5%. Allowing two ways for couples to pass step 9 (matching on three self-reported variables *or* attending the same assessment center on the same date) also reduced exclusions due to missing data.

### Sensitivity and specificity

Table 1 shows estimated sensitivity using “ground truth pairs” who had a genetically-related offspring in the UKB sample. Estimated sensitivity was slightly higher with geographic colocation (94.0%) than UKB colocation (91.4%). During the initial creation of the ground truth pair sample, 57 otherwise-qualifying pairs were excluded because one person had missing data for whether they had a spouse/partner. 52 of these 57 pairs consisted of one member who reported a household size of 2 and reported a spouse/partner, and one member who reported a household size of 1 and was not asked about household member types due to a skip pattern. When these 57 pairs were included with the ground truth couples, estimated sensitivity was notably lower (geo: 88.3%, ukb: 85.9%), primarily driven by the exclusion of the pairs with non-matching self-reported household sizes in step 2.

**Table 1:** Number of “ground truth” couples passing each criteria step. Table 1 estimates sensitivity by applying our criteria to a sample of ground truth couples. Each row shows the number of ground-truth couples meeting a given step’s requirements as well as the requirements of all preceding steps. The left half of the table uses a sample meeting our full definition of ground truth couples. In the right half of the table, this definition is loosened to include pairs where one member reported a spouse/partner and the other had missing data for household member types.

|  | Ground truth pairs |  | Ground truth pairs, including pairs where one person had missing data on household member types |  |
| --- | --- | --- | --- | --- |
|  | Geographic colocation | UKB colocation | Geographic colocation | UKB colocation |
| Starting number of ground truth couples | 895 | 895 | 952 | 952 |
| Classified as colocated | 886 (0.990) | 861 (0.962) | 942 (0.989) | 915 (0.961) |
| Step 1: No missing data on household size | 883 (0.987) | 858 (0.959) | 935 (0.982) | 908 (0.954) |
| Step 2: Match on self-reported household size | 865 (0.966) | 842 (0.941) | 866 (0.910) | 843 (0.886) |
| Step 3: Colocation group size <= household size | 860 (0.961) | 839 (0.937) | 861 (0.904) | 840 (0.882) |
| Step 4: Both members report living with a spouse or partner | 860 (0.961) | 839 (0.937) | 860 (0.903) | 839 (0.881) |
| Step 5: Only two people in the colocation group report a spouse/partner | 856 (0.956) | 835 (0.933) | 856 (0.899) | 835 (0.877) |
| Step 6: Pair is opposite-sex | 856 (0.956) | 835 (0.933) | 856 (0.899) | 835 (0.877) |
| Step 7: Pair members are not first or second degree genetic relatives | 856 (0.956) | 835 (0.933) | 856 (0.899) | 835 (0.877) |
| Step 8: Compatible self-reported household member types | 850 (0.950) | 829 (0.926) | 850 (0.893) | 829 (0.871) |
| Step 9: Met either match-confirming variable requirement | 841 (0.940) | 820 (0.916) | 841 (0.883) | 820 (0.861) |
| Step 10: Matched on non-missing geographic variables |  | 818 (0.914) |  | 818 (0.859) |

Table 2 presents the results of the pseudo-couple analysis using most steps of our full criteria. Each row shows the proportion of pseudo-couples passing the specified step and all preceding steps. In both analyses, very few pseudo-couples met the full match criteria, resulting in an estimated specificity of >99.9% for both the UKB-colocated and geographically-colocated samples. When pseudo-couples were drawn from within the same 1 km grid square, estimated specificity was lower than when pseudo-couples were drawn from the same assessment center, suggesting that individuals in close geographic proximity are more similar and more likely to produce false matches. Among individuals in close enough proximity to produce false colocation matches, specificity is likely to decline further. The analysis using 1 km grid squares also over-represents pseudo-couples from high-population-density squares, since low-density squares could only produce a small number of non-duplicate pseudo-couples.

**Table 2:** Proportion of randomly-paired “pseudo-couples” passing each criteria step. Table 2 shows the probabilities of randomly-paired “pseudo-couples” drawn from the same assessment center or 1 km grid square passing each stage of our criteria. Each row shows the proportion of pseudo-couples meeting a given criterion’s requirements as well as the requirements of all preceding criteria steps. Specificity can be estimated as “1 – the proportion of pseudo-couples passing criteria”. For steps 1-9, the proportion of pseudo-pairs passing our criteria for the UKB-colocated sample is notably higher than for geographic colocation. This is not a true difference, instead it reflects the fact that these estimates could not account for the unknown (but low) probability of an incorrect match occurring in the UKB colocation file.

|  | Pseudo-couples drawn from the same assessment center<br>(n=7,340,735) |  | Pseudo-couples drawn from the same 1km grid square<br>(n=5,521,581) |  |
| --- | --- | --- | --- | --- |
|  | Criteria for geographically-colocated sample | Criteria for UKB-colocated sample | Criteria for geographically-colocated sample | Criteria for UKB-colocated sample |
| Geographic colocation: Matched on 6/6 geographic variables | 1.8E-06 |  | 0.0009 |  |
| Step 1: No missing data on household size | 1.8E-06 | 0.9827 | 0.0008 | 0.9824 |
| Step 2: Match on self-reported household size | 9.5E-07 | 0.2971 | 0.0006 | 0.3044 |
| Step 4: Both members met requirements | 2.7E-07 | 0.2145 | 0.0004 | 0.2182 |
| Step 6: Pair met step 6 requirements | 2.7E-07 | 0.1066 | 0.0004 | 0.1088 |
| Step 7: Not second-degree or closer relatives | 2.7E-07 | 0.1066 | 0.0004 | 0.1088 |
| Step 8: Compatible household member types | 1.4E-07 | 0.1043 | 0.0004 | 0.1064 |
| Step 9: Met final match-confirming variable requirements | 1.4E-07 | 0.0149 | 0.0003 | 0.0238 |
| Matched on assessment center and assessment date | 1.4E-07 | 0.0004 | 0.0003 | 0.0077 |
| Matched on other match-confirming variables | **1.4E-07** | 0.0146 | **0.0003** | 0.0177 |
| Step 10: Matched on non-missing geographic variables |  | **1.8E-06** |  | **0.0009** |

**Table 3:** Differences in trait correlations and means for subsets of pairs that would be excluded by other match-checking criteria. Table 3 shows results when subsets of pairs in our final sample that would be excluded by an additional match-checking criterion are compared to the rest of the sample. Additional details of this analysis are provided in Supplemental Note 3 and Supplemental Tables 14-17.

|  | Traits where pairs in the subset have higher correlation | Traits where pairs in the subset have lower correlation | Number of traits with significant mean differences |
| --- | --- | --- | --- |
| Missing data on income (n=20777) | Birthplace (east coordinate), Birthplace (north coordinate), Whole body impedance, Number of children, Number of siblings, Walking (minutes/day) | Age, Educational attainment, Moderate physical activity (days/week), Time spent watching TV, Drinks full-fat or semi-skimmed milk | 59 |
| Non-matching income (n=15295) |  | Age, Educational attainment, Moderate physical activity (days/week), Vegetable intake, Time spent watching TV, Walking (minutes/day), Drinks full-fat or semi-skimmed milk, Eats processed meat >= 1x week | 11 |
| Time at current residence differs by ≥ 5 years (n=6249) |  | Age, Age at first intercourse, Educational attainment, Forced expiratory volume in 1 second, Number of children, Drinks alcohol, Smoked at least 100 times, Smoking status (Former), Smoking status (Never) | 34 |
| Age difference ≥ 10 years (n=3897) |  | Age, Age at first intercourse, Birthplace (east coordinate), Birthplace (north coordinate), Educational attainment, Forced expiratory volume in 1 second, Forced vital capacity, Hand grip strength, Height, Peak expiratory flow, Systolic blood pressure, Number of children, Number of illnesses, Number of medications, Number of sexual partners, Smoking status (Former), Smoking status (Never), Wears Glasses | 31 |

### Comparisons of the final UKB-colocated and geographically-colocated samples

We analyzed the 1,308 pairs that were unique to the UKB-colocated sample and the 4,540 pairs that were unique to the geographically-colocated sample. The UKB-colocated pairs not in the geographic sample were missing data on geographic variables (n=1,072) or had their households combined with others during geographic colocation (n=236), with 199 of the latter pairs reporting a housing type of “flat, maisonette, or apartment”. Most pairs unique to the geographic sample were not colocated according to the UKB colocation file (n=4,523); the remaining pairs, despite being grouped together in the UKB colocation file, were excluded from the final UKB-colocated sample because they were grouped with additional individuals. As shown in Supplemental Tables 9a-9c, pairs that were not in the UKB colocation file were more likely to have earlier intake years and to have attended certain assessment centers, suggesting that issues such as address formatting or regional factors may have contributed to discrepancies. These pairs were also more likely to report the housing type “flat, maisonette, or apartment,” suggesting that some may be incorrect matches between individuals living in close proximity.

Trait correlation results for the UKB-colocated and geographically-colocated samples (Figure 1, Supplemental Table 11) are consistent with previous studies, including Horwitz et al. (2023).[21] Correlations produced by our two colocation methods were nearly identical, which is unsurprising given the 95–99% overlap between the samples. In our bootstrap analysis (Figure 2, Supplemental Table 18), only 1 of 80 traits (number of children) showed a significant difference in correlation when comparing the geographic and UKB-colocated samples.

**Figure 1:**
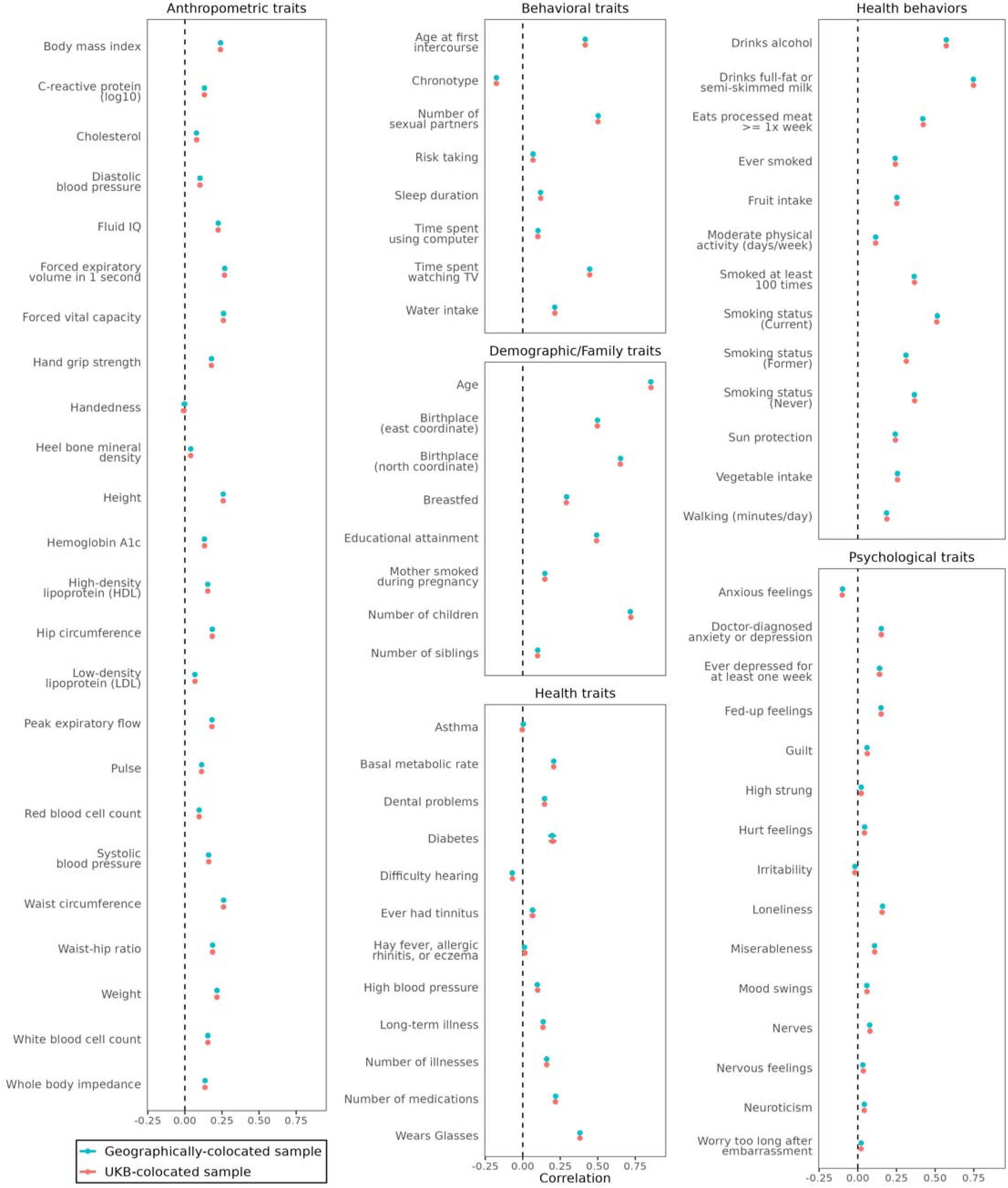
Trait correlations in geographically-colocated and UKB-colocated samples. Figure 1 shows partner correlations and 95% confidence intervals for 80 traits in both our UKB-colocated (red) and geographically-colocated (turquoise) samples.

**Figure 2:**
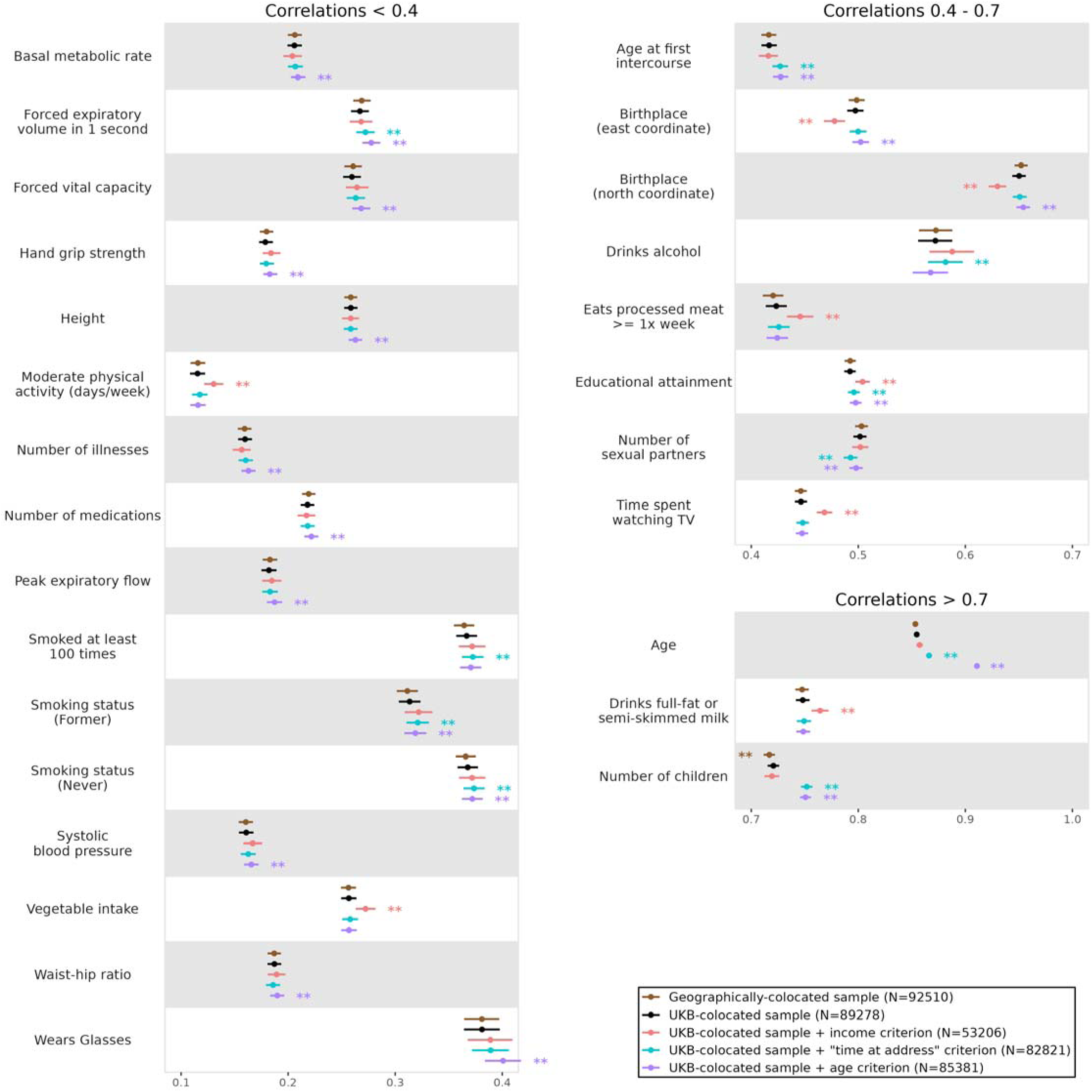
Differences in trait correlations when samples using additional match-checking criteria are compared to the main samples. Figure 2 shows differences in trait correlations for our full UKB-colocated sample, our full geographically-colocated sample, and three samples created by adding an additional match criterion (income, time at address, or age difference) to the UKB-colocated sample.

### Examination of couples that would be excluded by other commonly-used match variables

Table 3 and Supplemental Tables 14-17 show results comparing subsets of pairs that would be excluded by match-confirming variables used in previous studies. Results of the bootstrap analysis comparing our full eligible samples with these variables added to the criteria are shown in Figure 2 and Supplemental Table 18.

Excluding pairs based on income-related criteria showed high potential to bias analysis results. Pairs with missing data differed from the rest of the sample on 11 correlations and 59 trait means, while pairs with non-matching income differed on 8 correlations and 11 means. Applying an income-based criterion to our sample in the bootstrap analysis produced significant changes in correlation for 8 traits. Changes in correlation were often larger for income than for other criteria, in part due to the large number of pairs that were excluded. Many of these traits (such as educational attainment, health behaviors, and birth coordinates) have a known association with socioeconomic factors.

Pairs with an age difference ≥ 10 years and pairs with a difference of ≥5 years in time at their current address showed notably lower correlations for number of children in the subset analysis (0.37 and 0.29, respectively) than the comparison pairs (0.75) potentially indicating a higher proportion of re-marriages in these groups. Age-differing pairs also showed lower correlations for several age-associated traits such as blood pressure and hand grip strength. In the bootstrap analysis (Figure 2), these criteria often produced smaller changes in estimated correlations due to the smaller number of pairs excluded. However, excluding pairs for age differences still produced significant changes in correlation for 20 traits, while excluding pairs for differences in time at current residence changed correlations for 10 traits.

### Post-hoc examination of pairs discordant for missing data on a trait

Supplemental Figure 1 and Supplemental Tables 19-20 compare trait means between individuals whose partner has missing data on a trait and those whose partner has non-missing data. Significant differences were observed for 25 of 53 variables in at least one sex, with all 13 variables that were significant in both sexes showing consistent directions in differences. The impact of these differences on analyses depends on factors such as the type of analysis, the number of pairs excluded for missing data, the specific variable and reasons for missingness, and whether excluded pairs systematically differ from others (e.g., lower similarity on the trait or older age). Although a full investigation of missing data issues is beyond the scope of this paper, we present these findings to help researchers using UK Biobank couple data recognize and account for these patterns in their analyses.

## Discussion

In this paper, we present two related developments: a method for identifying UKB participants likely to reside in the same household without inferring their actual geographic locations, and a set of criteria for verifying putative couples in the UKB sample. These criteria are compatible with both the UKB colocation data (for researchers with access) and our geographic colocation method. Our criteria are designed to minimize bias and improve representativeness, avoiding variables strongly associated with factors like socioeconomic status or remarriage that could distort results in future studies. We evaluated the strengths and limitations of our approach and analyzed our final samples (which were larger than those used in most previous UKB studies) to other match variables and criteria from past research. To facilitate further research, we have released R code to implement our criteria (https://github.com/kkellysci/UKBCouples/), making couple-based analyses more accessible to researchers using UKB data.

Our geographic variable combination enables participant colocation detection without using sensitive data that could risk identifying participant locations. Unlike restricted variables such as grid coordinates, which directly indicate location, our method relies on derived measures, such as model-based air pollution estimates and Euclidean distance to the coastline. These variables, even when combined, are unlikely to pose a risk of identification of participants’ physical locations due to the complexity of their derivation and the numerous locations within the UK that could produce matching values.

However, when two individuals match on all six variables, it is highly likely they reside in the same location (although their location remains unknown), and the subsequent steps in our criteria confirm the match while filtering out spurious matches. This method performed well when compared to 1 km grid coordinate data and provides a privacy-preserving approach for detecting potential couples without requiring sensitive location variables.

Our geographically-colocated sample includes 4,523 couples (4.9%) who are not colocated according to the UKB colocation file. Several lines of evidence suggest that many of these additional couples are true couples who were missing from the UKB colocation data. First, our analysis of geographic variable combinations found that 80.7% of pairs that were geographically-colocated but not UKB-colocated attended the same assessment center on the same date, a notably higher proportion than the 0.4% observed when individuals attending the same assessment center were paired at random. Second, among our 895 ground truth pairs with genetically-related offspring, 34 (3.8%) pairs are not in the UKB colocation file despite matching on geographic variables. Finally, the UKB colocation file is considered preliminary data and was not released in a finalized form, so it is likely that this data contains some errors.

Excluding pairs with missing or discordant self-reported incomes had a notable impact on analysis results for several traits associated with socioeconomic factors. The proportion of income mismatches in our 895 ground truth pairs (21.7%) was similar to that in our full UKB-colocated sample (22.3%), suggesting that these mismatches are common in true pairs and not the result of incorrect pairing. The discordant pairs may reflect participants inaccurately estimating household income, or in some cases reporting individual rather than household income (a possibility supported by extreme mismatches such as “<£18,000” vs. “>£100,000”). An income-related criterion would also harm statistical power by excluding up to 36,072 pairs, and performs poorly at detecting incorrectly-matched pairs (Supplemental Table 4), so we strongly recommend against the use of income as a match-confirming variable.

Excluding pairs based on age differences or time at current residence resulted in smaller changes in partner correlations due to the smaller number of pairs excluded by these criteria. However, these exclusions risk removing true couples, particularly those with non-traditional characteristics such as remarriages, later-life relationships, or larger age differences. While restricting age differences might help differentiate couples from cohabiting relatives in datasets without other pertinent data, it is unnecessary in the UK Biobank where pairs can be checked using genetic relatedness and self-reported household member types.

Reported sample sizes have varied widely in previous studies of UKB couples. The smallest estimate for the number of couples, 18,984, was produced by a paper which based colocation on 1 km grid coordinates, likely leading to exclusion of many households that were incorrectly combined during colocation.[20] Studies producing samples of 40,000-50,000 couples also used a smaller number of variables for geographic colocation,[19,25] and sometimes used strict criteria (such as requiring that couples report exactly the same number of years at their current residence) likely to exclude many true couples. Two other papers produced sample sizes of 79,074 and 90,927,[16,24] more consistent our samples, while the largest known estimate is 105,381.[21]

Our results show that approximately 36% of UKB participants have a UKB-participating spouse or partner. Participation of a friend or family member has previously been reported to influence individuals’ research participation decisions, [35,36] and UK Biobank’s low participation rate will contribute to over-representation of individuals with characteristics likely to increase participation.[37] Although the large number of couples is beneficial for couples research, it also raises the issue of non-independence between observations. Other results have also identified potential non-independence in the UKB sample, such as the finding that 10.7% of UKB participants have a genetic first-degree relative in the sample (usually a sibling) and 30.3% have a third-degree or closer relative.[27] Researchers analyzing variables for which couples, household members, or genetic relatives might be more similar to each-other should carefully consider how this might influence their analysis.

Our study has several limitations. First, our analyses of sensitivity and specificity have several assumptions and limitations, described in detail in the methods section. Second, our criteria may exclude true couples due to missing data or inaccurate responses, and may exclude some couples in multi-family housing (eg. flats/apartments) due to their households being incorrectly merged with neighbors. (Approximately 4.9% of all UKB participants who report living with a spouse or partner have this housing type.) Third, couples with live-at-home adult children or “temporary” household members may also be excluded, as criteria like household size, member types, and number of vehicles are prone to mismatches when individuals disagree on who counts as part of their household. Additionally, the UK Biobank sample is non-representative in several ways[37] and subject to participation bias that may distort results[38]. While our criteria aim to minimize addition of bias, they do not address biases already present in the dataset. Individuals who participated in UK Biobank with their spouse/partner may also differ from UKB participants with a non-participating spouse/partner. Fifth, our criteria currently identify only opposite-sex couples. A modified version designed to detect same-sex couples has been developed, and further details will be described in Horwitz et al. (2025)[26]. Finally, although the risk of false matches is low, our criteria may still incorrectly identify a small number of false couples.

Our colocation method and couple identification criteria have several strengths. First, our selected combination of geographic variables does not rely on participant location coordinates or the UKB colocation data. This makes the criteria accessible to any researcher with standard UKB data while minimizing the risk of identifying participant locations.

Second, our pseudo-pair analysis showed that incorrectly colocated pairs are likely to be excluded by later criteria, improving accuracy with either colocation method. Third, our criteria are designed to reduce bias and enhance representativeness by avoiding variables linked to factors like socioeconomic status or remarriage, which could distort studies on assortative mating or couples more generally. We also offered flexibility in Step 9 by allowing pairs to meet the match-confirming check in two ways: either by matching on all confirming variables or by attending the same assessment center on the same date. This reduces the impact of incorrect or missing data for any single match-confirming variable.

Finally, our criteria produce large samples, providing high statistical power and enabling adequate sample sizes for studying couples in underrepresented or minority groups.

#### Box 1: Criteria for detecting and verifying potential couples

**Step 0:** Begin by identifying groups of colocated participants, using either the UKB colocation data or colocation via geographic variables.

**Step 1:** Exclude people who are missing data on “number of people in household”

**Step 2**: Split colocated groups that disagree on self-reported household size into separate groups that agree on household size.

**Step 3:** Exclude groups where the colocation group size exceeds its members’ self-reported household sizes.

**Step 4**: Keep only people who report living with a spouse/partner

**Step 5**: Keep only groups with exactly two individuals who passed step 4.

**Step 6:** Keep only opposite-sex pairs.

**Step 7:** Exclude pairs that are 1^st^ or 2^nd^ degree relatives

**Step 8:** Keep only pairs with compatible self-reported household member types (see Box 2)

**Step 9:** Keep only pairs matching on either:

- Assessment Center AND Date of Assessment Center visit OR
- Number of vehicles AND own vs. rent (broad)* AND gas hob/cooker

**Step 10 (optional)**: If using the UKB colocation file, exclude pairs with mismatches on any of the geographic colocation variables. Missing values are allowed as matches.

* Broad matching for the own vs. rent variable allowed missing values and uncommon responses such as “part mortgage, part rent” to count as matches, as shown in Supplemental Table 5a.

#### Box 2: Determining compatibility of self-reported household member types

**Criteria for compatible household member types:**

1. If one person reports living with their sibling, parent, or grandparent, the other person must report living with an unrelated individual or “other relative“
2. If one person reports a household member type of “Other relative” or “Unrelated”, the other person must report “Other relative”, “Unrelated”, “Sibling”, “Parent”, “Grandparent”, or “Grandchild” “Grandchild” is only allowed as an explanation if the other person does not also report grandchildren, to prevent unclear combinations like “Spouse/Partner, Child, Grandchild” with “Spouse/Partner, Child, Grandchild, Unrelated“
3. Exclude households where there could be multiple couples living together, since it would be difficult to determine whether the two individuals in the data are part of the same couple

a. If both people report household member types “other relative”, “unrelated” and household size > 3
b. If either person reports household member types “parent” or “grandparent” and household size > 3 (because the household might include two parents or two grandparents who are a couple)
4. If one person reports living with only their spouse/partner and nobody else, the other person must too
5. If one person reports living with relative type “Child or Stepchild”, the other person must too
6. If one person reports living with relative type “Grandchild”, the other person must report “Grandchild”, “Other relative”, or “Unrelated“

After accounting for differences in order, there were 17 compatible household member combinations found in the data, shown in Supplemental Table 7.

## Data Availability

UK Biobank data is available to researchers who complete the UK Biobank application process.
Code implementing our method is publicly available at https://github.com/kkellysci/UKBCouples/

## Acknowledgements

This research has been conducted using the UK Biobank Resource under Application Number 16651. This work used the Blanca condo computing resource at the University of Colorado Boulder. Blanca is jointly funded by computing users and the University of Colorado Boulder. Data storage supported by the University of Colorado Boulder ‘PetaLibrary’.

## Notes

### Competing Interest Statement

The authors have declared no competing interest.

### Funding Statement

This study was funded by the National Institute of Mental Health R01 grants MH130448 (KMK, TBH, MCK) and MH100141 (KMK,
MCK) and by the National Institute on Drug Abuse T32 Training Grant DA017637 (TBH).

### Author Declarations

This analysis used de-identified data from an existing dataset (UK Biobank) and was therefore exempt from human subjects review by the Institutional Review Board of the University of Colorado, Boulder. The UK Biobank has ethical approval from the North West Multi-centre Research Ethics Committee. Our use of geographic variables for participant colocation has been reviewed by UK Biobank's Scientific Team, who have given us approval to publish our method and criteria.

### Summary of Updates

Corrected to use our current title for the manuscript. Version 1 accidentally used the old title.

## References

[1] D. Meyler, J.P. Stimpson, M.K. Peek, Health concordance within couples: A systematic review, Soc. Sci. Med. 64 (2007) 2297–2310. 10.1016/j.socscimed.2007.02.007.

[2] B.J. Ayotte, F.M. Yang, R.N. Jones, Physical Health and Depression: A Dyadic Study of Chronic Health Conditions and Depressive Symptomatology in Older Adult Couples, J. Gerontol. Ser. B 65B (2010) 438–448. 10.1093/geronb/gbq033.

[3] D. Shiffman, J.Z. Louie, J.J. Devlin, C.M. Rowland, S. Mora, Concordance of Cardiovascular Risk Factors and Behaviors in a Multiethnic US Nationwide Cohort of Married Couples and Domestic Partners, JAMA Netw. Open 3 (2020) e2022119. 10.1001/jamanetworkopen.2020.22119.

[4] S.E. Jackson, A. Steptoe, J. Wardle, The Influence of Partner’s Behavior on Health Behavior Change: The English Longitudinal Study of Ageing, JAMA Intern. Med. 175 (2015) 385–392. 10.1001/jamainternmed.2014.7554.

[5] J.L. Treur, J.M. Vink, D.I. Boomsma, C.M. Middeldorp, Spousal resemblance for smoking: Underlying mechanisms and effects of cohort and age, Drug Alcohol Depend. 153 (2015) 221–228. 10.1016/j.drugalcdep.2015.05.018.

[6] C. Reczek, T. Pudrovska, D. Carr, M.B. Thomeer, D. Umberson, Marital Histories and Heavy Alcohol Use among Older Adults, J. Health Soc. Behav. 57 (2016) 77–96. 10.1177/0022146515628028.

[7] J.M. Zissimopoulos, B.R. Karney, A.J. Rauer, Marriage and economic well being at older ages, Rev. Econ. Househ. 13 (2015) 1–35. 10.1007/s11150-013-9205-x.

[8] S. Britt-Lutter, C. Dorius, D. Lawson, The Financial Implications of Cohabitation Among Young Adults, J. Financ. Plan. 31 (2018) 38–45.

[9] L.A. Neff, B.R. Karney, Stress Crossover in Newlywed Marriage: A Longitudinal and Dyadic Perspective, J. Marriage Fam. 69 (2007) 594–607. 10.1111/j.1741-3737.2007.00394.x.

[10] L.K. Soulsby, K.M. Bennett, Marriage and Psychological Wellbeing: The Role of Social Support, Psychology 6 (2015) 1349–1359. 10.4236/psych.2015.611132.

[11] R.C. Richmond, L.J. Howe, K. Heilbron, S. Jones, J. Liu, X. Wang, M.N. Weedon, M.K. Rutter, D.A. Lawlor, G. Davey Smith, C. Vetter, Correlations in sleeping patterns and circadian preference between spouses, Commun. Biol. 6 (2023) 1–14. 10.1038/s42003-023-05521-7.

[12] T. Lichard, F. Pertold, S. Škoda, Do women face a glass ceiling at home? The division of household labor among dual-earner couples, Rev. Econ. Househ. 19 (2021) 1209–1243. 10.1007/s11150-021-09558-7.

[13] K. Shah, D. Saxena, D. Mavalankar, Secondary attack rate of COVID-19 in household contacts: a systematic review, QJM Int. J. Med. 113 (2020) 841–850. 10.1093/qjmed/hcaa232.

[14] Y. Kobayashi, K. Yamagishi, I. Muraki, Y. Kokubo, I. Saito, H. Yatsuya, H. Iso, S. Tsugane, N. Sawada, Secondhand smoke and the risk of incident cardiovascular disease among never-smoking women, Prev. Med. 162 (2022) 107145. 10.1016/j.ypmed.2022.107145.

[15] B.Y. Hernandez, L.R. Wilkens, X. Zhu, P. Thompson, K. McDuffie, Y.B. Shvetsov, L.E. Kamemoto, J. Killeen, L. Ning, M.T. Goodman, Transmission of Human Papillomavirus in Heterosexual Couples, Emerg. Infect. Dis. 14 (2008) 888–894. 10.3201/eid1406.070616.2.

[16] T.B. Horwitz, J.V. Balbona, K.N. Paulich, M.C. Keller, Evidence of correlations between human partners based on systematic reviews and meta-analyses of 22 traits and UK Biobank analysis of 133 traits, Nat. Hum. Behav. 7 (2023) 1568–1583. 10.1038/s41562-023-01672-z.

[17] C.C. Minică, D.I. Boomsma, C.V. Dolan, E. de Geus, M.C. Neale, Empirical comparisons of multiple Mendelian randomization approaches in the presence of assortative mating, Int. J. Epidemiol. 49 (2020) 1185–1193. 10.1093/ije/dyaa013.

[18] R. Border, S. O’Rourke, T. de Candia, M.E. Goddard, P.M. Visscher, L. Yengo, M. Jones, M.C. Keller, Assortative mating biases marker-based heritability estimators, Nat. Commun. 13 (2022) 660. 10.1038/s41467-022-28294-9.

[19] R. Border, G. Athanasiadis, A. Buil, A.J. Schork, N. Cai, A.I. Young, T. Werge, J. Flint, K.S. Kendler, S. Sankararaman, A.W. Dahl, N.A. Zaitlen, Cross-trait assortative mating is widespread and inflates genetic correlation estimates, Science 378 (2022) 754–761. 10.1126/science.abo2059.

[20] L. Yengo, M.R. Robinson, M.C. Keller, K.E. Kemper, Y. Yang, M. Trzaskowski, J. Gratten, P. Turley, D. Cesarini, D.J. Benjamin, N.R. Wray, M.E. Goddard, J. Yang, P.M. Visscher, Imprint of assortative mating on the human genome, Nat. Hum. Behav. 2 (2018) 948–954. 10.1038/s41562-018-0476-3.

[21] A. Tenesa, K. Rawlik, P. Navarro, O. Canela-Xandri, Genetic determination of height-mediated mate choice, Genome Biol. 16 (2016) 269. 10.1186/s13059-015-0833-8.

[22] T.B. Horwitz, J.V. Balbona, K.N. Paulich, M.C. Keller, Evidence of correlations between human partners based on systematic reviews and meta-analyses of 22 traits and UK Biobank analysis of 133 traits, Nat. Hum. Behav. 7 (2023) 1568–1583. 10.1038/s41562-023-01672-z.

[23] M.R. Robinson, A. Kleinman, M. Graff, A.A.E. Vinkhuyzen, D. Couper, M.B. Miller, W.J. Peyrot, A. Abdellaoui, B.P. Zietsch, I.M. Nolte, J.V. van Vliet-Ostaptchouk, H. Snieder, S.E. Medland, N.G. Martin, P.K.E. Magnusson, W.G. Iacono, M. McGue, K.E. North, J. Yang, P.M. Visscher, Genetic evidence of assortative mating in humans, Nat. Hum. Behav. 1 (2017) 1–13. 10.1038/s41562-016-0016.

[24] K. Rawlik, O. Canela-Xandri, A. Tenesa, Indirect assortative mating for human disease and longevity, Heredity 123 (2019) 106–116. 10.1038/s41437-019-0185-3.

[25] L.J. Howe, D.J. Lawson, N.M. Davies, B. St. Pourcain, S.J. Lewis, G. Davey Smith, G. Hemani, Genetic evidence for assortative mating on alcohol consumption in the UK Biobank, Nat. Commun. 10 (2019) 5039. 10.1038/s41467-019-12424-x.

[26] T.B. Horwitz, K.M. Kelly, M.C. Keller, A Comparison of Partner Resemblance Trends Across Putative Same-Sex and Opposite-Sex Couples in the UK Biobank, Nat. Sci. Rep. (In press).

[27] C. Bycroft, C. Freeman, D. Petkova, G. Band, L.T. Elliott, K. Sharp, A. Motyer, D. Vukcevic, O. Delaneau, J. O’Connell, A. Cortes, S. Welsh, A. Young, M. Effingham, G. McVean, S. Leslie, N. Allen, P. Donnelly, J. Marchini, The UK Biobank resource with deep phenotyping and genomic data, Nature 562 (2018) 203–209. 10.1038/s41586-018-0579-z.

[28] A. Shaw, Drivers of Cousin Marriage among British Pakistanis, Hum. Hered. 77 (2014) 26–36. 10.1159/000358011.

[29] A. Manichaikul, J.C. Mychaleckyj, S.S. Rich, K. Daly, M. Sale, W.-M. Chen, Robust relationship inference in genome-wide association studies, Bioinformatics 26 (2010) 2867–2873. 10.1093/bioinformatics/btq559.

[30] UK Biobank, Deriving the Grid Coordinates (Version 2.1), (2024). https://biobank.ndph.ox.ac.uk/ukb/ukb/docs/UKgrid.pdf (accessed June 23, 2023).

[31] R. Beelen, G. Hoek, D. Vienneau, M. Eeftens, K. Dimakopoulou, X. Pedeli, M.-Y. Tsai, N. Künzli, T. Schikowski, A. Marcon, K.T. Eriksen, O. Raaschou-Nielsen, E. Stephanou, E. Patelarou, T. Lanki, T. Yli-Tuomi, C. Declercq, G. Falq, M. Stempfelet, M. Birk, J. Cyrys, S. von Klot, G. Nádor, M.J. Varró, A. Dėdelė, R. Gražulevičienė, A. Mölter, S. Lindley, C. Madsen, G. Cesaroni, A. Ranzi, C. Badaloni, B. Hoffmann, M. Nonnemacher, U. Krämer, T. Kuhlbusch, M. Cirach, A. de Nazelle, M. Nieuwenhuijsen, T. Bellander, M. Korek, D. Olsson, M. Strömgren, E. Dons, M. Jerrett, P. Fischer, M. Wang, B. Brunekreef, K. de Hoogh, Development of NO2 and NOx land use regression models for estimating air pollution exposure in 36 study areas in Europe – The ESCAPE project, Atmos. Environ. 72 (2013) 10–23. 10.1016/j.atmosenv.2013.02.037.

[32] D.W. Morley, K. de Hoogh, D. Fecht, F. Fabbri, M. Bell, P.S. Goodman, P. Elliott, S. Hodgson, A.L. Hansell, J. Gulliver, International scale implementation of the CNOSSOS-EU road traffic noise prediction model for epidemiological studies, Environ. Pollut. 206 (2015) 332–341. 10.1016/j.envpol.2015.07.031.

[33] UK Small Area Health Statistics Unit, Resource 2010: Environmental Exposures Metadata, (2014). https://biobank.ndph.ox.ac.uk/showcase/refer.cgi?id=2010 (accessed June 29, 2024).

[34] B. Wheeler, Resource 15374: Environmental indicators attributed to participant’s home location, (2016). https://biobank.ndph.ox.ac.uk/showcase/refer.cgi?id=15374 (accessed June 29, 2024).

[35] V.A. Diaz, A.G. Mainous, A.A. McCall, M.E. Geesey, Factors affecting research participation in African American college students, Fam. Med. 40 (2008) 46–51.

[36] K. Haas, D. Costley, M. Falkmer, A. Richdale, K. Sofronoff, T. Falkmer, Factors Influencing the Research Participation of Adults with Autism Spectrum Disorders, J. Autism Dev. Disord. 46 (2016) 1793–1805. 10.1007/s10803-016-2708-6.

[37] A. Fry, T.J. Littlejohns, C. Sudlow, N. Doherty, L. Adamska, T. Sprosen, R. Collins, N.E. Allen, Comparison of Sociodemographic and Health-Related Characteristics of UK Biobank Participants With Those of the General Population, Am. J. Epidemiol. 186 (2017) 1026–1034. 10.1093/aje/kwx246.

[38] T. Schoeler, D. Speed, E. Porcu, N. Pirastu, J.-B. Pingault, Z. Kutalik, Participation bias in the UK Biobank distorts genetic associations and downstream analyses, Nat. Hum. Behav. 7 (2023) 1216–1227. 10.1038/s41562-023-01579-9.

