## Supplemental material for "Criteria for Detection of Married or Cohabiting Couples in the UK Biobank" (from another version of this preprint: version 1, 2025-07-30)

**Supplemental Note 1: Selection of variables for geographic colocation** 2

**Supplemental Note 2: Selection of match-confirming variables** 3

**Supplemental Note 3: Methods for statistical analyses** 5

**Supplemental Figure 1: Trait means stratified by partner's missing data status for that trait** 6

**Supplemental Table 1: Potential variables for geographic colocation** 7

**Supplemental Table 2: Evaluation of variable combinations for geographic colocation** 8

**Supplemental table 3: Potential match-confirming variables** 9

**Supplemental Table 4: Proportion of matches on potential match-confirming variables between members of pairs that passed steps 1-7 and between members of random pseudo-couples** 10

**Supplemental Table 5a: Matches and mismatches among UKB-colocated "likely" pairs for the Own vs. rent variable** 11

**Supplemental Table 5b: Matches and mismatches among UKB-colocated "likely" pairs for the Housing Type variable** 11

**Supplemental table 6: Additional variables used in the couple detection criteria** 12

**Supplemental table 7: The 17 compatible household member type combinations detected in the data** 13

**Supplemental table 8: Results of applying each criteria step in the UKB-colocated and geographically-colocated samples** 14

**Supplemental Table 9a: Proportion of individuals in geographic pairs that are not UKB colocated, by assessment year** 16

**Supplemental Table 9b: Proportion of individuals in geographic pairs that are not UKB-colocated, by assessment center** 16

**Supplemental Table 9c: Proportion of individuals in geographic pairs that are not UKB-colocated, by housing type** 16

**Supplemental table 10: Phenotypes and traits** 17

**Supplemental Table 11: Trait correlations and data-derived confidence intervals for the UKB-colocated and geographically-colocated samples** 20

**Supplemental table 12: Subsets and comparison groups for examining the potential impacts of other eligibility variables used in previous studies** 23

**Supplemental table 13a: Matches and mismatches on self-reported household income in the final eligible UKB-colocated sample** 24

**Supplemental table 13b: Matches and mismatches on self-reported household income in a sample with additional eligibility filtering** 24

**Supplemental table 14: Comparison between pairs with missing data on income and pairs with complete income data** 25

**Supplemental table 15: Comparison between pairs that do not match on self-reported household income and those that do** 28

**Supplemental table 16: Comparison between pairs that differ by 5+ years for length of time at current address and those that do not** 31

**Supplemental table 17: Comparison between pairs with an age difference of >= 10 years to pairs with an age difference of < 10 years** 34

**Supplemental Table 18: Trait correlations and bootstrap confidence intervals for all samples, with p-values for comparisons to the full UKB-colocated sample** 37

**Supplemental Table 19: Trait missing value counts for the UKB-colocated pairs** 40

**Supplemental Table 20: Comparison of trait means for people whose partners are or are not missing data for that trait** 43

**Supplemental Note 1: Selection of variables for geographic colocation**

To select variables for geographic colocation, we began by examining variables derived from participant location and selected variables that had less than 2% missing data and fewer than 25% of values equal to the mode (most common value). We identified 15 qualifying variables (shown in Supplemental Table 1), and of these we selected 13 (excluding 1 km grid coordinates) for further analysis. We created all 8100 possible 3-13 variable combinations and tested the effectiveness of each variable combination for participant colocation by comparing results to the UKB colocation data.

To test the variable combinations, we began with a sample of 362,428 UK Biobank participants who reported living with a spouse/partner and had non-missing data for self-reported household size. First, we created a set of UKB-colocated groups within this sample using the UKB colocation file; this set was used for comparisons with the geographically-colocated groups. Next, for each combination of geographic variables, we created geographically-colocated groups that matched on all variables in the combination. We applied household size splitting (described in criteria step 2) to both the UKB-colocated and geographically-colocated groups. For each of our 8100 combinations of geographic variables, we compared geographically-colocated groups to UKB-colocated groups and to other measures likely to be informative for colocation accuracy.

We evaluated our geographic variable combinations based on several measures, including total number of geographically-colocated groups, consistency with the UKB colocation file, and consistency with 1 km grid coordinate data. Additionally, for groups that were geographically-colocated but not UKB-colocated, we examined what proportion of groups had members who attended the same assessment center on the same date (a variable combination with a high proportion of matches in UKB-colocated couples and a low proportion of spurious matches, see "Selection of Match-Confirming Variables") to assess the possibility that geographic colocation may have detected true groups that were missing from the UKB colocation file. For each variable combination, we also examined the proportion of the resulting groups matched or did not match for the other eligible geographic variables not used in that combination.

We found that an 8-variable combination of geographic variables produced groups that did not mismatch on any of the remaining geographic variables, so we restricted our analysis to combinations consisting of 4-8 variables. For each number of variables, we selected the combination that produced the highest proportion of geographically-colocated groups that were also UKB-colocated, and the combination that produced the lowest proportion of groups that failed to match on other eligible geographic variables not in the combination. For combinations involving 6-8 variables, all combinations that produced the best results for one measure also produced the best results for the other measure. The selected combinations are shown Supplemental Table 2.

From these options, we selected the six-variable combination "Nitrogen dioxide, Nitrogen oxides, Inverse distance to major road, Evening noise, Coast distance, TDI" for our analysis, because the additional gains from adding a 7$^{th}$ or 8$^{th}$ variable were minimal. All four groups that match on these variables but mismatch on other geographic variables are successfully excluded by later steps of our eligibility criteria.

**Supplemental Note 2: Selection of match-confirming variables**

*Methods*

To select variables for confirming likely couples, we analyzed pairs remaining after Step 7. Potential match-confirming variables are described in Supplemental Table 3. For each variable, we calculated the match rate among all Step 7 pairs and a subset of "partially-filtered pairs." Partially-filtered pairs were defined as pairs meeting all criteria through Step 7, attending the same assessment center on the same date, and having compatible household member types (as described in Box 2). This filtering was not used in our final criteria, it was used only to assess the potential for false negatives (mismatches among pairs likely to be true couples) when evaluating other variables not used in the filtering.

To evaluate the risk of false positives (matches among false couples), we created a set of pseudo-couples, consisting of randomly paired individuals drawn from the same assessment center. We began with a sample of 355,522 UKB participants who reported living with a spouse or partner, forming random pairs within each assessment center (excluding one individual at random from centers with an odd number of participants). These pseudo-couples were filtered to meet Step 6 eligibility criteria, such as being opposite-sex pairs who matched on household size. Additionally, pseudo-couples who were colocated in the UKB colocation data or matched on our geographic colocation variables were excluded to prevent random pairing with original spouses or partners. This process was repeated 30 times, generating over one million pseudo-couples for analysis.

Pseudo-couples were used to calculate the frequency of spurious matches (random pairs sharing a matching value) for each potential couple-confirming variable. Following individual evaluations, we tested combinations of variables, such as "number of vehicles AND presence of a gas hob/cooker." Pseudo-couples with missing data were excluded from the calculation of spurious match percents for the affected variable.

*Results*

Results for potential match-confirming variables and their combinations are presented in Supplemental Table 4. Variables were evaluated based on their match rates among partially-filtered pairs and their effectiveness in excluding falsely-matched pseudo-couples. The variable "Cooking/Heating types in home" allowed participants to select multiple responses, resulting in a low proportion of pairs with identical combinations. However, agreement was high for whether both members of a pair reported having a gas hob/cooker, making this a viable match-confirming variable. Similarly, "Number of Vehicles" and "Own vs. Rent (broad)" showed high match rates among partially-filtered pairs and relatively low match rates among pseudo-couples. These three variables demonstrated strong potential for accurately distinguishing true couples from non-couples.

A high proportion of UKB-colocated pairs (87.8%) attended the same assessment center on the same date, while spurious matches among pseudo-couples were very rare (0.4%), even though pseudo-couples were always drawn from the same assessment center. Matching on assessment date may indicate individuals who know each other and coordinated their appointments to travel together. To further examine the assumption that matching dates indicated pairs attending together, we created 248,866 pseudo-pairs consisting of non-colocated individuals who attended the same assessment center on the same date, and compared the difference in times recorded for reception at the assessment center (field 21811). Time differences were notably lower for our step 7 pairs who attended the same assessment center on the same date (median: 45 seconds, $95^{th}$ percentile: 4.2 minutes) than for the random pairs (median 2.9 hours, $95^{th}$ percentile 7.7 hours). Although assessment center and date are useful evidence to support a connection between members of a pair, it is still possible for couples to attend different centers or have different assessment dates. We did not make this variable mandatory to avoid excluding true couples, but its high match rate in colocated pairs and its low match rate among pseudo-couples suggested it could be a useful alternative for confirming matches when other variables are missing or inaccurate.

We analyzed combinations of the variables Number of Vehicles, Gas Hob/Cooker, and Own vs. Rent (broad) in the preliminary Step 7 sample. The step 7 sample is likely to contain a high proportion of true pairs, and the "partially-filtered" (see methods) subset of this sample should contain a higher proportion of true pairs. Requiring matches on all three variables yielded the lowest rate of spurious matches among pseudo-couples (11.1%) but matched only 87.9% of all UKB-colocated pairs and 88.6% of partially-filtered UKB-colocated pairs, suggesting a high false negative rate. Less stringent combinations improved match rates among partially-filtered pairs but substantially increased spurious matches. To address this, we tested an alternative criterion allowing pairs to qualify by matching on either "assessment center AND assessment date” or “Number of Vehicles, Own vs. Rent (broad), AND Gas Hob/Cooker.” This approach achieved a high match rate (98.1%) among UKB-colocated pairs while only modestly increasing spurious matches (to 11.5%), as well as reducing the number of pairs excluded for missing data by 91%.

**Supplemental Note 3: Methods for statistical analyses**

*Comparison of correlations and means in included vs. excluded subsets*

To compare correlations, we calculated partner correlations within each subset of interest and then transformed each correlation using a Fisher r-to-z transformation[1]. For each comparison, we then divided the difference between the two transformed correlations by the sum of their estimated variances (the formula for variance of a difference when there is no covariance between the correlations, which applies here because there is no sample overlap) and tested the null hypothesis that this value was equal to zero. For Fisher-transformed Pearson and Spearman correlations, we calculated variance as 1/(N-3) [2] and for Fisher-transformed tetrachoric correlations we approximated variance using the delta method[3]. We used t-tests to compare trait means (for continuous traits) or prevalences (for dichotomous traits). Under the Central Limit Theorem, our sample sizes are large enough to allow for the use of t-tests with non-normally-distributed variables[4]. For these analyses, t-tests were preferable to non-parametric tests due to the potential for differences in variance by subgroup.

*Bootstrap sample comparison*

To perform bootstrapping, we combined the samples into a single dataset with variables indicating a couples' membership in each sample, then performed 250,000 bootstrap sampling iterations. For each iteration, we recorded partner correlation for each trait in each sample, as well as differences between the Fisher-transformed correlations for our full UKB-colocated sample (the reference group) and each of the other samples. For each trait in each sample, we took the minimum of "the proportion of iterations in which the difference from the reference group value was above/below zero," and multiplied this value by 2 to produce the p-value for a two-tailed test of whether the difference between the samples was not equal to 0.


1. Fisher RA. Frequency distribution of the values of the correlation coefficient in samples from an indefinitely large population. *Biometrika*. 1915;10(4):507-521.

2. Myers L, Sirois MJ. Spearman Correlation Coefficients, Differences between. In: *Encyclopedia of Statistical Sciences*. John Wiley & Sons, Ltd; 2006. doi:10.1002/0471667196.ess5050.pub2

3. Efron B. Nonparametric standard errors and confidence intervals. *Can J Stat*. 1981;9(2):139-158. doi:10.2307/3314608

4. Lumley T, Diehr P, Emerson S, Chen L. The Importance of the Normality Assumption in Large Public Health Data Sets. *Annu Rev Public Health*. 2002;23(Volume 23, 2002):151-169. doi:10.1146/annurev.publhealth.23.100901.140546

### Supplemental Figure 1: Trait means stratified by partner's missing data status for that trait

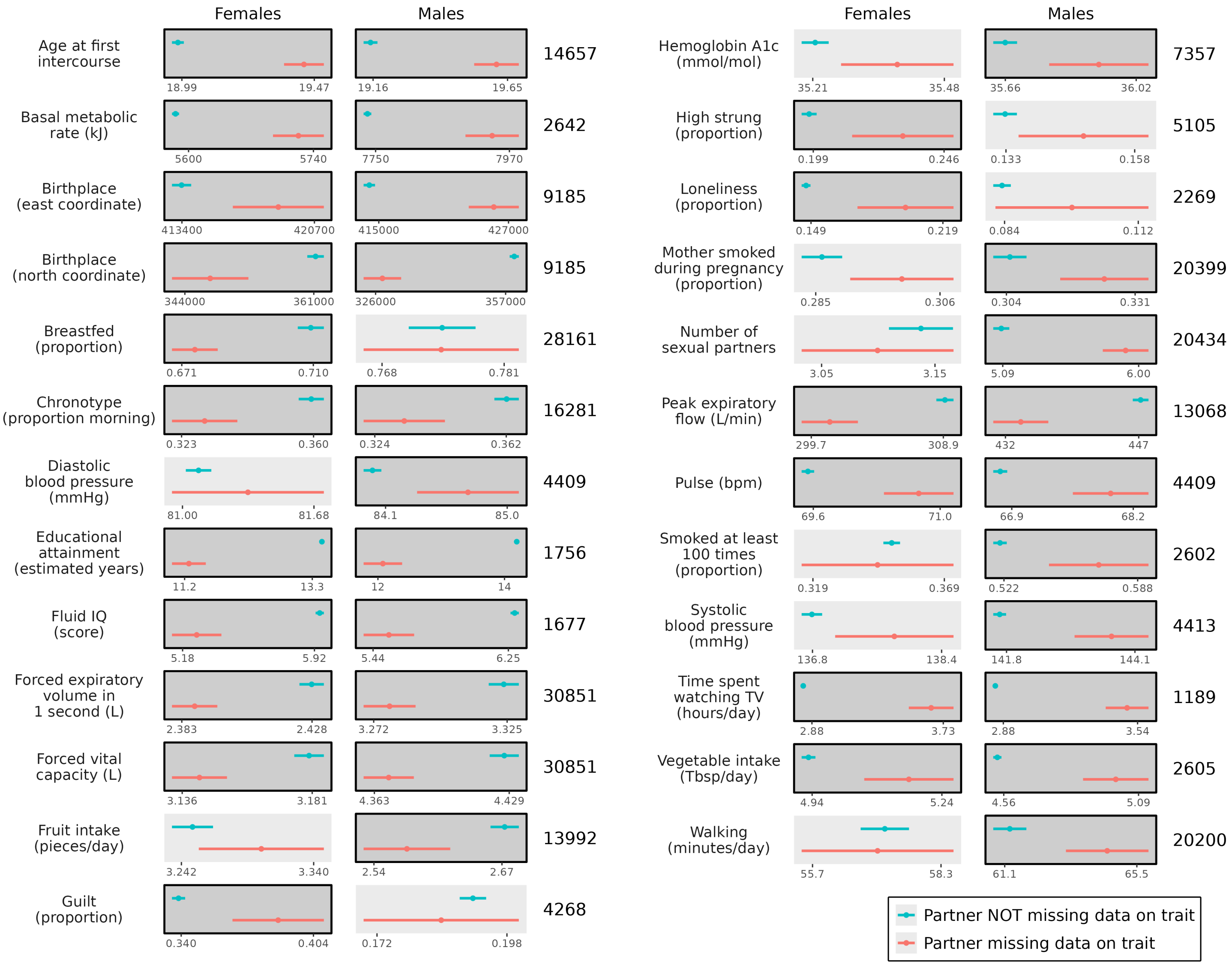


Supplemental Figure 1 compares trait means for people whose partner is missing data on that trait to means for people whose partner is not missing data. Plots show means with 95% confidence intervals. Differences that were significant at the Bonferroni-corrected alpha threshold of $4.72 * 10^{-4}$ are indicated using darker grey rectangles with black borders. The total number of pairs in which exactly one person (male or female) is missing data is shown to the right of each row.

**Supplemental Table 1: Potential variables for geographic colocation**

| Variable | UKB field | Details | Data source |
|---|---|---|---|
| East Grid Coordinate (1 km)*<br><br>North Grid Coordinate (1 km)* | 20074<br><br>20075 | This older (now discontinued) version of the grid coordinate variables was derived from participant postcode rather than full address. Since it is possible for one postcode to overlap with multiple 1 km grid squares, actual resolution for theses variables may be less precise than 1 km. UK Biobank now has updated 1 km grid coordinates (fields 22688 and 22689) based on full addresses, but these variables are classified as restricted data and only available to researchers who complete a special request process. | Participant postcodes overlapped with 1 km grid squares |
| Euclidean distance to coast<br><br>Natural environment percentage, buffer 300m | 24508<br><br>24507 | These variables were created by researchers at the University of Exeter Medical School under UK Biobank project ID 15374 and was provided to UK Biobank as a returned result.[1] They were created based on UK Biobank's 100m grid coordinates (fields 20033 and 20034). This version of the grid coordinates was derived from participant postcode only, not full address.<br><br>1. Wheeler B. Resource 15374: Environmental indicators attributed to participant's home location. https://biobank.ndph.ox.ac.uk/showcase/refer.cgi?id=15374 | 100m grid coordinates (derived from postcodes) |
| Townsend Deprivation Index (TDI) | 189 or 22189 | UK Biobank created this variable by matching the participant's postcode to the output area in the preceding national census. Field 189 contained a more precise version of TDI but was withdrawn and replaced with Field 22189 (which is rounded to 2 digits) to protect participant privacy. At the time of our analysis we had not yet received field 22189, so we rounded field 189 to two digits to emulate the use of field 22189. | Postcodes / census output area |
| Inverse distance to nearest major road<br>Inverse distance to nearest road<br>Nitrogen dioxide air pollution; 2010<br>Nitrogen oxides air pollution; 2010<br>Traffic intensity on the nearest major road<br>Average daytime sound level of noise pollution<br>Average evening sound level of noise pollution<br>Average night-time sound level of noise pollution<br>Average 16-hour sound level of noise pollution<br>Average 24-hour sound level of noise pollution | 24012<br>24010<br>24004<br>24003<br>24011<br>24020<br>24021<br>24022<br>24023<br>24024 | These variables were created by the UK Small Area Health Statistics Unit (SAHSU) in collaboration with multiple research teams.[1] Air pollution estimates for the year 2010 are based on Land Use Regression (LUR) models from the ESCAPE project,[2] noise pollution estimates are based on the CNOSSOS-EU model.[3] These variables were created based on geocoding of participants' full addresses and may offer greater precision for colocation than variables derived from postcode or grid coordinates.<br><br>1. UK Small Area Health Statistics Unit. Resource 2010: Environmental Exposures Metadata. https://biobank.ndph.ox.ac.uk/showcase/refer.cgi?id=2010<br>2. Beelen R, Hoek G, Vienneau D, et al. Development of NO2 and NOx land use regression models for estimating air pollution exposure in 36 study areas in Europe – The ESCAPE project. *Atmos Environ*. 2013;72:10-23. doi:10.1016/j.atmosenv.2013.02.037<br>3. Morley DW, de Hoogh K, Fecht D, et al. International scale implementation of the CNOSSOS-EU road traffic noise prediction model for epidemiological studies. *Environ Pollut*. 2015;206:332-341. doi:10.1016/j.envpol.2015.07.031 | Geo-coded participant addresses |

* Variable excluded from consideration prior to creation of Supplemental Table 2.

Supplemental Table 1 shows variables considered for use with our geographic colocation method. North and east grid coordinates were excluded from consideration so they could be used in evaluating the effectiveness of combinations of other variables. Grid coordinates were also not included in the final geographic colocation criteria because they are restricted variables that are more difficult for researchers to access.

**Supplemental Table 2: Evaluation of variable combinations for geographic colocation**

| Variable combination | Number of variables | Participants excluded for missing data | Results for geographically-colocated groups | | | |
|---|---|---|---|---|---|---|
| | | | Total | Mismatched on other geographic variables | Colocated in the UKB colocation file | Groups not in the UKB colocation file that matched on assessment date |
| Nitrogen oxides, 24-hour noise, Coast distance, TDI | 4 | 6906 | 97412 | 43 (0.00044) | 92244 (0.94695) | 4145 (0.80205) |
| Nitrogen dioxide, Inverse distance to major road, 24-hour noise, Coast distance | 4 | 6592 | 97599 | 135 (0.00138) | 92446 (0.9472) | 4145 (0.80439) |
| Nitrogen oxides, Inverse distance to major road, Evening noise, Coast distance, TDI | 5 | 6906 | 97415 | 11 (0.00011) | 92276 (0.94725) | 4142 (0.80599) |
| Nitrogen dioxide, Nitrogen oxides, Inverse distance to major road, Evening noise, Coast distance | 5 | 6592 | 97601 | 128 (0.00131) | 92454 (0.94726) | 4146 (0.80552) |
| **Nitrogen dioxide, Nitrogen oxides, Inverse distance to major road, Evening noise, Coast distance, TDI** | **6** | **6906** | **97413** | **4 (4e-05)** | **92281 (0.94732)** | **4141 (0.8069)** |
| Nitrogen dioxide, Nitrogen oxides, Inverse distance to nearest road, Inverse distance to major road, Evening noise, Coast distance, TDI | 7 | 6906 | 97413 | 2 (2e-05) | 92283 (0.94734) | 4141 (0.80721) |
| Nitrogen dioxide, Nitrogen oxides, Inverse distance to major road, Evening noise, Nighttime noise, Coast distance, TDI | 7 | 6906 | 97413 | 2 (2e-05) | 92283 (0.94734) | 4141 (0.80721) |
| Nitrogen oxides, Inverse distance to nearest road, Inverse distance to major road, Evening noise, Nighttime noise, Coast distance, TDI | 7 | 6906 | 97413 | 2 (2e-05) | 92283 (0.94734) | 4142 (0.80741) |
| Nitrogen dioxide, Nitrogen oxides, Inverse distance to nearest road, Inverse distance to major road, Evening noise, Nighttime noise, Coast distance, TDI | 8 | 6906 | 97413 | 0 (0) | 92285 (0.94736) | 4141 (0.80753) |

Supplemental Table 2 compares geographically-colocated groups (based on different sets of geographic variables for colocation) to a comparison set of n=94082 UKB-colocated groups. All groups consist of two or more individuals who report living with a spouse or partner. Results in this table show groups *after* household size splitting (described in step 2 of our criteria), which was applied to both the geographically-colocated groups and the UKB-colocated comparison set.

A group as classified as "Mismatched on other geographic variables" if its members have differing (and non-missing) values on at least one of the eligible geographic variables that wasn't part of the combination. A group is classified as "Colocated the UKB colocation file" if *all* members of the group are also part of a colocated group in the UKB colocation file, groups in which only *some* members are UKB-colocated are classified as not UKB colocated.

For geographic groups that were not confirmed by UKB colocation data, we examined the proportion that matched on date of assessment center attendance, a variable that had a very low rate of spurious matches (0.4%) in our analysis of match-confirming variables. The high proportion of matches among non-UKB-colocated groups suggests that some of these groups might be true groups which are missing from the UKB colocation file.

The total number of geographically-colocated groups remains stable at 97413 for combinations of six or more variables, however this does not indicate that the same groups are present in each case. For example, when the 6-variable combination is compared to the 8-variable combination, one duo created by the 6-variable combination is excluded, two trios are reduced to duos, and one group of four is split into two groups of two.

Variable combinations shown in the table were the top-performing N-variable combinations producing either the highest proportion of groups also colocated in the UKB colocation file, or the lowest proportion of groups that had mismatches on other geographic variables. For combinations using six or more variables, all combinations that produced the best results on one metric also produced the best results on the other metric. Our final selected set of geographic colocation variables is shown in bold.

### Supplemental table 3: Potential match-confirming variables

| Potential match criterion | UKB Field ID | Notes |
|---|---|---|
| Length of time at current address | 699 | Match criteria was "differs by <= 5 years". This was not selected as one of the final match criteria due to a relatively low percentage of matches in "likely" pairs and because couples where one person moved into the other's existing residence could legitimately differ on this value. |
| Housing type (flat, bungalow, etc.) | 670 | This variable had 5 possible values: "House or Bungalow", "Flat, Maisonette, or Apartment", "Mobile or Temporary Structure (i.e. caravan)", "None of the Above", and missing values. Two additional possible responses, "Sheltered Accommodation" and "Care Home" were not present in our data. Any individuals giving these responses were not present in our eligible preliminary sample because they were not asked the household size question.<br><br>For the "Exact" match criteria, two individuals had to give the same response.<br><br>For the "Broad" match criteria, we excluded as mismatches any pairs where one person reported "House or Bungalow" and the other reported "Flat, Maisonette, or Apartment", but we allowed the smaller categories (caravan, none of the above, missing) to match to any other value. These were treated as matches on the grounds that differences were more likely to result from differences in how the couple reported their housing (i.e., one person reporting the caravan the couple spends much of their time in while the other reports their permanent residence) than from actual differences in where the two individuals lived. |
| Own vs. rent home | 680 | This variable had 8 possible values: "Own outright (by you or someone in your household", "Own with mortgage", "Rent – from local authority, local council, housing association", "Rent – from private landlord or letting agency", "Pay part rent and part mortgage (shared ownership)", "Live in accommodation rent free", "None of the above", or missing values.<br><br>For the "Exact" match criterion, two individuals had to give the same response.<br><br>For the "Broad" match criterion, the four smaller categories ("Pay part rent and part mortgage (shared ownership)", "Live in accommodation rent free", "None of the above", or missing values) were allowed to match to any other value, on the grounds that mismatches were more likely to result from differences in how two individuals reported their home ownership status than from an actual difference. |
| Income | 738 | This variable had six possible values: the 5 bins originally used by UK Biobank ("Less than £18,000", "£18,000 to 30,999", "£31,000 to 51,999", "£52,000 to 100,000", and "Greater than £100,000"), plus missing values.<br><br>For the "Exact" match criterion, two individuals had to give the same response.<br><br>For the "Broad" match criterion, individuals reporting adjacent bins (e.g., "18,000 to 30,999" and "31,000 to 51,999") were treated as matches. |
| Went to assessment center on same date | 53 | Date an individual attended the assessment center for their baseline/intake visit. |
| Number of vehicles | 728 | Possible values were "None", "One", "Two", "Three", "Four or more", or missing values. An exact match was required, and missing values were excluded. |
| Age difference | 21003 | Our preliminary match criterion required that a pair's ages when they attended the assessment center differ by < 20 years. This was not selected as one of the final match criteria because it is possible for genuine couples to differ by 20 or more years in age, and the exclusion of such couples could bias estimates of assortative mating. In addition, the criteria excluding genetically related individuals and requiring compatible household member types are likely to prevent misclassification of cohabiting relative pairs (such as parent/offspring) that are unlikely to be married or romantic/sexual partners as spouse pairs. Thus, checking age difference is not necessary for this purpose. |
| Cooking methods in home | 6139<br>10860 (pilot) | Individuals could give multiple responses for each cooking/heating type present. Possible responses in the main assessment interview were: "A gas hob or gas cooker", "A gas fire that you use regularly in winter time", "An open solid fuel fire that you use regularly in winter time", "None of the above", and missing values. Possible responses in the pilot assessment interview were: "A gas hob", "A gas cooker", "An open solid fuel fire that you use regularly in winter time", and "None of the above". Although match percentages were poor when looking at whether potential couples agreed for all values each person reported in response to this question, agreement was high on whether a gas hob or cooker was present in the home, so only the category "gas hob or cooker" was used in the final match criteria. |

**Supplemental Table 4: Proportion of matches on potential match-confirming variables between members of pairs that passed steps 1-7 and between members of random pseudo-couples**

| | Geographically-colocated pairs | | | | | UKB-colocated pairs | | | Pseudo-couples (N=2660551) |
|---|---|---|---|---|---|---|---|---|---|
| | Missing | All (N=96470) | Partially-filtered (N=82799) | UKB colocated (N=91681) | Not UKB colocated (N=4789) | Missing | All (N=93471) | Partially-filtered (N=80181) | |
| Years at current address differs by <= 5 | 238 | 0.938 | 0.267 | 0.94 | 0.903 | 229 | 0.939 | 0.939 | 0.267 |
| Housing type (flat, bungalow, etc.) (exact) | 40 | 0.995 | 0.919 | 0.995 | 0.987 | 40 | 0.995 | 0.995 | 0.919 |
| Housing type (flat, bungalow, etc.) (broad) | 0 | 0.997 | 0.924 | 0.997 | 0.992 | 0 | 0.997 | 0.998 | 0.924 |
| Own vs. rent home (exact) | 712 | 0.96 | 0.462 | 0.961 | 0.944 | 699 | 0.96 | 0.964 | 0.462 |
| **Own vs. rent home (broad)** | **0** | **0.966** | **0.486** | **0.966** | **0.957** | **0** | **0.966** | **0.969** | **0.486** |
| Income (exact) | 22493 | 0.772 | 0.241 | 0.773 | 0.754 | 21780 | 0.773 | 0.786 | 0.24 |
| Income (broad)[a] | 22493 | 0.985 | 0.64 | 0.986 | 0.976 | 21780 | 0.986 | 0.987 | 0.641 |
| **Went to the same assessment center on the same date** | **0** | **0.878** | **0.004** | **0.88** | **0.848** | **0** | **0.878** | **--** | **0.004** |
| **Number of vehicles** | **461** | **0.943** | **0.371** | **0.943** | **0.936** | **444** | **0.943** | **0.947** | **0.371** |
| Age difference < 20 years | 0 | 0.998 | 0.907 | 0.998 | 0.997 | 0 | 0.998 | 0.998 | 0.907 |
| Cooking methods in home | 531 | 0.777 | 0.265 | 0.776 | 0.779 | 506 | 0.776 | 0.782 | 0.264 |
| **Gas hob/cooker** | **207** | **0.964** | **0.602** | **0.964** | **0.954** | **205** | **0.964** | **0.965** | **0.602** |
| Gas fire | 578 | 0.845 | 0.534 | 0.845 | 0.851 | 548 | 0.845 | 0.848 | 0.534 |
| Solid fuel fire | 207 | 0.954 | 0.827 | 0.953 | 0.955 | 205 | 0.953 | 0.957 | 0.827 |
| None of the above | 207 | 0.941 | 0.741 | 0.941 | 0.943 | 205 | 0.941 | 0.942 | 0.741 |
| **Compatible household member types** | **0** | **0.978** | **0.497** | **0.978** | **0.977** | **0** | **0.978** | **--** | **0.497** |
| Own vs. rent (broad) AND Gas hob/cooker | 200 | 0.931 | 0.292 | 0.932 | 0.914 | 198 | 0.931 | 0.935 | 0.292 |
| Vehicles AND Gas hob/cooker | 592 | 0.909 | 0.223 | 0.91 | 0.894 | 575 | 0.909 | 0.914 | 0.223 |
| Vehicles AND Own vs. rent (broad) | 450 | 0.911 | 0.185 | 0.912 | 0.899 | 433 | 0.912 | 0.918 | 0.185 |
| Vehicles AND Own vs. rent (broad) AND Gas hob/cooker | 576 | 0.879 | 0.111 | 0.88 | 0.861 | 559 | 0.879 | 0.886 | 0.111 |
| At least two of: Vehicles, Own vs. rent (broad), Gas hob/cooker | 92 | 0.993 | 0.479 | 0.993 | 0.985 | 90 | 0.993 | 0.994 | 0.479 |
| **(Assess cent/date) OR (Vehicles AND Own vs. rent (broad) AND Gas hob/cooker)** | **51** | **0.981** | **0.115** | **0.982** | **0.971** | **50** | **0.981** | **--** | **0.115** |

Supplemental Table 4 shows variables evaluated for use as match-confirming variables. Results are shown for the preliminary samples (pairs meeting criteria through Step 7) for both the geographically-colocated and UKB-colocated samples. Results are shown for all pairs in the preliminary sample, and for "partially-filtered pairs" (pairs that have compatible household member types and attended the same assessment center on the same date). For the geographically-colocated pairs, results are also shown stratified by "UKB confirmed" status (whether or not a pair is also colocated according to the UKB colocation data). The final column shows the percent of spurious matches found in randomly-created "pseudo-couples" consisting of random individuals drawn from the same assessment center.

[a] The criterion "Income (broad)" allows reported income categories to differ by one level (see Supplemental Table 13a), details and definitions for the other "broad" variables are shown in supplemental tables 5a-5b.

#### Supplemental Table 5a: Matches and mismatches among UKB-colocated "likely" pairs for the Own vs. rent variable

| | Own outright | Own with mortgage | Rent (public) | Rent (private) | Part mortgage part rent | Live rent free | None of the above | Missing data |
|---|---|---|---|---|---|---|---|---|
| **Own outright** | 48484 | 2360 | 3 | 22 | 9 | 47 | 93 | 172 |
| **Own with mortgage** | | 25581 | 6 | 20 | 38 | 33 | 80 | 70 |
| **Rent (public)** | | | 1582 | 65 | 4 | 11 | 4 | 1 |
| **Rent (private)** | | | | 730 | 2 | 9 | 20 | 0 |
| **Part mortgage part rent** | | | | | 64 | 1 | 3 | 0 |
| **Live rent free** | | | | | | 223 | 40 | 4 |
| **None of the above** | | | | | | | 45 | 1 |
| **Missing data** | | | | | | | | 354 |

"Rent (public)" indicates "Rent[s] - from local authority, local council, housing association"

"Part mortgage part rent" indicates "Pay[s] part rent and part mortgage (shared ownership)"

#### Supplemental Table 5b: Matches and mismatches among UKB-colocated "likely" pairs for the Housing Type variable

| | House or bungalow | Flat, maisonette, or apartment | Mobile or temporary structure (i.e. caravan) | None of the above | Missing data |
|---|---|---|---|---|---|
| **House or bungalow** | 77371 | 187 | 17 | 166 | 14 |
| **Flat, maisonette, or apartment** | | 2312 | 1 | 11 | 2 |
| **Mobile or temporary structure (i.e. caravan)** | | | 57 | 11 | 0 |
| **None of the above** | | | | 19 | 0 |
| **Missing data** | | | | | 13 |

Supplemental tables 5a and 5b shows the number of responses among UKB-colocated "likely" pairs (pairs that met eligibility criteria 1-7, reported compatible relative types, and attended the assessment center on the same date) for each combination of possible responses to the variables "own vs. rent home" and "housing type". Responses are shown independently of partner sex/order, so all response combinations appear only on the diagonal or in the upper-right triangle of the table.

In Supplemental Table 5a, the rate of mismatches is high for individuals giving less common responses such as "Part mortgage part rent", and "None of the above", while in Supplemental Table 5b the rate of mismatches is high for the responses "Mobile or temporary structure (i.e. caravan)" or "None of the above". Such mismatches are more likely to be the result of differences in how members of a couple interpreted and answered the questions than the result of non-couple status, so we allowed these mismatches to pass under the "broad" versions of match-checking for these variables. Response combinations considered to be exact matches are shown in green, while additional combinations allowed under broad matching are shown in yellow.

**Supplemental table 6: Additional variables used in the couple detection criteria**

| Variable description | Relevant criterion/criteria | UKB Field ID | Notes |
|---|---|---|---|
| Household size | 2 | 709 | Self-reported number of people in a participant's household. Other household members are often not UK Biobank participants, so this number is frequently larger than colocation group size.<br><br>The full text of the question was: "Including yourself, how many people are living together in your household? (Include those who usually live in the house such as students living away from home during term, partners in the armed forces or professions such as pilots)". This question was not asked of individuals who reported living in sheltered accommodation or a care home. |
| Colocation group size | 3 | Not applicable | This value reflects the number of other UK Biobank participants a person is colocated with, according to either the UK Biobank colocation file or colocation via geographic variables. |
| Self-reported sex | 6 | 31 | Because UK Biobank does not have questions for sex assigned at birth and gender identity, we did ***not*** exclude individuals whose self-reported sex (field 31) differed from their inferred sex based on their sex chromosomes (field 22001). As discussed in Ackley and Duchowney 2023,[1] such mismatches are more likely to result from intersex and transgender participants than from sample mislabeling. |
| Genetic kinship with other UK Biobank participants | 7 | Field 22021<br><br>UK Biobank Relatedness File | Field 22021 had five potential values: -1 (Participant excluded from kinship inference process), 0 (No kinship found), 1 (At least one relative found), 10 (Ten or more third-degree relatives identified), and NA (missing data). Individuals with a value of -1 were not checked for relatedness (often because their genetic data failed quality control), and individuals with a value of NA were not checked for relatedness because they did not have genetic data available.<br><br>The UK Biobank relatedness file can be downloaded by approved UK Biobank researchers with the command "gfetch rel", as described in UK Biobank Resource 668. This file contains one potential relative pair per line, with columns for each individual's participant ID (IID_1, IID_2), the percent of markers at which both individuals are heterozygous (HetHet), the percent of markers at which two individuals have no alleles in common (IBS0), and their kinship coefficient (Kinship). Pairs with a kinship coefficient of less than 0.0442 are not in the relatedness file. Pairs that are not in the relatedness file can be assumed to be unrelated ***unless*** one or both individuals has a value of -1 or missing data in field 22021, in which case relatedness between the two individuals was not checked. |
| Household member types | 4, 6, 8 | 6141 | This variable included 8 categories of household member type: "Husband, wife, or partner", "Son and/or daughter (include step-children)", "Brother and/or sister", "Mother and/or father", "Grandparent", "Grandchild", "Other related", and "Other unrelated". Participants could report the presence of each household member type but not the number of people of each type (eg. whether they lived with one grandchild or multiple grandchildren). |
| Final match-confirming variables | 9 | | See Supplemental Table 2 |
| Geographic match-confirming variables | 10 | | See Supplemental Table 1 |

1. Ackley SF, Zimmerman SC, Flatt JD, Riley AR, Sevelius J, Duchowny KA. Discordance in chromosomal and self-reported sex in the UK Biobank: Implications for transgender- and intersex-inclusive data collection. *Proc Natl Acad Sci*. 2023;120(18):e2218700120. doi:10.1073/pnas.2218700120

**Supplemental table 7: The 17 compatible household member type combinations detected in the data**

| One person reports | The other person reports | Household size limit (rule 3) |
|---|---|---|
| Spouse/Partner | Spouse/Partner | |
| Spouse/Partner, Child | Spouse/Partner, Child | |
| Spouse/Partner, Child, Grandchild | Spouse/Partner, Child, Grandchild | |
| Spouse/Partner, Child, Grandchild | Spouse/Partner, Child, Other relative | |
| Spouse/Partner, Child, Grandchild | Spouse/Partner, Child, Unrelated | |
| Spouse/Partner, Child, Other relative | Spouse/Partner, Child, Sibling | |
| Spouse/Partner, Child, Sibling | Spouse/Partner, Child, Unrelated | |
| Spouse/Partner, Grandchild | Spouse/Partner, Grandchild | |
| Spouse/Partner, Grandchild | Spouse/Partner, Other relative | |
| Spouse/Partner, Grandchild | Spouse/Partner, Unrelated | |
| Spouse/Partner, Other relative | Spouse/Partner, Other relative | <= 3 |
| Spouse/Partner, Other relative | Spouse/Partner, Parent | <= 3 |
| Spouse/Partner, Other relative | Spouse/Partner, Sibling | |
| Spouse/Partner, Other relative | Spouse/Partner, Unrelated | <= 3 |
| Spouse/Partner, Parent | Spouse/Partner, Unrelated | <= 3 |
| Spouse/Partner, Sibling | Spouse/Partner, Unrelated | |
| Spouse/Partner, Unrelated | Spouse/Partner, Unrelated | <= 3 |

**Supplemental table 8: Results of applying each criteria step in the UKB-colocated and geographically-colocated samples**

| | **Geographically-colocated sample** | **UKB-colocated sample** |
|---|---|---|
| Starting number of groups of 2 or more people, after excluding withdrawn individuals | The initial sample had 224243 people in 110954 colocated groups of size 2-21. | The initial sample had 211474 people in 105563 colocated groups of size 2-5. |
| **Step 1:** Exclude people who are missing data on "number of people in household" | Excluded 1222 people, and as a result 637 other people had their group size reduced to 1.<br><br>After excluding newly size-one groups, there were 222384 people in 110099 groups of size 2-21. | Excluded 886 people, and as a result 514 other people had their group size reduced to 1.<br><br>After excluding newly size-one groups, there were 210074 people in 104871 groups of size 2-5. |
| **Step 2**: Split colocated groups into groups where people agree on household size. One group of colocated people who report household sizes of 2, 2, 4, 4, 4 would be split into one group of 2, 2 and one group of 4, 4, 4. | 100282 groups of size 2-8 remained unsplit, and 9817 groups of size 2-21 were split into 19838 groups of size 1-16 (2-6 new groups per split group).<br><br>After removing size-one groups, there were 203560 people in 101296 groups of size 2-16. | 96583 groups of size 2-4 remained unsplit, and 8288 groups of size 2-5 were split into 16592 groups of size 1-3 (2-3 new groups per split group).<br><br>After removing size-one groups, there were 193650 people in 96751 groups of size 2-4. |
| **Step 3:** Exclude individuals whose colocation group size is greater than their self-reported household size (eg. people from colocation groups that probably combine multiple households).<br><br>Since all individuals in a group match on household size, this step excludes entire groups. | Excluded 3978 people (1576 complete groups).<br><br>After removing these groups, there were 199582 people in 99720 groups of size 2-4. | Excluded 739 people (347 complete groups).<br><br>After removing these groups, there were 192911 people in 96404 groups of size 2-4. |
| **Step 4**: Keep only people who report living with a spouse/partner | Excluded 3460 people (including 101 with missing data for household member types).<br><br>After this step, there were 196122 people in 98932 groups of size 1-4. | Excluded 3176 people (including 99 with missing data for household member types).<br><br>After this step, there were 189735 people in 95675 groups of size 1-3. |
| **Step 5**: Keep only groups of size 2 | Kept 97119 pairs. 1778 groups were excluded for size=1, and 35 groups were excluded for size > 2. | Kept 94028 pairs. 1631 groups were excluded for size=1, and 16 groups were excluded for size > 2. |
| **Step 6:** Keep only opposite-sex pairs**.** | At the beginning of this step, there were 97119 pairs.<br><br>At the end of this step, there were 96485 pairs. | At the beginning of this step, there were 94028 pairs.<br><br>At the end of this step, there were 93487 pairs. |
| **Step 7:** Exclude pairs that are $1^{st}$ or $2^{nd}$ degree relatives | Excluded 15 pairs for relatedness (kinship coefficient range: 0.089-0.249). Of the pairs that were kept, 92381 were confirmed unrelated, and 4110 pairs could not be checked.<br><br>96470 pairs remain. | Excluded 16 pairs for relatedness (kinship coefficient range: 0.089-0.249). Of the pairs that were kept, 89524 were confirmed unrelated, and 3975 pairs could not be checked.<br><br>93471 pairs remain. |

| | | |
|---|---|---|
| **Step 8**: Compatible household member types<br><br>(Details of determining compatibility are shown in Box 2) | Excluded 2147 pairs for incompatible reported household member types.<br>94323 pairs remain. | Excluded 2085 pairs for incompatible reported household member types.<br>91386 pairs remain. |
| **Step 9**: Matches on at least one of the following:<br><br>• Date of assessment center visit AND Assessment center<br>• Number of vehicles AND own vs. rent (broad) AND gas hob/cooker | Excluded 1813 pairs, 1762 pairs for incompatible responses and 51 pairs for missing data.<br><br>Of the remaining pairs:<br>9875 pairs matched on assessment date and center, 9711 pairs matched on the other variables, and 72924 pairs matched on both. | Excluded 1758 pairs, 1708 pairs for incompatible responses and 50 pairs for missing data.<br><br>Of the remaining pairs:<br>9545 pairs matched on assessment date and center, 9447 pairs matched on the other variables, and 70636 pairs matched on both. |
| **Step 10 (optional)**: Matches on the same geographic variables as the geographically-colocated sample. | Excluded 0 pairs for non-matching values on one or more geographic variables. | Excluded 350 pairs for non-matching values on one or more geographic variables. |
| **Final Sample** | 92510 pairs<br><br>Of these, 87970 pairs are also in the UKB-colocated sample. | 89278 pairs<br><br>For comparison with geographic colocation, 1072 pairs would be excluded (1072 for missing geovars and 0 for mismatches), leaving 88206 pairs. |

Supplemental Table 8 shows the results of applying each step of our criteria to the UKB sample, using both the UKB colocation file and our geographic colocation method.

The final comparison totals do not match due to other factors (such as exclusion of incorrectly-combined households in steps 3 and 5) beyond whether each colocation method considers a pair to be colocated.

**Supplemental Table 9a: Proportion of individuals in geographic pairs that are not UKB colocated, by assessment year**

| Assessment year | Proportion of individuals in pairs whose pairs are not in the UKB colocation file |
|---|---|
| 2006 | 0.079 |
| 2007 | 0.070 |
| 2008 | 0.056 |
| 2009 | 0.043 |
| 2010 | 0.035 |

Logistic regression: OR 0.782, p=1.03e-88

**Supplemental Table 9b: Proportion of individuals in geographic pairs that are not UKB-colocated, by assessment center**

| Assessment center | Proportion of individuals in pairs whose pairs are not in the UKB colocation file |
|---|---|
| Barts | 0.082 |
| Birmingham | 0.042 |
| Bristol | 0.044 |
| Bury | 0.054 |
| Cardiff | 0.084 |
| Croydon | 0.039 |
| Edinburgh | 0.047 |
| Glasgow | 0.085 |
| Hounslow | 0.042 |
| Leeds | 0.045 |
| Liverpool | 0.049 |
| Manchester | 0.070 |
| Middlesborough | 0.039 |
| Newcastle | 0.063 |
| Nottingham | 0.035 |
| Oxford | 0.044 |
| Reading | 0.045 |
| Sheffield | 0.033 |
| Stockport | 0.079 |
| Stoke | 0.058 |
| Swansea | 0.039 |
| Wrexham | 0.027 |

p (unadjusted): 6.28e-129

From a likelihood ratio test comparing a logistic model with assessment center to an intercept-only model.

p (adjusted for assessment year): 8.78e-66

From a likelihood ratio test comparing a logistic model with assessment center and assessment year to a model with assessment year only.

**Supplemental Table 9c: Proportion of individuals in geographic pairs that are not UKB-colocated, by housing type**

| | |
|---|---|
| Not Flat/apartment | 0.047 |
| Flat/apartment | 0.115 |

Logistic regression: OR 2.63, p=1.21e-103

Supplemental Tables 9a-9c show the proportion of individuals in geographic pairs that are not colocated according to the UKB colocation file, stratified by other variables such as assessment year or assessment center. Analyses were performed using individuals as the unit of observation, to allow for pairs that differed on factors such as assessment year or center. Only individuals who were members of a geographically-colocated pair were included in the analysis.

Logistic regression models examine odds of being in a UKB-non-colocated pair, as predicted by the other variable. The reference group was individuals in UKB-confirmed geographic pairs, individuals who were not in geographic pairs were not included in the models.

### Supplemental table 10: Phenotypes and traits

| | UKB Field ID | Details |
|---|---|---|
| **Anthropometric traits** | | |
| Body mass index (kg/m$^2$) | 21002 & 50 | Weight (kilograms)/height(meters)$^2$ |
| C-reactive protein (log10) | 30710 | log10 transformed |
| Cholesterol (mmol/L) | 30690 | |
| Diastolic blood pressure (mmHg) | 4079 | Mean of two measurements, f.4079.0.0 and f.4079.0.1 |
| Fluid IQ | 20016 | |
| Forced expiratory volume in 1 second (L) | 20150 | |
| Forced vital capacity (L) | 20151 | |
| Hand grip strength (Kg) | 46 & 47 | 46: Left grip strength; 47: Right grip strength. Grip strength was calculated as the maximum of these values for a given participant. |
| Handedness | 1707 | Right-handed coded as 0, left-handed coded as 1, ambidextrous excluded. |
| Heel bone mineral density<br>(T-score standard deviations) | 78 | |
| Height (cm) | 50 | |
| Hemoglobin A1c (mmol/mol) | 30750 | |
| High-density lipoprotein (HDL) (mmol/L) | 30760 | |
| Hip circumference (cm) | 49 | |
| Low-density lipoprotein (LDL) (mmol/L) | 30780 | |
| Peak expiratory flow (L/min) | 3064 | Mean of up to three measurements, f.3064.0.0, f.3064.0.1, f.3064.0.2 |
| Pulse (bpm) | 102 | Mean of two measurements, f.102.0.0 and f.102.0.1 |
| Red blood cell count ($10^{12}$ cells/L) | 30010 | |
| Systolic blood pressure (mmHg) | 4080 | Mean of two measurements, f.4079.0.0 and f.4079.0.1 |
| Waist circumference (cm) | 48 | |
| Waist-hip ratio | 48 & 49 | Waist circumference (centimeters)/hip circumference (centimeters) |
| Weight (kg) | 21002 | |
| White blood cell count ($10^9$ cells/L) | 30000 | |
| Whole body impedance (ohms) | 23106 | |
| **Behavioral traits** | | |
| Age at first intercourse | 2139 | |
| Chronotype | 1180 | Responses of “Definitely a morning person” or “More of a morning person than an evening person” were coded as 0.<br>Responses of “More of an evening person than a morning person” or “Definitely an evening person” were coded as 1. |
| Number of sexual partners | 2149 | |
| Risk taking | 2040 | |

| | | |
|---|---|---|
| Sleep duration (hours) | 1160 | Includes naps |
| Time spent using computer (hours/day) | 1080 | Does not include time during work. Those who responded "less than an hour a day" were coded as 0. |
| Time spent watching TV (hours/day) | 1070 | Those who responded "less than an hour a day" were coded as 0. |
| Water intake (glasses/day) | 1528 | Those who responded "less than one" were coded as 0. |
| **Demographic/Family traits** | | |
| Age | 21003 | |
| Birthplace (east coordinate) | 130 | |
| Birthplace (north coordinate) | 129 | |
| Breastfed | 1677 | |
| Educational attainment | 6138 & 10722 | Which of the following qualifications do you have? (You can select more than one). Recoded such that:<br>1 (College or university degree) = 20<br>2 (A/AS Levels or equivalent) = 13<br>3 (O Levels/GCSEs or equivalent) = 10<br>4 (CSEs or equivalent) = 10<br>5 (NVQ, HND, HNC or equivalent) = Age_Completed_Edu (Field ID 845) - 5 *if* Age_Completed_Edu - 5 < 20; otherwise, assigned a value of 19<br>6 (Other professional qualification) = 15<br>-7 (None of the above) = 7<br>Those who reported completing their education before the age of 12 (according to Field ID 845) were excluded.<br>Coding was based on that used by the Okbay, 2022 educational attainment genome-wide association study (https://doi.org/10.1038/s41588-022-01016-z) but was modified to limit possible values for “NVQ, HND, HNC or equivalent” to 19 or lower. |
| Mother smoked during pregnancy | 1787 | |
| Number of children | 2405 & 2734 | 2405: How many children have you fathered? 2734: How many children have you given birth to? |
| Number of siblings | 1873 & 1883 | Sum of “Number of brothers” and “Number of sisters” |
| **Health behaviors** | | |
| Drink alcohol | 20117 | Current drinkers coded as 1, former drinkers and never-drinkers coded as 0. |
| Drinks full-fat or semi-skimmed milk | 1418 | Coded as 1 if a participant reported a milk type of “Full Cream” or “Semi-skimmed”, or 0 if a participant reported “Skimmed milk”, “Soya milk”, “Other milk type”, or “Never/rarely have milk” |
| Eats processed meat >= 1x week | 1349 | Coded as 1 if a participant reported a frequency of once a week (or more) and 0 if a participant responded “Never” or “Less than once a week” |
| Ever smoked | 20160 | Derived using variables "current tobacco smoking" and "past tobacco smoking." Coded as 1 if current tobacco smoking=most days or occasionally OR Past tobacco smoking=most days or occasionally or tried once or twice. Coded as 0 if Current tobacco smoking= no AND Past tobacco smoking=never. |
| Fruit Intake (pieces/day) | 1309, 1319 | Sum of “Fresh fruit intake” and “Dried fruit intake” |
| Moderate physical activity (days/week) | 884 | |
| Smoked at least 100 times | 2644, 1239, & 1249 | 2644: In your lifetime, have you smoked a total of at least 100 times (collected from participants who indicated they smoked occasionally or just tried once or twice)? People who were not asked this question because they currently smoke on most or all days (Field ID 1239) were coded as 1, as were people who were not asked this question because they used to smoke on all or most days (Field ID 1249). People who were not asked this question because they never smoked were coded as 0, while people who declined to answer the current smoking question but said "Never" to former smoking were excluded. |
| Smoking status (Current) | 20116 | Current smokers coded as 1, former smokers and never smokers coded as 0. |
| Smoking status (Former) | 20116 | Former smokers coded as 1, current smokers and never smokers coded as 0. |
| Smoking status (Never) | 20116 | Never smokers coded as 1, current smokers and former smokers coded as 0. |

| | | |
|---|---|---|
| Sun protection | 2267 | Coded as 1 if a participant indicated they “Always” or “Often” used sunscreen or other sun protection when out in the sun and 0 if a participant responded that they “Sometimes” or “Never/Rarely” do this. Participants who responded “Do not go out in sunshine” were set to missing. |
| Vegetable Intake (Tablespoons/day) | 1289, 1299 | Sum of “Cooked vegetable intake” and “Salad / Raw vegetable intake” |
| Walking (minutes/day) | 874 & 10953 | |
| **Health traits** | | |
| Asthma | 6152 | Coded as 1 if a participant’s responses included asthma. |
| Basal metabolic rate (kJ) | 23105 | |
| Dental problems | 6149 | Coded as 1 if a participant’s responses included mouth ulcers, painful gums, bleeding gums, loose teeth, toothache, or dentures |
| Diabetes | 2443 | |
| Difficulty hearing | 2247 | Do you have any difficulty with your hearing? Those who responsed "yes" or "I am completely deaf" were coded as 1. |
| Ever had tinnitus | 4803 | Responses indicating current/previous tinnitus are coded as 1, responses indicating participant has never experienced tinnitus are coded as 0. |
| Hay fever, allergic rhinitis, or eczema | 6152 | Coded as 1 if a participant’s responses included "Hayfever, allergic rhinitis or eczema" |
| High blood pressure | 6150 | Coded as 1 if a participant’s responses included “High blood pressure” |
| Long-term illness | 2188 | |
| Number of illnesses | 135 | |
| Number of medications | 137 | |
| Wears Glasses | 2207 | |
| **Psychological traits** | | |
| Anxious feelings | 1980 | |
| Doctor-diagnosed anxiety or depression | 2100 & 2090 | An answer of “yes” on either phenotype was coded as 1, an answer of “no” on both was coded as 0. |
| Ever depressed for at least one week | 4598 | |
| Fed-up feelings | 1960 | |
| Guilt | 2030 | |
| High strung | 1990 | |
| Hurt feelings | 1950 | |
| Irritability | 1940 | |
| Loneliness | 2020 | |
| Miserableness | 1930 | |
| Mood swings | 1920 | |
| Nerves | 2010 | |
| Nervous feelings | 1970 | |
| Neuroticism | 20127 | |
| Worry too long after embarrassment | 2000 | |

**Supplemental Table 11: Trait correlations and data-derived confidence intervals for the UKB-colocated and geographically-colocated samples**

| | | Geographically-colocated pairs | | | UKB-colocated pairs | | |
|---|---|---|---|---|---|---|---|
| | **Correlation Type** | **N** | **Mean (males and females) (95% CI)** | **Spouse/partner correlation (95% CI)** | **N** | **Mean (males and females) (95% CI)** | **Spouse/partner correlation (95% CI)** |
| **Anthropometric traits** | | | | | | | |
| Body mass index | Pearson | 89852 | 27.219 (27.200,27.238) | 0.240 (0.234,0.247) | 86703 | 27.207 (27.188,27.227) | 0.240 (0.233,0.246) |
| C-reactive protein (log10) | Pearson | 81586 | 0.135 (0.133,0.137) | 0.132 (0.125,0.139) | 78722 | 0.134 (0.132,0.136) | 0.132 (0.125,0.138) |
| Cholesterol | Pearson | 81946 | 5.701 (5.696,5.707) | 0.078 (0.071,0.085) | 79086 | 5.701 (5.696,5.707) | 0.079 (0.072,0.086) |
| Diastolic blood pressure | Pearson | 84787 | 82.548 (82.501,82.594) | 0.102 (0.095,0.109) | 81890 | 82.543 (82.496,82.590) | 0.102 (0.095,0.108) |
| Fluid IQ | Pearson | 29858 | 6.115 (6.098,6.132) | 0.225 (0.214,0.236) | 29150 | 6.119 (6.102,6.136) | 0.224 (0.213,0.235) |
| Forced expiratory volume in 1 second | Pearson | 51500 | 2.875 (2.871,2.880) | 0.269 (0.261,0.277) | 49805 | 2.876 (2.871,2.880) | 0.267 (0.259,0.275) |
| Forced vital capacity | Pearson | 51498 | 3.803 (3.797,3.809) | 0.261 (0.253,0.269) | 49818 | 3.803 (3.797,3.809) | 0.260 (0.251,0.268) |
| Hand grip strength | Pearson | 90974 | 33.428 (33.377,33.480) | 0.180 (0.174,0.186) | 87700 | 33.405 (33.353,33.457) | 0.179 (0.172,0.185) |
| Handedness | tetrachoric | 89547 | 0.095 (0.094,0.097) | -0.003 (-0.023,0.017) | 86432 | 0.095 (0.094,0.097) | -0.007 (-0.027,0.013) |
| Heel bone mineral density | Pearson | 48976 | -0.349 (-0.356,-0.343) | 0.039 (0.031,0.048) | 47180 | -0.350 (-0.357,-0.343) | 0.039 (0.030,0.048) |
| Height | Pearson | 91437 | 169.035 (168.993,169.077) | 0.258 (0.252,0.265) | 88233 | 169.054 (169.011,169.097) | 0.259 (0.252,0.265) |
| Hemoglobin A1c | Pearson | 79853 | 35.437 (35.416,35.458) | 0.132 (0.125,0.139) | 77074 | 35.437 (35.416,35.459) | 0.132 (0.125,0.139) |
| High-density lipoprotein (HDL) | Pearson | 69464 | 1.432 (1.430,1.434) | 0.154 (0.147,0.161) | 67041 | 1.433 (1.431,1.435) | 0.154 (0.147,0.162) |
| Hip circumference | Pearson | 90087 | 103.026 (102.990,103.063) | 0.185 (0.179,0.191) | 86953 | 103.017 (102.980,103.054) | 0.185 (0.178,0.191) |
| Low-density lipoprotein (LDL) | Pearson | 81747 | 3.565 (3.561,3.570) | 0.067 (0.061,0.074) | 78886 | 3.565 (3.561,3.570) | 0.068 (0.061,0.075) |
| Peak expiratory flow | Pearson | 76823 | 377.840 (377.212,378.468) | 0.183 (0.176,0.190) | 74193 | 378.113 (377.474,378.752) | 0.182 (0.175,0.189) |
| Pulse | Pearson | 84131 | 68.175 (68.124,68.225) | 0.113 (0.106,0.120) | 81269 | 68.158 (68.107,68.210) | 0.112 (0.106,0.119) |
| Red blood cell count | Pearson | 84703 | 4.537 (4.535,4.539) | 0.096 (0.090,0.103) | 81763 | 4.538 (4.536,4.539) | 0.096 (0.089,0.103) |
| Systolic blood pressure | Pearson | 84445 | 139.210 (139.125,139.295) | 0.160 (0.154,0.167) | 81572 | 139.235 (139.149,139.322) | 0.161 (0.154,0.167) |
| Waist circumference | Pearson | 90641 | 90.443 (90.385,90.501) | 0.261 (0.255,0.267) | 87477 | 90.430 (90.371,90.489) | 0.261 (0.255,0.267) |
| Waist-hip ratio | Pearson | 91273 | 0.876 (0.875,0.876) | 0.187 (0.181,0.193) | 88084 | 0.876 (0.875,0.876) | 0.187 (0.181,0.194) |
| Weight | Pearson | 90015 | 78.042 (77.974,78.109) | 0.216 (0.210,0.223) | 86858 | 78.027 (77.959,78.096) | 0.216 (0.210,0.222) |
| White blood cell count | Pearson | 85040 | 6.808 (6.800,6.816) | 0.155 (0.148,0.161) | 82128 | 6.809 (6.801,6.817) | 0.155 (0.148,0.162) |
| Whole body impedance | Pearson | 88646 | 595.233 (594.823,595.643) | 0.136 (0.130,0.143) | 85562 | 595.277 (594.860,595.695) | 0.135 (0.129,0.142) |
| **Behavioral traits** | | | | | | | |
| Age at first intercourse | Pearson | 72593 | 19.053 (19.037,19.070) | 0.416 (0.410,0.422) | 70082 | 19.063 (19.047,19.080) | 0.417 (0.411,0.423) |
| Chronotype | tetrachoric | 73820 | 0.360 (0.358,0.363) | -0.175 (-0.187,-0.164) | 71187 | 0.361 (0.358,0.363) | -0.176 (-0.188,-0.164) |
| Number of sexual partners | Spearman | 66595 | 4.125 (4.095,4.156) | 0.503 (0.497,0.509) | 64360 | 4.109 (4.078,4.140) | 0.502 (0.496,0.507) |

| | | | | | | | |
|---|---|---|---|---|---|---|---|
| Risk taking | tetrachoric | 86890 | 0.228 (0.226,0.230) | 0.068 (0.055,0.082) | 83873 | 0.227 (0.225,0.229) | 0.068 (0.055,0.082) |
| Sleep duration | Pearson | 89791 | 7.240 (7.235,7.244) | 0.119 (0.112,0.125) | 86696 | 7.240 (7.235,7.244) | 0.120 (0.113,0.126) |
| Time spent using computer | Spearman | 90343 | 0.917 (0.912,0.923) | 0.101 (0.095,0.108) | 87217 | 0.918 (0.912,0.924) | 0.101 (0.094,0.107) |
| Time spent watching TV | Spearman | 90814 | 2.834 (2.827,2.841) | 0.446 (0.441,0.451) | 87647 | 2.832 (2.825,2.839) | 0.446 (0.441,0.452) |
| Water intake | Spearman | 90939 | 2.459 (2.450,2.469) | 0.213 (0.207,0.219) | 87773 | 2.457 (2.448,2.467) | 0.214 (0.208,0.220) |
| **Demographic/Family traits** | | | | | | | |
| Age | Pearson | 92510 | 57.817 (57.784,57.850) | 0.853 (0.852,0.855) | 89278 | 57.845 (57.812,57.879) | 0.855 (0.853,0.856) |
| Birthplace (east coordinate) | Pearson | 79842 | 413413.080 (413045.357,413780.803) | 0.499 (0.493,0.504) | 77083 | 413751.343 (413377.447,414125.239) | 0.497 (0.492,0.503) |
| Birthplace (north coordinate) | Pearson | 79815 | 360507.316 (359748.228,361266.404) | 0.652 (0.648,0.656) | 77052 | 360144.282 (359372.337,360916.227) | 0.650 (0.646,0.654) |
| Breastfed | tetrachoric | 55000 | 0.741 (0.738,0.744) | 0.292 (0.278,0.306) | 53054 | 0.742 (0.739,0.744) | 0.291 (0.276,0.305) |
| Educational attainment | Pearson | 90287 | 13.810 (13.787,13.832) | 0.492 (0.487,0.497) | 87175 | 13.826 (13.803,13.849) | 0.492 (0.487,0.497) |
| Mother smoked during pregnancy | tetrachoric | 69264 | 0.296 (0.294,0.299) | 0.147 (0.135,0.160) | 66895 | 0.295 (0.293,0.298) | 0.147 (0.135,0.160) |
| Number of children | Spearman | 91150 | 1.947 (1.942,1.952) | 0.717 (0.714,0.720) | 88018 | 1.947 (1.943,1.952) | 0.721 (0.717,0.724) |
| Number of siblings | Spearman | 91741 | 2.017 (2.009,2.025) | 0.099 (0.093,0.106) | 88543 | 2.013 (2.004,2.021) | 0.099 (0.092,0.105) |
| **Health behaviors** | | | | | | | |
| Drinks alcohol | tetrachoric | 92374 | 0.937 (0.936,0.939) | 0.572 (0.557,0.588) | 89156 | 0.938 (0.937,0.939) | 0.572 (0.556,0.588) |
| Drinks full-fat or semi-skimmed milk | tetrachoric | 92415 | 0.710 (0.708,0.712) | 0.747 (0.741,0.754) | 89188 | 0.710 (0.708,0.712) | 0.748 (0.742,0.754) |
| Eats processed meat >= 1x week | tetrachoric | 92254 | 0.639 (0.637,0.642) | 0.420 (0.411,0.430) | 89033 | 0.639 (0.637,0.642) | 0.423 (0.414,0.433) |
| Ever smoked | tetrachoric | 91872 | 0.591 (0.589,0.593) | 0.242 (0.232,0.252) | 88656 | 0.590 (0.588,0.592) | 0.243 (0.233,0.254) |
| Fruit intake | Pearson | 74915 | 2.941 (2.931,2.951) | 0.253 (0.246,0.259) | 72511 | 2.961 (2.951,2.972) | 0.252 (0.245,0.259) |
| Moderate physical activity (days/week) | Pearson | 83698 | 3.616 (3.605,3.627) | 0.116 (0.109,0.122) | 80788 | 3.616 (3.605,3.627) | 0.115 (0.109,0.122) |
| Smoked at least 100 times | tetrachoric | 89749 | 0.436 (0.433,0.438) | 0.364 (0.355,0.374) | 86610 | 0.435 (0.432,0.437) | 0.367 (0.357,0.377) |
| Smoking status (Current) | tetrachoric | 91850 | 0.073 (0.072,0.075) | 0.514 (0.499,0.530) | 88634 | 0.073 (0.072,0.074) | 0.511 (0.495,0.526) |
| Smoking status (Former) | tetrachoric | 91850 | 0.366 (0.364,0.369) | 0.311 (0.301,0.321) | 88634 | 0.366 (0.364,0.368) | 0.314 (0.303,0.324) |
| Smoking status (Never) | tetrachoric | 91850 | 0.560 (0.558,0.562) | 0.366 (0.356,0.375) | 88634 | 0.561 (0.559,0.564) | 0.368 (0.358,0.377) |
| Sun protection | tetrachoric | 91345 | 0.587 (0.584,0.589) | 0.242 (0.232,0.252) | 88181 | 0.587 (0.585,0.589) | 0.243 (0.233,0.254) |
| Vegetable intake | Pearson | 87465 | 4.731 (4.720,4.743) | 0.256 (0.250,0.263) | 84419 | 4.733 (4.722,4.745) | 0.257 (0.250,0.263) |
| Walking (minutes/day) | Spearman | 67750 | 59.271 (58.908,59.634) | 0.186 (0.179,0.193) | 65398 | 59.217 (58.848,59.585) | 0.188 (0.181,0.195) |
| **Health traits** | | | | | | | |
| Asthma | tetrachoric | 92267 | 0.109 (0.107,0.110) | 0.003 (-0.015,0.021) | 89051 | 0.109 (0.107,0.110) | -0.004 (-0.022,0.015) |
| Basal metabolic rate | Pearson | 87815 | 6660.930 (6654.704,6667.155) | 0.206 (0.200,0.213) | 84764 | 6661.015 (6654.675,6667.355) | 0.206 (0.199,0.212) |
| Dental problems | tetrachoric | 91756 | 0.382 (0.379,0.384) | 0.145 (0.135,0.155) | 88574 | 0.381 (0.379,0.383) | 0.145 (0.135,0.156) |
| Diabetes | tetrachoric | 92104 | 0.050 (0.049,0.051) | 0.196 (0.170,0.221) | 88892 | 0.050 (0.049,0.051) | 0.199 (0.173,0.225) |

| | | | | | | | |
|---|---|---|---|---|---|---|---|
| Difficulty hearing | tetrachoric | 85492 | 0.276 (0.274,0.278) | -0.070 (-0.082,-0.058) | 82514 | 0.276 (0.274,0.278) | -0.069 (-0.081,-0.056) |
| Ever had tinnitus | tetrachoric | 30440 | 0.292 (0.288,0.295) | 0.065 (0.046,0.085) | 29722 | 0.291 (0.288,0.295) | 0.065 (0.045,0.085) |
| Hay fever, allergic rhinitis, or eczema | tetrachoric | 92267 | 0.224 (0.222,0.226) | 0.011 (-0.001,0.024) | 89051 | 0.224 (0.222,0.226) | 0.015 (0.002,0.027) |
| High blood pressure | tetrachoric | 92191 | 0.282 (0.280,0.284) | 0.097 (0.085,0.108) | 88972 | 0.282 (0.280,0.284) | 0.099 (0.087,0.111) |
| Long-term illness | tetrachoric | 88682 | 0.314 (0.312,0.316) | 0.136 (0.125,0.147) | 85600 | 0.313 (0.311,0.316) | 0.135 (0.124,0.146) |
| Number of illnesses | Spearman | 91752 | 1.813 (1.805,1.821) | 0.159 (0.153,0.166) | 88543 | 1.812 (1.804,1.820) | 0.160 (0.153,0.166) |
| Number of medications | Spearman | 91718 | 2.410 (2.399,2.421) | 0.219 (0.213,0.225) | 88516 | 2.406 (2.395,2.418) | 0.218 (0.212,0.224) |
| Wears Glasses | tetrachoric | 92356 | 0.916 (0.915,0.917) | 0.381 (0.365,0.397) | 89129 | 0.916 (0.915,0.917) | 0.381 (0.364,0.398) |
| **Psychological traits** | | | | | | | |
| Anxious feelings | tetrachoric | 88119 | 0.570 (0.568,0.572) | -0.099 (-0.109,-0.088) | 85034 | 0.570 (0.567,0.572) | -0.101 (-0.112,-0.090) |
| Doctor-diagnosed anxiety or depression | tetrachoric | 91373 | 0.307 (0.305,0.309) | 0.152 (0.141,0.163) | 88188 | 0.306 (0.304,0.308) | 0.152 (0.140,0.163) |
| Ever depressed for at least one week | tetrachoric | 30283 | 0.477 (0.473,0.480) | 0.141 (0.123,0.159) | 29564 | 0.476 (0.472,0.480) | 0.141 (0.123,0.158) |
| Fed-up feelings | tetrachoric | 88733 | 0.377 (0.375,0.379) | 0.150 (0.140,0.161) | 85660 | 0.376 (0.374,0.378) | 0.151 (0.140,0.162) |
| Guilt | tetrachoric | 87919 | 0.266 (0.264,0.268) | 0.059 (0.047,0.072) | 84899 | 0.265 (0.263,0.267) | 0.061 (0.049,0.074) |
| High strung | tetrachoric | 86972 | 0.166 (0.164,0.167) | 0.022 (0.007,0.037) | 83987 | 0.165 (0.163,0.167) | 0.022 (0.007,0.038) |
| Hurt feelings | tetrachoric | 87436 | 0.551 (0.549,0.553) | 0.045 (0.034,0.056) | 84369 | 0.550 (0.548,0.553) | 0.043 (0.032,0.054) |
| Irritability | tetrachoric | 84489 | 0.289 (0.287,0.291) | -0.019 (-0.031,-0.007) | 81555 | 0.288 (0.286,0.291) | -0.019 (-0.031,-0.007) |
| Loneliness | tetrachoric | 90089 | 0.116 (0.114,0.117) | 0.160 (0.143,0.177) | 86964 | 0.115 (0.113,0.116) | 0.157 (0.140,0.174) |
| Miserableness | tetrachoric | 89341 | 0.409 (0.407,0.411) | 0.108 (0.098,0.119) | 86237 | 0.408 (0.405,0.410) | 0.109 (0.098,0.120) |
| Mood swings | tetrachoric | 88062 | 0.439 (0.436,0.441) | 0.058 (0.048,0.069) | 84983 | 0.438 (0.435,0.440) | 0.059 (0.048,0.070) |
| Nerves | tetrachoric | 86248 | 0.204 (0.202,0.205) | 0.077 (0.063,0.090) | 83239 | 0.203 (0.201,0.205) | 0.079 (0.065,0.092) |
| Nervous feelings | tetrachoric | 88025 | 0.236 (0.234,0.238) | 0.033 (0.020,0.045) | 84952 | 0.235 (0.233,0.237) | 0.036 (0.023,0.049) |
| Neuroticism | Spearman | 61557 | 3.966 (3.948,3.983) | 0.042 (0.034,0.050) | 59442 | 3.959 (3.941,3.977) | 0.042 (0.034,0.050) |
| Worry too long after embarrassment | tetrachoric | 85213 | 0.474 (0.471,0.476) | 0.022 (0.011,0.033) | 82222 | 0.474 (0.471,0.476) | 0.021 (0.010,0.032) |

These values are used in Figure 1, but could not be used for statistical comparisons due to sample overlap. P-values for correlations based on bootstrap comparisons are included in Supplemental Table 18.

**Supplemental table 12: Subsets and comparison groups for examining the potential impacts of other eligibility variables used in previous studies**

| Subgroup | Comparison group |
|---|---|
| 20777 pairs with missing data for self-reported household income | 68501 pairs with complete data for self-reported household income |
| 15295 pairs with differing self-reported household income categories | 53206 pairs with matching self-reported household income |
| 6249 pairs that differ by 5+ years for length of time at current address | 82821 pairs that differ by 0-4 years for length of time at current address |
| 3897 pairs with an age difference >= 10 years | 85381 pairs with an age difference < 10 years |

**Supplemental table 13a: Matches and mismatches on self-reported household income in the final eligible UKB-colocated sample**

| | < £18000 | £ 18000 - 30999 | £31000 - 51999 | £52000 - 100000 | > £ 100000 | Missing |
|---|---|---|---|---|---|---|
| **< £18000** | 7694 | 3182 | 316 | 67 | 17 | 978 |
| **£18000 - 30999** | | 13388 | 5021 | 427 | 33 | 1237 |
| **£31000 - 51999** | | | 15567 | 4597 | 83 | 1003 |
| **£52000 - 100000** | | | | 13509 | 1552 | 588 |
| **> £100000** | | | | | 3048 | 92 |
| **Missing** | | | | | | 16879 |

**Supplemental table 13b: Matches and mismatches on self-reported household income in a sample with additional eligibility filtering**

| | < £18000 | £ 18000 - 30999 | £31000 - 51999 | £52000 - 100000 | > £ 100000 | Missing |
|---|---|---|---|---|---|---|
| **< £18000** | 6693 | 2560 | 234 | 51 | 15 | 791 |
| **£18000 - 30999** | | 11481 | 3797 | 285 | 24 | 974 |
| **£31000 - 51999** | | | 12663 | 3194 | 49 | 728 |
| **£52000 - 100000** | | | | 10097 | 980 | 417 |
| **> £100000** | | | | | 2081 | 48 |
| **Missing** | | | | | | 13286 |

Supplemental tables 13a and 13b demonstrate the range of (mis)matched responses for household income in UKB-colocated couples meeting our full eligibility criteria (13a) and among pairs meeting an even more stringent criteria requiring them to fulfill both parts of the step 9 match-confirming criterion (13b).

A broad range of mismatches exist, including mismatches between the lowest category (< £18,000) and highest category (> £100,000), and these mismatches persist even in a very stringently-filtered subset of our eligible pairs. This suggests that many of the mismatches result from another factor (such as individuals misunderstanding the question and reporting their personal income instead of their household income) rather than from falsely-matched pairs.

Responses are shown independent of partner number/order, so all response combinations appear only in the upper-right triangle of the table

**Supplemental table 14: Comparison between pairs with missing data on income and pairs with complete income data**

| | Trait means | | | Trait correlations | | |
|---|---|---|---|---|---|---|
| | **Excluded subset** | **Retained subset** | **P** | **Excluded subset** | **Retained subset** | **P** |
| **Anthropometric traits** | | | | | | |
| Body mass index | 27.44 (27.40,27.48) | 27.14 (27.12,27.16) | 1.61e-36 ** | 0.229 (0.216,0.242) | 0.242 (0.235,0.249) | 0.099 |
| C-reactive protein (log10) | 0.17 (0.17,0.18) | 0.12 (0.12,0.13) | 7.11e-75 ** | 0.119 (0.105,0.133) | 0.133 (0.125,0.141) | 0.097 |
| Cholesterol | 5.70 (5.68,5.71) | 5.70 (5.70,5.71) | 0.237 | 0.084 (0.070,0.099) | 0.079 (0.071,0.087) | 0.508 |
| Diastolic blood pressure | 82.69 (82.59,82.79) | 82.50 (82.45,82.55) | 8E-04 | 0.077 (0.063,0.091) | 0.109 (0.101,0.117) | 9.27E-05 |
| Fluid IQ | 5.72 (5.69,5.76) | 6.22 (6.20,6.24) | 2.17e-119 ** | 0.195 (0.171,0.219) | 0.222 (0.210,0.235) | 0.048 |
| Forced expiratory volume in 1 second | 2.77 (2.76,2.78) | 2.90 (2.90,2.91) | 4.08e-122 ** | 0.243 (0.225,0.260) | 0.265 (0.256,0.274) | 0.027 |
| Forced vital capacity | 3.69 (3.67,3.70) | 3.84 (3.83,3.84) | 2.85e-99 ** | 0.232 (0.215,0.250) | 0.259 (0.250,0.268) | 0.009 |
| Hand grip strength | 32.57 (32.46,32.67) | 33.66 (33.60,33.72) | 2.92e-67 ** | 0.165 (0.151,0.178) | 0.178 (0.171,0.186) | 0.081 |
| Handedness | 0.09 (0.09,0.10) | 0.10 (0.09,0.10) | 0.067 | -0.028 (-0.071,0.015) | -0.002 (-0.024,0.021) | 0.290 |
| Heel bone mineral density | -0.40 (-0.41,-0.38) | -0.34 (-0.34,-0.33) | 9.09e-14 ** | 0.047 (0.029,0.066) | 0.036 (0.026,0.046) | 0.288 |
| Height | 168.33 (168.24,168.42) | 169.27 (169.22,169.32) | 2.05e-73 ** | 0.267 (0.255,0.280) | 0.252 (0.245,0.259) | 0.039 |
| Hemoglobin A1c | 35.91 (35.86,35.95) | 35.30 (35.27,35.32) | 3.79e-115 ** | 0.126 (0.112,0.141) | 0.130 (0.122,0.138) | 0.629 |
| High-density lipoprotein (HDL) | 1.42 (1.42,1.43) | 1.44 (1.43,1.44) | 1.75e-07 ** | 0.159 (0.144,0.175) | 0.153 (0.144,0.161) | 0.461 |
| Hip circumference | 103.17 (103.09,103.25) | 102.97 (102.93,103.01) | 1.13e-05 ** | 0.180 (0.167,0.194) | 0.186 (0.178,0.193) | 0.494 |
| Low-density lipoprotein (LDL) | 3.56 (3.55,3.57) | 3.57 (3.56,3.57) | 0.209 | 0.071 (0.057,0.086) | 0.068 (0.060,0.076) | 0.683 |
| Peak expiratory flow | 363.18 (361.84,364.52) | 382.47 (381.75,383.20) | 2.97e-135 ** | 0.165 (0.151,0.180) | 0.180 (0.172,0.188) | 0.080 |
| Pulse | 68.44 (68.33,68.55) | 68.07 (68.02,68.13) | 5.33e-09 ** | 0.109 (0.094,0.123) | 0.113 (0.106,0.121) | 0.565 |
| Red blood cell count | 4.54 (4.54,4.55) | 4.54 (4.53,4.54) | 0.031 | 0.098 (0.084,0.112) | 0.096 (0.088,0.103) | 0.783 |
| Systolic blood pressure | 141.01 (140.83,141.19) | 138.70 (138.61,138.80) | 4.36e-107 ** | 0.138 (0.124,0.152) | 0.165 (0.157,0.172) | 0.001 |
| Waist circumference | 91.07 (90.95,91.20) | 90.24 (90.17,90.30) | 8.36e-32 ** | 0.252 (0.239,0.264) | 0.263 (0.256,0.270) | 0.140 |
| Waist-hip ratio | 0.88 (0.88,0.88) | 0.87 (0.87,0.87) | 4.40e-40 ** | 0.181 (0.168,0.195) | 0.187 (0.180,0.194) | 0.470 |
| Weight | 77.96 (77.82,78.10) | 78.05 (77.97,78.13) | 0.290 | 0.212 (0.199,0.225) | 0.217 (0.210,0.224) | 0.488 |
| White blood cell count | 6.88 (6.86,6.89) | 6.79 (6.78,6.80) | 1.43e-19 ** | 0.147 (0.133,0.161) | 0.157 (0.150,0.165) | 0.191 |
| Whole body impedance | 593.71 (592.84,594.59) | 595.75 (595.27,596.22) | 5.96e-05 ** | 0.162 (0.148,0.175) | 0.127 (0.119,0.135) | 1.28e-05 ** |
| **Behavioral traits** | | | | | | |
| Age at first intercourse | 19.14 (19.10,19.18) | 19.05 (19.03,19.06) | 2.53e-05 ** | 0.436 (0.422,0.450) | 0.413 (0.406,0.419) | 0.004 |
| Chronotype | 0.35 (0.35,0.36) | 0.36 (0.36,0.37) | 0.015 | -0.157 (-0.183,-0.131) | -0.181 (-0.195,-0.168) | 0.099 |
| Number of sexual partners | 3.35 (3.29,3.41) | 4.27 (4.24,4.31) | 1.70e-134 ** | 0.488 (0.474,0.502) | 0.499 (0.493,0.506) | 0.147 |

| | | | | | | |
|---|---|---|---|---|---|---|
| Risk taking | 0.21 (0.21,0.21) | 0.23 (0.23,0.23) | 1.37e-20 ** | 0.045 (0.014,0.075) | 0.073 (0.058,0.089) | 0.100 |
| Sleep duration | 7.26 (7.25,7.27) | 7.23 (7.23,7.24) | 2.84e-07 ** | 0.111 (0.097,0.125) | 0.123 (0.115,0.130) | 0.136 |
| Time spent using computer | 0.84 (0.82,0.85) | 0.94 (0.94,0.95) | 3.01e-51 ** | 0.101 (0.087,0.115) | 0.098 (0.091,0.106) | 0.730 |
| Time spent watching TV | 3.05 (3.03,3.06) | 2.77 (2.76,2.78) | 3.21e-224 ** | 0.388 (0.376,0.400) | 0.460 (0.454,0.466) | 2.07e-27 ** |
| Water intake | 2.42 (2.40,2.44) | 2.47 (2.46,2.48) | 6.16e-05 ** | 0.212 (0.199,0.225) | 0.215 (0.208,0.222) | 0.703 |
| **Demographic/Family traits** | | | | | | |
| Age | 59.76 (59.69,59.82) | 57.27 (57.23,57.30) | 0.00e+00 ** | 0.841 (0.837,0.845) | 0.854 (0.852,0.856) | 4.48e-09 ** |
| Birthplace (east coordinate) | 412095.10 (411363.29,412826.92) | 414255.89 (413822.05,414689.73) | 6.44e-07 ** | 0.570 (0.560,0.579) | 0.478 (0.472,0.484) | 5.82e-50 ** |
| Birthplace (north coordinate) | 365148.17 (363601.27,366695.08) | 358619.68 (357729.78,359509.58) | 7.58e-13 ** | 0.712 (0.705,0.720) | 0.633 (0.628,0.637) | 3.91e-66 ** |
| Breastfed | 0.76 (0.76,0.77) | 0.74 (0.73,0.74) | 1.08e-14 ** | 0.318 (0.285,0.350) | 0.284 (0.267,0.300) | 0.068 |
| Educational attainment | 12.44 (12.39,12.48) | 14.22 (14.20,14.25) | 0.00e+00 ** | 0.441 (0.429,0.452) | 0.492 (0.486,0.498) | 1.23e-15 ** |
| Mother smoked during pregnancy | 0.28 (0.28,0.29) | 0.30 (0.30,0.30) | 1.31e-08 ** | 0.162 (0.134,0.190) | 0.143 (0.128,0.158) | 0.242 |
| Number of children | 1.97 (1.96,1.98) | 1.94 (1.93,1.95) | 2.93e-09 ** | 0.735 (0.729,0.741) | 0.717 (0.713,0.720) | 1.13e-06 ** |
| Number of siblings | 2.07 (2.05,2.08) | 2.00 (1.99,2.01) | 1.80e-11 ** | 0.123 (0.110,0.137) | 0.091 (0.083,0.098) | 4.07e-05 ** |
| **Health behaviors** | | | | | | |
| Drinks alcohol | 0.92 (0.92,0.93) | 0.94 (0.94,0.94) | 1.02e-43 ** | 0.568 (0.537,0.597) | 0.572 (0.553,0.590) | 0.809 |
| Drinks full-fat or semi-skimmed milk | 0.71 (0.70,0.71) | 0.71 (0.71,0.71) | 0.240 | 0.720 (0.706,0.734) | 0.756 (0.749,0.763) | 3.63e-06 ** |
| Eats processed meat >= 1x week | 0.65 (0.64,0.65) | 0.64 (0.64,0.64) | 0.005 | 0.396 (0.375,0.416) | 0.431 (0.420,0.442) | 0.003 |
| Ever smoked | 0.59 (0.59,0.60) | 0.59 (0.59,0.59) | 0.117 | 0.246 (0.225,0.268) | 0.243 (0.231,0.254) | 0.766 |
| Fruit intake | 2.96 (2.94,2.98) | 2.96 (2.95,2.97) | 0.868 | 0.249 (0.235,0.263) | 0.253 (0.245,0.261) | 0.633 |
| Moderate physical activity (days/week) | 3.76 (3.74,3.79) | 3.58 (3.56,3.59) | 5.22e-41 ** | 0.088 (0.073,0.103) | 0.122 (0.114,0.129) | 7.12e-05 ** |
| Smoked at least 100 times | 0.45 (0.44,0.45) | 0.43 (0.43,0.43) | 2.09e-11 ** | 0.371 (0.351,0.391) | 0.365 (0.354,0.377) | 0.632 |
| Smoking status (Current) | 0.08 (0.07,0.08) | 0.07 (0.07,0.07) | 0.005 | 0.477 (0.443,0.509) | 0.521 (0.503,0.539) | 0.019 |
| Smoking status (Former) | 0.38 (0.37,0.38) | 0.36 (0.36,0.36) | 2.16e-08 ** | 0.309 (0.288,0.330) | 0.315 (0.303,0.327) | 0.604 |
| Smoking status (Never) | 0.55 (0.54,0.55) | 0.57 (0.56,0.57) | 3.63e-12 ** | 0.364 (0.344,0.383) | 0.369 (0.358,0.380) | 0.656 |
| Sun protection | 0.57 (0.57,0.58) | 0.59 (0.59,0.59) | 6.40e-09 ** | 0.263 (0.241,0.285) | 0.237 (0.225,0.249) | 0.041 |
| Vegetable intake | 4.81 (4.78,4.83) | 4.71 (4.70,4.72) | 3.01e-11 ** | 0.240 (0.226,0.253) | 0.262 (0.255,0.269) | 0.004 |
| Walking (minutes/day) | 60.41 (59.57,61.24) | 58.92 (58.50,59.33) | 0.002 | 0.220 (0.203,0.236) | 0.180 (0.171,0.188) | 2.06e-05 ** |
| **Health traits** | | | | | | |
| Asthma | 0.11 (0.10,0.11) | 0.11 (0.11,0.11) | 0.022 | -0.006 (-0.045,0.032) | -0.003 (-0.024,0.018) | 0.878 |
| Basal metabolic rate | 6612.46 (6599.46,6625.45) | 6675.70 (6668.44,6682.96) | 8.41e-17 ** | 0.212 (0.199,0.225) | 0.203 (0.196,0.210) | 0.242 |
| Dental problems | 0.41 (0.40,0.41) | 0.37 (0.37,0.38) | 3.63e-32 ** | 0.161 (0.140,0.183) | 0.139 (0.127,0.151) | 0.075 |
| Diabetes | 0.06 (0.06,0.06) | 0.05 (0.05,0.05) | 1.59e-21 ** | 0.183 (0.133,0.232) | 0.203 (0.172,0.233) | 0.508 |

| | | | | | | |
|---|---|---|---|---|---|---|
| Difficulty hearing | 0.30 (0.29,0.30) | 0.27 (0.27,0.27) | 1.20e-20 ** | -0.070 (-0.095,-0.044) | -0.069 (-0.084,-0.055) | 0.976 |
| Ever had tinnitus | 0.31 (0.30,0.31) | 0.29 (0.28,0.29) | 1.35E-04 | 0.077 (0.035,0.119) | 0.061 (0.038,0.083) | 0.503 |
| Hay fever, allergic rhinitis, or eczema | 0.20 (0.20,0.20) | 0.23 (0.23,0.23) | 1.25e-41 ** | 0.022 (-0.006,0.049) | 0.010 (-0.004,0.025) | 0.485 |
| High blood pressure | 0.31 (0.31,0.32) | 0.27 (0.27,0.28) | 1.08e-49 ** | 0.084 (0.061,0.107) | 0.101 (0.088,0.115) | 0.212 |
| Long-term illness | 0.34 (0.34,0.35) | 0.30 (0.30,0.31) | 5.54e-42 ** | 0.139 (0.115,0.161) | 0.131 (0.118,0.144) | 0.586 |
| Number of illnesses | 1.94 (1.92,1.96) | 1.77 (1.76,1.78) | 7.70e-67 ** | 0.164 (0.151,0.177) | 0.156 (0.149,0.164) | 0.326 |
| Number of medications | 2.66 (2.63,2.68) | 2.33 (2.32,2.34) | 6.14e-116 ** | 0.217 (0.204,0.230) | 0.215 (0.208,0.222) | 0.846 |
| Wears Glasses | 0.93 (0.93,0.93) | 0.91 (0.91,0.91) | 9.43e-42 ** | 0.369 (0.330,0.407) | 0.382 (0.363,0.400) | 0.562 |
| **Psychological traits** | | | | | | |
| Anxious feelings | 0.58 (0.57,0.58) | 0.57 (0.56,0.57) | 4.23e-06 ** | -0.095 (-0.118,-0.072) | -0.103 (-0.115,-0.090) | 0.573 |
| Doctor-diagnosed anxiety or depression | 0.29 (0.29,0.30) | 0.31 (0.31,0.31) | 2.24e-09 ** | 0.145 (0.120,0.169) | 0.153 (0.140,0.166) | 0.545 |
| Ever depressed for at least one week | 0.45 (0.44,0.46) | 0.48 (0.48,0.49) | 6.63e-11 ** | 0.135 (0.096,0.174) | 0.141 (0.121,0.161) | 0.808 |
| Fed-up feelings | 0.38 (0.38,0.39) | 0.37 (0.37,0.38) | 0.005 | 0.153 (0.130,0.175) | 0.150 (0.138,0.163) | 0.851 |
| Guilt | 0.25 (0.25,0.25) | 0.27 (0.27,0.27) | 1.13e-13 ** | 0.085 (0.058,0.111) | 0.054 (0.040,0.068) | 0.051 |
| High strung | 0.17 (0.17,0.17) | 0.16 (0.16,0.17) | 0.017 | 0.031 (-0.001,0.063) | 0.020 (0.002,0.037) | 0.535 |
| Hurt feelings | 0.56 (0.56,0.57) | 0.55 (0.54,0.55) | 4.76e-07 ** | 0.045 (0.022,0.068) | 0.042 (0.030,0.055) | 0.849 |
| Irritability | 0.28 (0.28,0.29) | 0.29 (0.29,0.29) | 8.47E-04 | -0.032 (-0.058,-0.006) | -0.016 (-0.029,-0.002) | 0.270 |
| Loneliness | 0.12 (0.11,0.12) | 0.11 (0.11,0.12) | 0.533 | 0.175 (0.138,0.210) | 0.152 (0.133,0.172) | 0.292 |
| Miserableness | 0.40 (0.39,0.40) | 0.41 (0.41,0.41) | 2.50e-06 ** | 0.133 (0.110,0.156) | 0.102 (0.090,0.114) | 0.018 |
| Mood swings | 0.45 (0.44,0.45) | 0.43 (0.43,0.44) | 6.32e-08 ** | 0.070 (0.048,0.093) | 0.055 (0.043,0.067) | 0.249 |
| Nerves | 0.21 (0.20,0.21) | 0.20 (0.20,0.20) | 0.030 | 0.097 (0.069,0.125) | 0.074 (0.059,0.090) | 0.166 |
| Nervous feelings | 0.25 (0.24,0.25) | 0.23 (0.23,0.23) | 2.25e-10 ** | 0.062 (0.035,0.089) | 0.028 (0.013,0.043) | 0.031 |
| Neuroticism | 3.96 (3.92,4.00) | 3.96 (3.94,3.98) | 0.927 | 0.051 (0.033,0.069) | 0.040 (0.031,0.049) | 0.255 |
| Worry too long after embarrassment | 0.46 (0.46,0.47) | 0.48 (0.47,0.48) | 7.84e-07 ** | 0.024 (0.000,0.047) | 0.021 (0.008,0.033) | 0.840 |

**Supplemental table 15: Comparison between pairs that do not match on self-reported household income and those that do**

| | Trait means | | | Trait correlations | | |
|---|---|---|---|---|---|---|
| | **Excluded subset** | **Retained subset** | **P** | **Excluded subset** | **Retained subset** | **P** |
| **Anthropometric traits** | | | | | | |
| Body mass index | 27.18 (27.13,27.22) | 27.13 (27.10,27.15) | 0.078 | 0.234 (0.219,0.249) | 0.244 (0.236,0.252) | 0.255 |
| C-reactive protein (log10) | 0.12 (0.12,0.13) | 0.12 (0.12,0.13) | 0.907 | 0.135 (0.119,0.152) | 0.132 (0.123,0.141) | 0.762 |
| Cholesterol | 5.74 (5.72,5.75) | 5.69 (5.69,5.70) | 5.70e-08 ** | 0.066 (0.049,0.083) | 0.082 (0.073,0.091) | 0.092 |
| Diastolic blood pressure | 82.58 (82.46,82.69) | 82.48 (82.42,82.54) | 0.126 | 0.115 (0.099,0.131) | 0.107 (0.099,0.116) | 0.412 |
| Fluid IQ | 6.21 (6.17,6.25) | 6.23 (6.20,6.25) | 0.525 | 0.199 (0.171,0.226) | 0.228 (0.214,0.242) | 0.064 |
| Forced expiratory volume in 1 second | 2.90 (2.89,2.91) | 2.90 (2.90,2.91) | 0.849 | 0.254 (0.235,0.274) | 0.268 (0.258,0.279) | 0.228 |
| Forced vital capacity | 3.84 (3.83,3.85) | 3.84 (3.83,3.84) | 0.515 | 0.241 (0.221,0.261) | 0.264 (0.254,0.275) | 0.039 |
| Hand grip strength | 33.79 (33.67,33.92) | 33.62 (33.55,33.69) | 0.018 | 0.158 (0.143,0.174) | 0.184 (0.176,0.192) | 0.004 |
| Handedness | 0.10 (0.09,0.10) | 0.10 (0.09,0.10) | 0.249 | 0.013 (-0.035,0.061) | -0.006 (-0.032,0.020) | 0.491 |
| Heel bone mineral density | -0.32 (-0.34,-0.31) | -0.34 (-0.35,-0.33) | 0.124 | 0.038 (0.017,0.060) | 0.035 (0.023,0.047) | 0.784 |
| Height | 169.30 (169.20,169.40) | 169.26 (169.21,169.32) | 0.564 | 0.230 (0.214,0.245) | 0.258 (0.250,0.266) | 8.56E-04 |
| Hemoglobin A1c | 35.35 (35.29,35.40) | 35.29 (35.26,35.31) | 0.042 | 0.107 (0.090,0.124) | 0.137 (0.128,0.146) | 0.002 |
| High-density lipoprotein (HDL) | 1.44 (1.43,1.44) | 1.44 (1.43,1.44) | 0.621 | 0.159 (0.141,0.177) | 0.151 (0.141,0.160) | 0.437 |
| Hip circumference | 103.07 (102.98,103.16) | 102.94 (102.89,102.99) | 0.016 | 0.181 (0.166,0.197) | 0.187 (0.179,0.195) | 0.512 |
| Low-density lipoprotein (LDL) | 3.59 (3.58,3.60) | 3.56 (3.55,3.56) | 1.47e-08 ** | 0.054 (0.037,0.070) | 0.072 (0.063,0.081) | 0.063 |
| Peak expiratory flow | 382.25 (380.72,383.77) | 382.54 (381.71,383.36) | 0.740 | 0.164 (0.147,0.181) | 0.185 (0.176,0.194) | 0.035 |
| Pulse | 68.21 (68.08,68.33) | 68.04 (67.97,68.10) | 0.017 | 0.109 (0.093,0.126) | 0.115 (0.106,0.123) | 0.566 |
| Red blood cell count | 4.53 (4.53,4.54) | 4.54 (4.53,4.54) | 0.436 | 0.094 (0.078,0.111) | 0.096 (0.087,0.105) | 0.855 |
| Systolic blood pressure | 138.73 (138.53,138.94) | 138.70 (138.58,138.81) | 0.745 | 0.158 (0.141,0.174) | 0.167 (0.158,0.175) | 0.322 |
| Waist circumference | 90.38 (90.23,90.52) | 90.20 (90.12,90.27) | 0.029 | 0.259 (0.244,0.273) | 0.264 (0.256,0.272) | 0.552 |
| Waist-hip ratio | 0.87 (0.87,0.88) | 0.87 (0.87,0.87) | 0.675 | 0.179 (0.163,0.194) | 0.189 (0.181,0.197) | 0.251 |
| Weight | 78.18 (78.01,78.34) | 78.01 (77.92,78.10) | 0.086 | 0.213 (0.198,0.228) | 0.219 (0.210,0.227) | 0.536 |
| White blood cell count | 6.76 (6.74,6.78) | 6.80 (6.79,6.81) | 1.38E-04 | 0.154 (0.137,0.170) | 0.158 (0.149,0.167) | 0.626 |
| Whole body impedance | 595.09 (594.07,596.10) | 595.94 (595.40,596.48) | 0.146 | 0.125 (0.109,0.141) | 0.128 (0.119,0.136) | 0.800 |
| **Behavioral traits** | | | | | | |
| Age at first intercourse | 19.01 (18.98,19.05) | 19.06 (19.03,19.08) | 0.070 | 0.401 (0.387,0.416) | 0.416 (0.408,0.424) | 0.079 |
| Chronotype | 0.36 (0.35,0.36) | 0.36 (0.36,0.37) | 0.057 | -0.181 (-0.210,-0.153) | -0.181 (-0.196,-0.166) | 0.994 |
| Number of sexual partners | 4.43 (4.35,4.50) | 4.23 (4.19,4.27) | 3.38e-06 ** | 0.490 (0.476,0.503) | 0.502 (0.495,0.509) | 0.123 |

| | | | | | | |
|---|---|---|---|---|---|---|
| Risk taking | 0.25 (0.25,0.26) | 0.23 (0.22,0.23) | 9.55e-16 ** | 0.053 (0.021,0.084) | 0.078 (0.061,0.096) | 0.164 |
| Sleep duration | 7.23 (7.22,7.24) | 7.23 (7.23,7.24) | 0.249 | 0.101 (0.085,0.117) | 0.129 (0.121,0.138) | 0.002 |
| Time spent using computer | 0.98 (0.97,1.00) | 0.93 (0.92,0.94) | 6.45e-10 ** | 0.100 (0.084,0.116) | 0.097 (0.089,0.106) | 0.770 |
| Time spent watching TV | 2.73 (2.71,2.75) | 2.78 (2.77,2.79) | 5.91e-07 ** | 0.428 (0.415,0.441) | 0.468 (0.462,0.475) | 4.60e-08 ** |
| Water intake | 2.47 (2.45,2.49) | 2.47 (2.45,2.48) | 0.785 | 0.191 (0.176,0.206) | 0.222 (0.214,0.230) | 5.29E-04 |
| **Demographic/Family traits** | | | | | | |
| Age | 57.23 (57.15,57.31) | 57.28 (57.23,57.32) | 0.301 | 0.842 (0.837,0.846) | 0.857 (0.855,0.860) | 1.23e-09 ** |
| Birthplace (east coordinate) | 412025.10 (411092.49,412957.71) | 414888.10 (414398.11,415378.09) | 1.01e-07 ** | 0.479 (0.465,0.492) | 0.478 (0.471,0.485) | 0.924 |
| Birthplace (north coordinate) | 357868.69 (355946.75,359790.64) | 358832.84 (357829.01,359836.67) | 0.383 | 0.641 (0.631,0.651) | 0.630 (0.625,0.636) | 0.054 |
| Breastfed | 0.74 (0.74,0.75) | 0.74 (0.73,0.74) | 0.067 | 0.260 (0.225,0.295) | 0.290 (0.272,0.308) | 0.138 |
| Educational attainment | 14.27 (14.22,14.32) | 14.21 (14.18,14.24) | 0.045 | 0.448 (0.435,0.461) | 0.504 (0.498,0.510) | 4.61e-15 ** |
| Mother smoked during pregnancy | 0.30 (0.29,0.30) | 0.30 (0.30,0.30) | 0.821 | 0.145 (0.114,0.175) | 0.143 (0.126,0.159) | 0.911 |
| Number of children | 1.96 (1.94,1.97) | 1.93 (1.93,1.94) | 0.002 | 0.707 (0.699,0.715) | 0.719 (0.715,0.723) | 0.005 |
| Number of siblings | 1.99 (1.97,2.01) | 2.00 (1.99,2.01) | 0.632 | 0.089 (0.074,0.105) | 0.091 (0.083,0.100) | 0.835 |
| **Health behaviors** | | | | | | |
| Drinks alcohol | 0.94 (0.94,0.95) | 0.94 (0.94,0.94) | 0.088 | 0.515 (0.470,0.556) | 0.588 (0.567,0.608) | 0.002 |
| Drinks full-fat or semi-skimmed milk | 0.72 (0.71,0.72) | 0.71 (0.71,0.71) | 0.044 | 0.727 (0.711,0.743) | 0.764 (0.756,0.772) | 3.19e-05 ** |
| Eats processed meat >= 1x week | 0.63 (0.63,0.64) | 0.64 (0.64,0.64) | 0.126 | 0.381 (0.357,0.405) | 0.446 (0.433,0.458) | 2.37e-06 ** |
| Ever smoked | 0.59 (0.58,0.60) | 0.59 (0.59,0.59) | 0.675 | 0.239 (0.215,0.264) | 0.244 (0.230,0.257) | 0.761 |
| Fruit intake | 2.96 (2.94,2.99) | 2.96 (2.95,2.98) | 0.954 | 0.232 (0.215,0.249) | 0.259 (0.250,0.268) | 0.004 |
| Moderate physical activity (days/week) | 3.60 (3.57,3.62) | 3.57 (3.55,3.58) | 0.074 | 0.091 (0.074,0.107) | 0.131 (0.122,0.139) | 2.22e-05 ** |
| Smoked at least 100 times | 0.43 (0.43,0.44) | 0.43 (0.43,0.43) | 0.196 | 0.343 (0.319,0.366) | 0.372 (0.359,0.384) | 0.032 |
| Smoking status (Current) | 0.08 (0.07,0.08) | 0.07 (0.07,0.07) | 9.99E-05 | 0.487 (0.448,0.524) | 0.532 (0.511,0.552) | 0.038 |
| Smoking status (Former) | 0.36 (0.35,0.36) | 0.36 (0.36,0.37) | 0.166 | 0.291 (0.266,0.315) | 0.322 (0.309,0.335) | 0.028 |
| Smoking status (Never) | 0.56 (0.56,0.57) | 0.57 (0.56,0.57) | 0.463 | 0.358 (0.335,0.381) | 0.372 (0.360,0.384) | 0.304 |
| Sun protection | 0.59 (0.58,0.59) | 0.59 (0.59,0.59) | 0.252 | 0.239 (0.213,0.264) | 0.237 (0.223,0.250) | 0.902 |
| Vegetable intake | 4.71 (4.68,4.74) | 4.71 (4.70,4.73) | 0.932 | 0.226 (0.210,0.241) | 0.272 (0.264,0.280) | 1.30e-07 ** |
| Walking (minutes/day) | 58.85 (57.98,59.73) | 58.93 (58.47,59.40) | 0.871 | 0.136 (0.118,0.153) | 0.193 (0.183,0.202) | 2.32e-08 ** |
| **Health traits** | | | | | | |
| Asthma | 0.11 (0.10,0.11) | 0.11 (0.11,0.11) | 0.160 | -0.036 (-0.081,0.010) | 0.006 (-0.018,0.029) | 0.111 |
| Basal metabolic rate | 6686.31 (6670.80,6701.83) | 6672.65 (6664.44,6680.87) | 0.127 | 0.199 (0.184,0.215) | 0.204 (0.196,0.212) | 0.619 |
| Dental problems | 0.37 (0.37,0.38) | 0.37 (0.37,0.38) | 0.224 | 0.115 (0.089,0.141) | 0.146 (0.132,0.159) | 0.038 |
| Diabetes | 0.05 (0.04,0.05) | 0.05 (0.05,0.05) | 0.470 | 0.178 (0.110,0.245) | 0.209 (0.175,0.244) | 0.415 |

| | | | | | | |
|---|---|---|---|---|---|---|
| Difficulty hearing | 0.27 (0.26,0.27) | 0.27 (0.27,0.27) | 0.157 | -0.075 (-0.105,-0.044) | -0.068 (-0.084,-0.052) | 0.708 |
| Ever had tinnitus | 0.29 (0.28,0.30) | 0.29 (0.28,0.29) | 0.742 | 0.036 (-0.015,0.087) | 0.067 (0.042,0.092) | 0.283 |
| Hay fever, allergic rhinitis, or eczema | 0.22 (0.22,0.23) | 0.23 (0.23,0.24) | 1.27e-05 ** | 0.016 (-0.015,0.047) | 0.009 (-0.008,0.025) | 0.695 |
| High blood pressure | 0.27 (0.27,0.28) | 0.27 (0.27,0.28) | 0.694 | 0.081 (0.052,0.109) | 0.107 (0.092,0.122) | 0.110 |
| Long-term illness | 0.30 (0.29,0.30) | 0.31 (0.30,0.31) | 5.11E-04 | 0.129 (0.101,0.157) | 0.132 (0.117,0.146) | 0.883 |
| Number of illnesses | 1.72 (1.70,1.74) | 1.79 (1.78,1.80) | 7.22e-10 ** | 0.155 (0.139,0.170) | 0.157 (0.148,0.165) | 0.829 |
| Number of medications | 2.28 (2.25,2.31) | 2.34 (2.33,2.36) | 3.30e-05 ** | 0.208 (0.193,0.223) | 0.217 (0.209,0.225) | 0.309 |
| Wears Glasses | 0.91 (0.91,0.91) | 0.91 (0.91,0.91) | 0.580 | 0.356 (0.315,0.395) | 0.389 (0.368,0.410) | 0.143 |
| **Psychological traits** | | | | | | |
| Anxious feelings | 0.56 (0.56,0.57) | 0.57 (0.57,0.57) | 0.020 | -0.111 (-0.137,-0.085) | -0.100 (-0.114,-0.086) | 0.458 |
| Doctor-diagnosed anxiety or depression | 0.31 (0.30,0.31) | 0.31 (0.31,0.31) | 0.381 | 0.137 (0.109,0.164) | 0.158 (0.143,0.172) | 0.184 |
| Ever depressed for at least one week | 0.49 (0.48,0.50) | 0.48 (0.48,0.49) | 0.084 | 0.152 (0.107,0.197) | 0.138 (0.115,0.160) | 0.582 |
| Fed-up feelings | 0.37 (0.37,0.38) | 0.37 (0.37,0.38) | 0.811 | 0.136 (0.110,0.162) | 0.154 (0.141,0.168) | 0.225 |
| Guilt | 0.27 (0.27,0.28) | 0.27 (0.27,0.27) | 0.180 | 0.029 (-0.000,0.059) | 0.062 (0.046,0.078) | 0.062 |
| High strung | 0.16 (0.16,0.17) | 0.16 (0.16,0.17) | 0.859 | 0.001 (-0.036,0.038) | 0.025 (0.005,0.045) | 0.262 |
| Hurt feelings | 0.55 (0.55,0.56) | 0.55 (0.54,0.55) | 0.055 | 0.059 (0.033,0.085) | 0.038 (0.023,0.052) | 0.160 |
| Irritability | 0.29 (0.28,0.30) | 0.29 (0.29,0.29) | 0.918 | -0.013 (-0.042,0.016) | -0.016 (-0.032,-0.001) | 0.858 |
| Loneliness | 0.12 (0.12,0.13) | 0.11 (0.11,0.11) | 1.07e-05 ** | 0.136 (0.095,0.177) | 0.157 (0.135,0.180) | 0.376 |
| Miserableness | 0.41 (0.41,0.42) | 0.41 (0.41,0.41) | 0.753 | 0.081 (0.055,0.107) | 0.108 (0.094,0.122) | 0.079 |
| Mood swings | 0.44 (0.43,0.44) | 0.43 (0.43,0.44) | 0.067 | 0.055 (0.029,0.081) | 0.055 (0.042,0.069) | 0.982 |
| Nerves | 0.20 (0.20,0.21) | 0.20 (0.20,0.20) | 0.961 | 0.061 (0.028,0.094) | 0.078 (0.060,0.095) | 0.385 |
| Nervous feelings | 0.23 (0.23,0.24) | 0.23 (0.23,0.23) | 0.439 | -0.005 (-0.037,0.027) | 0.037 (0.020,0.054) | 0.021 |
| Neuroticism | 3.97 (3.93,4.02) | 3.95 (3.93,3.98) | 0.452 | 0.021 (0.002,0.040) | 0.045 (0.035,0.055) | 0.032 |
| Worry too long after embarrassment | 0.48 (0.47,0.48) | 0.48 (0.47,0.48) | 0.655 | 0.010 (-0.016,0.036) | 0.024 (0.010,0.038) | 0.355 |

**Supplemental table 16: Comparison between pairs that differ by 5+ years for length of time at current address and those that do not**

| | Trait means | | | Trait correlations | | |
|---|---|---|---|---|---|---|
| | **Excluded subset** | **Retained subset** | **P** | **Excluded subset** | **Retained subset** | **P** |
| **Anthropometric traits** | | | | | | |
| Body mass index | 27.47 (27.39,27.54) | 27.19 (27.17,27.21) | 4.90e-12 ** | 0.238 (0.215,0.262) | 0.239 (0.233,0.246) | 0.937 |
| C-reactive protein (log10) | 0.17 (0.16,0.17) | 0.13 (0.13,0.13) | 1.67e-14 ** | 0.134 (0.108,0.160) | 0.131 (0.124,0.138) | 0.828 |
| Cholesterol | 5.68 (5.66,5.70) | 5.70 (5.70,5.71) | 0.055 | 0.091 (0.065,0.117) | 0.078 (0.071,0.086) | 0.375 |
| Diastolic blood pressure | 82.61 (82.43,82.78) | 82.54 (82.49,82.59) | 0.479 | 0.112 (0.086,0.137) | 0.101 (0.094,0.108) | 0.418 |
| Fluid IQ | 5.92 (5.85,5.98) | 6.14 (6.12,6.15) | 3.69e-10 ** | 0.188 (0.146,0.230) | 0.226 (0.214,0.237) | 0.094 |
| Forced expiratory volume in 1 second | 2.82 (2.80,2.83) | 2.88 (2.88,2.89) | 1.97e-12 ** | 0.186 (0.153,0.219) | 0.272 (0.264,0.281) | 3.97e-07 ** |
| Forced vital capacity | 3.74 (3.72,3.76) | 3.81 (3.80,3.81) | 9.09e-09 ** | 0.200 (0.168,0.233) | 0.263 (0.255,0.272) | 2.12E-04 |
| Hand grip strength | 33.09 (32.89,33.28) | 33.43 (33.38,33.49) | 7.84E-04 | 0.161 (0.137,0.186) | 0.180 (0.173,0.186) | 0.159 |
| Handedness | 0.10 (0.09,0.10) | 0.10 (0.09,0.10) | 0.324 | -0.089 (-0.167,-0.011) | -0.001 (-0.022,0.020) | 0.033 |
| Heel bone mineral density | -0.39 (-0.42,-0.37) | -0.35 (-0.35,-0.34) | 7.13E-04 | 0.032 (-0.002,0.066) | 0.039 (0.030,0.048) | 0.695 |
| Height | 168.87 (168.71,169.03) | 169.07 (169.03,169.12) | 0.019 | 0.261 (0.238,0.284) | 0.258 (0.252,0.265) | 0.827 |
| Hemoglobin A1c | 35.64 (35.56,35.73) | 35.42 (35.40,35.44) | 3.25e-07 ** | 0.143 (0.116,0.169) | 0.131 (0.124,0.139) | 0.422 |
| High-density lipoprotein (HDL) | 1.42 (1.41,1.43) | 1.43 (1.43,1.44) | 2.37E-04 | 0.159 (0.131,0.186) | 0.154 (0.146,0.161) | 0.742 |
| Hip circumference | 103.26 (103.11,103.40) | 103.00 (102.96,103.04) | 7.73E-04 | 0.182 (0.158,0.206) | 0.185 (0.178,0.191) | 0.832 |
| Low-density lipoprotein (LDL) | 3.55 (3.53,3.57) | 3.57 (3.56,3.57) | 0.068 | 0.079 (0.052,0.105) | 0.067 (0.060,0.074) | 0.399 |
| Peak expiratory flow | 369.38 (366.99,371.77) | 378.84 (378.18,379.50) | 8.44e-14 ** | 0.167 (0.140,0.194) | 0.183 (0.176,0.190) | 0.255 |
| Pulse | 68.38 (68.19,68.58) | 68.14 (68.08,68.19) | 0.017 | 0.114 (0.088,0.140) | 0.112 (0.105,0.119) | 0.885 |
| Red blood cell count | 4.53 (4.52,4.54) | 4.54 (4.54,4.54) | 0.131 | 0.092 (0.066,0.117) | 0.096 (0.089,0.103) | 0.724 |
| Systolic blood pressure | 139.22 (138.89,139.54) | 139.24 (139.15,139.33) | 0.909 | 0.140 (0.115,0.166) | 0.163 (0.156,0.170) | 0.095 |
| Waist circumference | 91.23 (91.00,91.46) | 90.36 (90.30,90.43) | 1.04e-12 ** | 0.262 (0.239,0.285) | 0.260 (0.254,0.267) | 0.892 |
| Waist-hip ratio | 0.88 (0.88,0.88) | 0.88 (0.87,0.88) | 1.14e-13 ** | 0.197 (0.173,0.221) | 0.186 (0.179,0.193) | 0.385 |
| Weight | 78.53 (78.27,78.80) | 77.99 (77.92,78.06) | 1.09E-04 | 0.219 (0.195,0.243) | 0.216 (0.209,0.222) | 0.809 |
| White blood cell count | 6.90 (6.87,6.93) | 6.80 (6.79,6.81) | 2.17e-10 ** | 0.169 (0.144,0.194) | 0.154 (0.147,0.161) | 0.247 |
| Whole body impedance | 593.29 (591.70,594.88) | 595.43 (594.99,595.86) | 0.011 | 0.115 (0.090,0.140) | 0.137 (0.130,0.144) | 0.095 |
| **Behavioral traits** | | | | | | |
| Age at first intercourse | 18.67 (18.60,18.73) | 19.09 (19.08,19.11) | 3.52e-37 ** | 0.265 (0.238,0.291) | 0.427 (0.421,0.433) | 3.91e-35 ** |
| Chronotype | 0.36 (0.35,0.37) | 0.36 (0.36,0.36) | 0.876 | -0.137 (-0.182,-0.091) | -0.179 (-0.192,-0.167) | 0.072 |
| Number of sexual partners | 5.83 (5.69,5.98) | 3.99 (3.96,4.02) | 1.18e-126 ** | 0.469 (0.445,0.492) | 0.493 (0.487,0.499) | 0.049 |

| | | | | | | |
|---|---|---|---|---|---|---|
| Risk taking | 0.24 (0.24,0.25) | 0.23 (0.22,0.23) | 1.83e-05 ** | 0.039 (-0.010,0.089) | 0.071 (0.057,0.085) | 0.232 |
| Sleep duration | 7.26 (7.24,7.27) | 7.24 (7.23,7.24) | 0.061 | 0.089 (0.064,0.114) | 0.122 (0.115,0.129) | 0.013 |
| Time spent using computer | 0.94 (0.92,0.97) | 0.92 (0.91,0.92) | 0.029 | 0.131 (0.106,0.156) | 0.098 (0.091,0.105) | 0.012 |
| Time spent watching TV | 2.89 (2.87,2.92) | 2.83 (2.82,2.83) | 6.19e-06 ** | 0.426 (0.405,0.446) | 0.448 (0.443,0.454) | 0.040 |
| Water intake | 2.45 (2.42,2.49) | 2.46 (2.45,2.47) | 0.877 | 0.189 (0.164,0.213) | 0.216 (0.210,0.223) | 0.031 |
| **Demographic/Family traits** | | | | | | |
| Age | 58.27 (58.15,58.40) | 57.81 (57.78,57.85) | 4.42e-12 ** | 0.699 (0.686,0.712) | 0.866 (0.864,0.868) | 1.53e-259 ** |
| Birthplace (east coordinate) | 412786.56 (411397.11,414176.02) | 413816.05 (413427.33,414204.78) | 0.162 | 0.459 (0.437,0.480) | 0.500 (0.494,0.505) | 1.68E-04 |
| Birthplace (north coordinate) | 353934.54 (351078.42,356790.67) | 360612.55 (359809.87,361415.22) | 1.03e-05 ** | 0.636 (0.620,0.652) | 0.651 (0.647,0.655) | 0.074 |
| Breastfed | 0.75 (0.74,0.76) | 0.74 (0.74,0.74) | 0.365 | 0.324 (0.268,0.377) | 0.288 (0.273,0.303) | 0.224 |
| Educational attainment | 13.28 (13.19,13.36) | 13.87 (13.85,13.90) | 3.03e-38 ** | 0.435 (0.415,0.456) | 0.496 (0.491,0.501) | 6.08e-09 ** |
| Mother smoked during pregnancy | 0.31 (0.30,0.32) | 0.29 (0.29,0.30) | 0.017 | 0.144 (0.094,0.192) | 0.148 (0.134,0.161) | 0.877 |
| Number of children | 1.70 (1.68,1.72) | 1.97 (1.96,1.97) | 3.22e-130 ** | 0.371 (0.350,0.393) | 0.752 (0.749,0.755) | 0.00e+00 ** |
| Number of siblings | 2.02 (1.99,2.05) | 2.01 (2.00,2.02) | 0.770 | 0.062 (0.037,0.087) | 0.101 (0.095,0.108) | 0.002 |
| **Health behaviors** | | | | | | |
| Drinks alcohol | 0.94 (0.93,0.94) | 0.94 (0.94,0.94) | 0.974 | 0.427 (0.354,0.496) | 0.582 (0.565,0.598) | 6.72e-06 ** |
| Drinks full-fat or semi-skimmed milk | 0.71 (0.70,0.72) | 0.71 (0.71,0.71) | 0.936 | 0.735 (0.710,0.759) | 0.749 (0.743,0.756) | 0.277 |
| Eats processed meat >= 1x week | 0.63 (0.62,0.64) | 0.64 (0.64,0.64) | 0.010 | 0.395 (0.357,0.432) | 0.426 (0.416,0.436) | 0.121 |
| Ever smoked | 0.61 (0.60,0.62) | 0.59 (0.59,0.59) | 8.71e-08 ** | 0.174 (0.135,0.214) | 0.249 (0.238,0.259) | 3.25E-04 |
| Fruit intake | 2.93 (2.89,2.97) | 2.96 (2.95,2.98) | 0.146 | 0.243 (0.217,0.268) | 0.254 (0.247,0.261) | 0.421 |
| Moderate physical activity (days/week) | 3.71 (3.66,3.75) | 3.61 (3.60,3.62) | 2.42e-05 ** | 0.087 (0.061,0.113) | 0.117 (0.110,0.124) | 0.028 |
| Smoked at least 100 times | 0.48 (0.47,0.49) | 0.43 (0.43,0.43) | 1.01e-22 ** | 0.283 (0.245,0.320) | 0.373 (0.362,0.383) | 4.34e-06 ** |
| Smoking status (Current) | 0.10 (0.10,0.11) | 0.07 (0.07,0.07) | 4.03e-34 ** | 0.533 (0.482,0.580) | 0.505 (0.489,0.522) | 0.306 |
| Smoking status (Former) | 0.37 (0.37,0.38) | 0.37 (0.36,0.37) | 0.045 | 0.211 (0.171,0.250) | 0.321 (0.311,0.331) | 6.20e-08 ** |
| Smoking status (Never) | 0.52 (0.51,0.53) | 0.56 (0.56,0.57) | 9.26e-21 ** | 0.285 (0.247,0.322) | 0.374 (0.364,0.384) | 3.42e-06 ** |
| Sun protection | 0.58 (0.57,0.59) | 0.59 (0.59,0.59) | 0.016 | 0.210 (0.170,0.250) | 0.246 (0.235,0.257) | 0.093 |
| Vegetable intake | 4.87 (4.82,4.91) | 4.72 (4.71,4.74) | 2.65e-09 ** | 0.241 (0.217,0.265) | 0.258 (0.251,0.264) | 0.196 |
| Walking (minutes/day) | 63.30 (61.78,64.81) | 58.91 (58.53,59.29) | 3.84e-08 ** | 0.185 (0.157,0.214) | 0.188 (0.180,0.196) | 0.860 |
| **Health traits** | | | | | | |
| Asthma | 0.12 (0.11,0.12) | 0.11 (0.11,0.11) | 6.55E-04 | 0.049 (-0.017,0.113) | -0.009 (-0.028,0.010) | 0.095 |
| Basal metabolic rate | 6666.78 (6642.57,6690.99) | 6660.65 (6654.07,6667.23) | 0.632 | 0.197 (0.173,0.222) | 0.207 (0.200,0.213) | 0.471 |
| Dental problems | 0.41 (0.40,0.42) | 0.38 (0.38,0.38) | 3.21e-14 ** | 0.151 (0.112,0.190) | 0.144 (0.133,0.155) | 0.731 |
| Diabetes | 0.06 (0.05,0.06) | 0.05 (0.05,0.05) | 7.40e-05 ** | 0.173 (0.078,0.264) | 0.201 (0.174,0.228) | 0.562 |

| | | | | | | |
|---|---|---|---|---|---|---|
| Difficulty hearing | 0.29 (0.28,0.29) | 0.28 (0.27,0.28) | 0.022 | -0.051 (-0.098,-0.004) | -0.070 (-0.083,-0.057) | 0.453 |
| Ever had tinnitus | 0.30 (0.29,0.32) | 0.29 (0.29,0.29) | 0.124 | 0.009 (-0.067,0.084) | 0.069 (0.049,0.090) | 0.128 |
| Hay fever, allergic rhinitis, or eczema | 0.22 (0.22,0.23) | 0.22 (0.22,0.23) | 0.833 | 0.038 (-0.010,0.086) | 0.013 (-0.001,0.026) | 0.322 |
| High blood pressure | 0.30 (0.30,0.31) | 0.28 (0.28,0.28) | 2.84e-08 ** | 0.094 (0.051,0.137) | 0.099 (0.087,0.111) | 0.819 |
| Long-term illness | 0.34 (0.33,0.35) | 0.31 (0.31,0.31) | 2.64e-12 ** | 0.145 (0.103,0.186) | 0.134 (0.122,0.145) | 0.619 |
| Number of illnesses | 1.92 (1.89,1.96) | 1.80 (1.79,1.81) | 7.58e-14 ** | 0.145 (0.120,0.169) | 0.160 (0.154,0.167) | 0.231 |
| Number of medications | 2.60 (2.55,2.65) | 2.39 (2.38,2.40) | 4.63e-18 ** | 0.209 (0.185,0.232) | 0.218 (0.212,0.225) | 0.447 |
| Wears Glasses | 0.92 (0.91,0.92) | 0.92 (0.91,0.92) | 0.388 | 0.260 (0.188,0.328) | 0.389 (0.372,0.406) | 2.52E-04 |
| **Psychological traits** | | | | | | |
| Anxious feelings | 0.57 (0.56,0.58) | 0.57 (0.57,0.57) | 0.919 | -0.059 (-0.100,-0.017) | -0.104 (-0.116,-0.093) | 0.036 |
| Doctor-diagnosed anxiety or depression | 0.35 (0.34,0.36) | 0.30 (0.30,0.31) | 2.33e-23 ** | 0.145 (0.103,0.186) | 0.152 (0.140,0.164) | 0.736 |
| Ever depressed for at least one week | 0.56 (0.54,0.57) | 0.47 (0.47,0.47) | 4.35e-26 ** | 0.142 (0.074,0.210) | 0.138 (0.119,0.156) | 0.892 |
| Fed-up feelings | 0.38 (0.37,0.39) | 0.38 (0.37,0.38) | 0.097 | 0.162 (0.121,0.202) | 0.151 (0.139,0.162) | 0.600 |
| Guilt | 0.27 (0.26,0.28) | 0.26 (0.26,0.27) | 0.112 | 0.054 (0.007,0.100) | 0.061 (0.048,0.074) | 0.755 |
| High strung | 0.17 (0.17,0.18) | 0.16 (0.16,0.17) | 0.017 | 0.017 (-0.039,0.074) | 0.022 (0.007,0.038) | 0.871 |
| Hurt feelings | 0.57 (0.56,0.58) | 0.55 (0.55,0.55) | 2.50e-07 ** | 0.035 (-0.007,0.077) | 0.043 (0.032,0.055) | 0.724 |
| Irritability | 0.29 (0.28,0.30) | 0.29 (0.29,0.29) | 0.831 | -0.025 (-0.071,0.021) | -0.019 (-0.031,-0.006) | 0.805 |
| Loneliness | 0.12 (0.11,0.12) | 0.11 (0.11,0.12) | 0.403 | 0.127 (0.061,0.192) | 0.160 (0.142,0.178) | 0.345 |
| Miserableness | 0.42 (0.41,0.43) | 0.41 (0.40,0.41) | 0.023 | 0.134 (0.094,0.175) | 0.107 (0.096,0.118) | 0.202 |
| Mood swings | 0.45 (0.44,0.46) | 0.44 (0.43,0.44) | 0.022 | 0.101 (0.061,0.141) | 0.055 (0.044,0.066) | 0.033 |
| Nerves | 0.21 (0.21,0.22) | 0.20 (0.20,0.20) | 0.007 | 0.109 (0.058,0.158) | 0.076 (0.062,0.090) | 0.222 |
| Nervous feelings | 0.24 (0.23,0.25) | 0.24 (0.23,0.24) | 0.423 | 0.019 (-0.031,0.068) | 0.037 (0.023,0.050) | 0.486 |
| Neuroticism | 4.02 (3.95,4.09) | 3.95 (3.93,3.97) | 0.066 | 0.085 (0.054,0.115) | 0.038 (0.030,0.047) | 0.004 |
| Worry too long after embarrassment | 0.47 (0.46,0.48) | 0.47 (0.47,0.48) | 0.236 | 0.030 (-0.012,0.071) | 0.021 (0.009,0.032) | 0.678 |

**Supplemental table 17: Comparison between pairs with an age difference of >= 10 years to pairs with an age difference of < 10 years**

| | Trait means | | | Trait correlations | | |
|---|---|---|---|---|---|---|
| | **Excluded subset** | **Retained subset** | **P** | **Excluded subset** | **Retained subset** | **P** |
| **Anthropometric traits** | | | | | | |
| Body mass index | 27.11 (27.01,27.20) | 27.21 (27.19,27.23) | 0.038 | 0.214 (0.183,0.244) | 0.241 (0.234,0.247) | 0.088 |
| C-reactive protein (log10) | 0.13 (0.12,0.15) | 0.13 (0.13,0.14) | 0.935 | 0.133 (0.100,0.166) | 0.132 (0.125,0.139) | 0.939 |
| Cholesterol | 5.58 (5.56,5.61) | 5.71 (5.70,5.71) | 4.20e-20 ** | 0.059 (0.026,0.093) | 0.080 (0.073,0.087) | 0.240 |
| Diastolic blood pressure | 82.22 (82.00,82.45) | 82.56 (82.51,82.61) | 0.004 | 0.072 (0.040,0.104) | 0.103 (0.096,0.110) | 0.069 |
| Fluid IQ | 6.02 (5.94,6.10) | 6.12 (6.11,6.14) | 0.017 | 0.205 (0.153,0.256) | 0.225 (0.214,0.236) | 0.458 |
| Forced expiratory volume in 1 second | 2.90 (2.88,2.92) | 2.87 (2.87,2.88) | 0.016 | 0.077 (0.034,0.120) | 0.278 (0.269,0.286) | 5.99e-20 ** |
| Forced vital capacity | 3.84 (3.81,3.86) | 3.80 (3.80,3.81) | 0.012 | 0.110 (0.066,0.153) | 0.268 (0.260,0.276) | 4.32e-13 ** |
| Hand grip strength | 33.69 (33.46,33.93) | 33.39 (33.34,33.45) | 0.015 | 0.112 (0.080,0.143) | 0.183 (0.176,0.189) | 1.24e-05 ** |
| Handedness | 0.09 (0.09,0.10) | 0.10 (0.09,0.10) | 0.878 | -0.054 (-0.152,0.045) | -0.005 (-0.026,0.015) | 0.342 |
| Heel bone mineral density | -0.30 (-0.34,-0.27) | -0.35 (-0.36,-0.35) | 0.007 | 0.041 (-0.002,0.085) | 0.039 (0.030,0.049) | 0.930 |
| Height | 169.19 (168.99,169.39) | 169.05 (169.00,169.09) | 0.177 | 0.182 (0.151,0.212) | 0.263 (0.257,0.269) | 2.55e-07 ** |
| Hemoglobin A1c | 35.19 (35.09,35.30) | 35.45 (35.43,35.47) | 3.43e-06 ** | 0.104 (0.070,0.137) | 0.135 (0.127,0.142) | 0.076 |
| High-density lipoprotein (HDL) | 1.42 (1.41,1.43) | 1.43 (1.43,1.44) | 0.003 | 0.134 (0.098,0.170) | 0.155 (0.148,0.163) | 0.262 |
| Hip circumference | 102.75 (102.57,102.94) | 103.03 (102.99,103.07) | 0.004 | 0.159 (0.127,0.190) | 0.186 (0.179,0.192) | 0.095 |
| Low-density lipoprotein (LDL) | 3.48 (3.46,3.50) | 3.57 (3.56,3.57) | 6.80e-17 ** | 0.054 (0.021,0.088) | 0.068 (0.061,0.075) | 0.425 |
| Peak expiratory flow | 380.58 (377.70,383.46) | 378.00 (377.35,378.66) | 0.087 | 0.099 (0.064,0.133) | 0.187 (0.180,0.194) | 6.37e-07 ** |
| Pulse | 68.26 (68.02,68.50) | 68.15 (68.10,68.21) | 0.397 | 0.110 (0.077,0.142) | 0.113 (0.106,0.119) | 0.858 |
| Red blood cell count | 4.52 (4.51,4.53) | 4.54 (4.54,4.54) | 5.33e-05 ** | 0.117 (0.085,0.150) | 0.095 (0.088,0.102) | 0.184 |
| Systolic blood pressure | 136.81 (136.40,137.23) | 139.35 (139.26,139.44) | 1.08e-31 ** | 0.069 (0.036,0.101) | 0.166 (0.159,0.172) | 8.28e-09 ** |
| Waist circumference | 90.03 (89.74,90.33) | 90.45 (90.39,90.51) | 0.007 | 0.217 (0.187,0.247) | 0.263 (0.257,0.269) | 0.004 |
| Waist-hip ratio | 0.87 (0.87,0.88) | 0.88 (0.88,0.88) | 0.031 | 0.136 (0.105,0.167) | 0.190 (0.183,0.196) | 8.29E-04 |
| Weight | 77.85 (77.51,78.19) | 78.04 (77.97,78.11) | 0.289 | 0.181 (0.150,0.212) | 0.218 (0.211,0.224) | 0.023 |
| White blood cell count | 6.90 (6.86,6.94) | 6.80 (6.80,6.81) | 2.73e-06 ** | 0.177 (0.144,0.208) | 0.154 (0.147,0.161) | 0.177 |
| Whole body impedance | 596.12 (594.08,598.15) | 595.24 (594.81,595.67) | 0.408 | 0.104 (0.072,0.136) | 0.137 (0.130,0.144) | 0.047 |
| **Behavioral traits** | | | | | | |
| Age at first intercourse | 18.38 (18.30,18.46) | 19.09 (19.08,19.11) | 9.21e-62 ** | 0.178 (0.143,0.212) | 0.427 (0.421,0.434) | 1.67e-50 ** |
| Chronotype | 0.38 (0.37,0.39) | 0.36 (0.36,0.36) | 0.001 | -0.090 (-0.147,-0.033) | -0.180 (-0.192,-0.168) | 0.002 |
| Number of sexual partners | 6.88 (6.67,7.09) | 3.99 (3.96,4.02) | 5.07e-150 ** | 0.276 (0.241,0.311) | 0.498 (0.492,0.504) | 3.38e-40 ** |

| | | | | | | |
|---|---|---|---|---|---|---|
| Risk taking | 0.29 (0.28,0.30) | 0.22 (0.22,0.23) | 3.15e-31 ** | 0.080 (0.022,0.138) | 0.066 (0.052,0.080) | 0.640 |
| Sleep duration | 7.21 (7.19,7.23) | 7.24 (7.24,7.25) | 0.011 | 0.102 (0.071,0.134) | 0.120 (0.114,0.127) | 0.272 |
| Time spent using computer | 1.01 (0.98,1.04) | 0.91 (0.91,0.92) | 3.14e-09 ** | 0.107 (0.075,0.138) | 0.101 (0.094,0.107) | 0.695 |
| Time spent watching TV | 2.71 (2.68,2.75) | 2.84 (2.83,2.84) | 3.84e-12 ** | 0.428 (0.402,0.454) | 0.447 (0.442,0.453) | 0.146 |
| Water intake | 2.66 (2.61,2.71) | 2.45 (2.44,2.46) | 7.78e-17 ** | 0.216 (0.185,0.246) | 0.214 (0.207,0.220) | 0.896 |
| **Demographic/Family traits** | | | | | | |
| Age | 55.72 (55.54,55.90) | 57.94 (57.91,57.98) | 3.86e-124 ** | -0.068 (-0.099,-0.037) | 0.911 (0.910,0.912) | 0.00e+00 ** |
| Birthplace (east coordinate) | 419808.65 (417936.14,421681.15) | 413497.36 (413115.83,413878.90) | 1.03e-10 ** | 0.377 (0.346,0.406) | 0.502 (0.497,0.508) | 1.59e-17 ** |
| Birthplace (north coordinate) | 345553.55 (341800.38,349306.72) | 360756.73 (359968.10,361545.37) | 9.03e-15 ** | 0.548 (0.523,0.572) | 0.654 (0.650,0.658) | 7.92e-20 ** |
| Breastfed | 0.73 (0.72,0.75) | 0.74 (0.74,0.74) | 0.179 | 0.216 (0.144,0.286) | 0.295 (0.280,0.309) | 0.032 |
| Educational attainment | 13.95 (13.84,14.06) | 13.82 (13.80,13.84) | 0.020 | 0.384 (0.356,0.410) | 0.498 (0.492,0.503) | 1.12e-17 ** |
| Mother smoked during pregnancy | 0.30 (0.29,0.31) | 0.30 (0.29,0.30) | 0.469 | 0.078 (0.015,0.140) | 0.151 (0.137,0.164) | 0.025 |
| Number of children | 1.66 (1.63,1.69) | 1.96 (1.96,1.97) | 3.70e-85 ** | 0.285 (0.255,0.314) | 0.751 (0.748,0.754) | 0.00e+00 ** |
| Number of siblings | 2.14 (2.10,2.18) | 2.01 (2.00,2.02) | 5.45e-10 ** | 0.078 (0.046,0.109) | 0.100 (0.093,0.106) | 0.182 |
| **Health behaviors** | | | | | | |
| Drinks alcohol | 0.92 (0.92,0.93) | 0.94 (0.94,0.94) | 1.35e-07 ** | 0.639 (0.574,0.696) | 0.568 (0.551,0.584) | 0.036 |
| Drinks full-fat or semi-skimmed milk | 0.70 (0.69,0.71) | 0.71 (0.71,0.71) | 0.032 | 0.737 (0.704,0.767) | 0.749 (0.742,0.755) | 0.465 |
| Eats processed meat >= 1x week | 0.61 (0.60,0.62) | 0.64 (0.64,0.64) | 1.27e-07 ** | 0.400 (0.353,0.446) | 0.424 (0.414,0.434) | 0.315 |
| Ever smoked | 0.60 (0.59,0.61) | 0.59 (0.59,0.59) | 0.035 | 0.206 (0.156,0.256) | 0.245 (0.235,0.255) | 0.135 |
| Fruit intake | 2.89 (2.84,2.94) | 2.96 (2.95,2.98) | 0.003 | 0.268 (0.235,0.300) | 0.252 (0.245,0.259) | 0.343 |
| Moderate physical activity (days/week) | 3.55 (3.49,3.60) | 3.62 (3.61,3.63) | 0.010 | 0.108 (0.075,0.140) | 0.116 (0.109,0.123) | 0.627 |
| Smoked at least 100 times | 0.46 (0.45,0.47) | 0.43 (0.43,0.44) | 3.03e-06 ** | 0.281 (0.231,0.329) | 0.371 (0.361,0.381) | 2.57E-04 |
| Smoking status (Current) | 0.10 (0.10,0.11) | 0.07 (0.07,0.07) | 5.61e-18 ** | 0.585 (0.524,0.640) | 0.505 (0.488,0.521) | 0.014 |
| Smoking status (Former) | 0.36 (0.35,0.37) | 0.37 (0.36,0.37) | 0.282 | 0.195 (0.144,0.246) | 0.319 (0.308,0.329) | 2.02e-06 ** |
| Smoking status (Never) | 0.54 (0.53,0.55) | 0.56 (0.56,0.56) | 2.87e-05 ** | 0.272 (0.223,0.319) | 0.372 (0.362,0.382) | 3.65e-05 ** |
| Sun protection | 0.56 (0.55,0.57) | 0.59 (0.59,0.59) | 1.24e-06 ** | 0.270 (0.220,0.318) | 0.242 (0.231,0.252) | 0.280 |
| Vegetable intake | 4.76 (4.70,4.81) | 4.73 (4.72,4.74) | 0.415 | 0.254 (0.223,0.284) | 0.257 (0.250,0.263) | 0.864 |
| Walking (minutes/day) | 61.09 (59.26,62.93) | 59.13 (58.75,59.51) | 0.040 | 0.187 (0.152,0.223) | 0.188 (0.181,0.196) | 0.972 |
| **Health traits** | | | | | | |
| Asthma | 0.11 (0.11,0.12) | 0.11 (0.11,0.11) | 0.144 | 0.014 (-0.071,0.099) | -0.005 (-0.023,0.014) | 0.683 |
| Basal metabolic rate | 6662.71 (6633.07,6692.34) | 6660.94 (6654.45,6667.43) | 0.909 | 0.147 (0.116,0.179) | 0.209 (0.202,0.216) | 1.66E-04 |
| Dental problems | 0.40 (0.39,0.41) | 0.38 (0.38,0.38) | 8.64E-04 | 0.118 (0.067,0.167) | 0.147 (0.136,0.157) | 0.267 |
| Diabetes | 0.05 (0.05,0.06) | 0.05 (0.05,0.05) | 0.298 | 0.197 (0.068,0.319) | 0.200 (0.173,0.226) | 0.966 |

| | | | | | | |
|---|---|---|---|---|---|---|
| Difficulty hearing | 0.26 (0.25,0.27) | 0.28 (0.27,0.28) | 0.002 | -0.084 (-0.147,-0.021) | -0.068 (-0.080,-0.055) | 0.611 |
| Ever had tinnitus | 0.29 (0.27,0.31) | 0.29 (0.29,0.30) | 0.801 | 0.040 (-0.054,0.133) | 0.066 (0.046,0.086) | 0.589 |
| Hay fever, allergic rhinitis, or eczema | 0.23 (0.22,0.24) | 0.22 (0.22,0.23) | 0.070 | -0.019 (-0.080,0.042) | 0.016 (0.003,0.029) | 0.268 |
| High blood pressure | 0.26 (0.25,0.27) | 0.28 (0.28,0.29) | 2.43E-04 | 0.040 (-0.019,0.098) | 0.102 (0.090,0.114) | 0.040 |
| Long-term illness | 0.32 (0.31,0.33) | 0.31 (0.31,0.32) | 0.193 | 0.159 (0.105,0.213) | 0.134 (0.122,0.145) | 0.361 |
| Number of illnesses | 1.77 (1.73,1.81) | 1.81 (1.81,1.82) | 0.033 | 0.098 (0.067,0.129) | 0.163 (0.156,0.169) | 5.89e-05 ** |
| Number of medications | 2.34 (2.29,2.40) | 2.41 (2.40,2.42) | 0.026 | 0.146 (0.115,0.177) | 0.222 (0.215,0.228) | 2.09e-06 ** |
| Wears Glasses | 0.89 (0.88,0.89) | 0.92 (0.92,0.92) | 1.01e-16 ** | 0.025 (-0.062,0.112) | 0.401 (0.384,0.417) | 2.93e-18 ** |
| **Psychological traits** | | | | | | |
| Anxious feelings | 0.55 (0.54,0.57) | 0.57 (0.57,0.57) | 0.006 | -0.054 (-0.105,-0.002) | -0.103 (-0.114,-0.092) | 0.066 |
| Doctor-diagnosed anxiety or depression | 0.34 (0.33,0.35) | 0.30 (0.30,0.31) | 4.26e-11 ** | 0.156 (0.103,0.208) | 0.151 (0.139,0.163) | 0.870 |
| Ever depressed for at least one week | 0.53 (0.51,0.55) | 0.47 (0.47,0.48) | 3.16e-08 ** | 0.188 (0.105,0.269) | 0.138 (0.119,0.156) | 0.245 |
| Fed-up feelings | 0.41 (0.39,0.42) | 0.37 (0.37,0.38) | 4.96e-08 ** | 0.093 (0.041,0.144) | 0.153 (0.142,0.164) | 0.023 |
| Guilt | 0.28 (0.27,0.29) | 0.26 (0.26,0.27) | 0.002 | 0.028 (-0.030,0.087) | 0.063 (0.050,0.076) | 0.257 |
| High strung | 0.18 (0.17,0.19) | 0.16 (0.16,0.17) | 0.006 | -0.001 (-0.072,0.069) | 0.023 (0.008,0.039) | 0.504 |
| Hurt feelings | 0.57 (0.55,0.58) | 0.55 (0.55,0.55) | 0.009 | 0.040 (-0.012,0.092) | 0.043 (0.032,0.054) | 0.921 |
| Irritability | 0.31 (0.30,0.32) | 0.29 (0.29,0.29) | 7.36e-05 ** | -0.013 (-0.069,0.044) | -0.019 (-0.032,-0.007) | 0.816 |
| Loneliness | 0.14 (0.13,0.15) | 0.11 (0.11,0.12) | 2.54e-09 ** | 0.157 (0.080,0.232) | 0.157 (0.139,0.174) | 0.989 |
| Miserableness | 0.44 (0.43,0.45) | 0.41 (0.40,0.41) | 1.82e-09 ** | 0.122 (0.071,0.173) | 0.108 (0.097,0.119) | 0.602 |
| Mood swings | 0.47 (0.45,0.48) | 0.44 (0.43,0.44) | 5.48e-07 ** | 0.077 (0.026,0.128) | 0.058 (0.047,0.069) | 0.475 |
| Nerves | 0.21 (0.20,0.22) | 0.20 (0.20,0.20) | 0.017 | 0.137 (0.074,0.199) | 0.076 (0.062,0.090) | 0.064 |
| Nervous feelings | 0.23 (0.22,0.24) | 0.24 (0.23,0.24) | 0.254 | 0.093 (0.030,0.155) | 0.033 (0.020,0.046) | 0.067 |
| Neuroticism | 4.08 (3.99,4.17) | 3.95 (3.93,3.97) | 0.005 | 0.029 (-0.009,0.068) | 0.042 (0.034,0.050) | 0.524 |
| Worry too long after embarrassment | 0.47 (0.46,0.48) | 0.47 (0.47,0.48) | 0.300 | 0.024 (-0.028,0.077) | 0.021 (0.010,0.033) | 0.911 |

**Supplemental Table 18: Trait correlations and bootstrap confidence intervals for all samples, with p-values for comparisons to the full UKB-colocated sample**

| | UKB-colocated sample | Geographically-colocated sample | | UKB-colocated sample + income match criterion | | UKB-colocated sample + residence time criterion | | UKB-colocated sample + age match criterion | |
|---|---|---|---|---|---|---|---|---|---|
| | Correlation (95% CI) | Correlation (95% CI) | p | Correlation (95% CI) | p | Correlation (95% CI) | p | Correlation (95% CI) | p |
| **Anthropometric traits** | | | | | | | | | |
| Body mass index | 0.240 (0.233,0.246) | 0.240 (0.234,0.247) | 0.533 | 0.244 (0.236,0.253) | 0.096 | 0.239 (0.233,0.247) | 0.994 | 0.241 (0.234,0.248) | 0.127 |
| C-reactive protein (log10) | 0.132 (0.125,0.138) | 0.132 (0.125,0.139) | 0.633 | 0.132 (0.123,0.141) | 0.805 | 0.131 (0.124,0.138) | 0.645 | 0.132 (0.125,0.139) | 0.855 |
| Cholesterol | 0.079 (0.072,0.086) | 0.078 (0.071,0.085) | 0.292 | 0.082 (0.073,0.091) | 0.309 | 0.078 (0.071,0.086) | 0.406 | 0.080 (0.073,0.087) | 0.433 |
| Diastolic blood pressure | 0.102 (0.095,0.108) | 0.102 (0.095,0.109) | 0.669 | 0.107 (0.098,0.116) | 0.043 | 0.101 (0.094,0.108) | 0.558 | 0.103 (0.096,0.110) | 0.089 |
| Fluid IQ | 0.224 (0.213,0.235) | 0.225 (0.214,0.236) | 0.374 | 0.228 (0.214,0.242) | 0.355 | 0.226 (0.214,0.237) | 0.306 | 0.225 (0.214,0.236) | 0.472 |
| Forced expiratory volume in 1 second | 0.267 (0.259,0.275) | 0.269 (0.261,0.277) | 0.077 | 0.268 (0.258,0.279) | 0.752 | 0.272 (0.264,0.281) | < 8e-6 ** | 0.278 (0.269,0.286) | < 8e-6 ** |
| Forced vital capacity | 0.260 (0.251,0.268) | 0.261 (0.252,0.269) | 0.336 | 0.264 (0.254,0.275) | 0.160 | 0.263 (0.255,0.272) | 0.001 | 0.268 (0.260,0.277) | < 8e-6 ** |
| Hand grip strength | 0.179 (0.173,0.186) | 0.180 (0.173,0.186) | 0.572 | 0.184 (0.176,0.193) | 0.053 | 0.180 (0.173,0.187) | 0.357 | 0.183 (0.177,0.190) | < 8e-6 ** |
| Handedness | -0.007 (-0.028,0.013) | -0.003 (-0.023,0.017) | 0.095 | -0.006 (-0.032,0.020) | 0.871 | -0.001 (-0.022,0.020) | 0.016 | -0.005 (-0.026,0.015) | 0.320 |
| Heel bone mineral density | 0.039 (0.030,0.048) | 0.039 (0.031,0.048) | 0.703 | 0.035 (0.023,0.047) | 0.302 | 0.039 (0.029,0.048) | 0.909 | 0.039 (0.030,0.049) | 0.697 |
| Height | 0.259 (0.252,0.265) | 0.258 (0.252,0.265) | 0.914 | 0.258 (0.250,0.266) | 0.959 | 0.258 (0.252,0.265) | 0.832 | 0.263 (0.256,0.269) | < 8e-6 ** |
| Hemoglobin A1c | 0.132 (0.125,0.140) | 0.132 (0.125,0.139) | 0.440 | 0.137 (0.128,0.147) | 0.134 | 0.131 (0.124,0.139) | 0.389 | 0.135 (0.127,0.142) | 0.012 |
| High-density lipoprotein (HDL) | 0.154 (0.147,0.162) | 0.154 (0.147,0.161) | 0.872 | 0.151 (0.141,0.161) | 0.269 | 0.154 (0.146,0.162) | 0.597 | 0.155 (0.148,0.163) | 0.304 |
| Hip circumference | 0.185 (0.178,0.191) | 0.185 (0.178,0.192) | 0.564 | 0.187 (0.178,0.196) | 0.355 | 0.185 (0.178,0.192) | 0.795 | 0.186 (0.179,0.193) | 0.125 |
| Low-density lipoprotein (LDL) | 0.068 (0.061,0.075) | 0.067 (0.061,0.074) | 0.585 | 0.072 (0.063,0.081) | 0.207 | 0.067 (0.060,0.074) | 0.394 | 0.068 (0.061,0.075) | 0.623 |
| Peak expiratory flow | 0.182 (0.175,0.189) | 0.183 (0.176,0.190) | 0.335 | 0.185 (0.176,0.194) | 0.368 | 0.183 (0.176,0.190) | 0.334 | 0.187 (0.180,0.194) | < 8e-6 ** |
| Pulse | 0.112 (0.105,0.119) | 0.113 (0.106,0.120) | 0.504 | 0.115 (0.106,0.124) | 0.455 | 0.112 (0.105,0.119) | 0.841 | 0.113 (0.105,0.120) | 0.823 |
| Red blood cell count | 0.096 (0.089,0.103) | 0.096 (0.090,0.103) | 0.542 | 0.096 (0.087,0.105) | 0.979 | 0.096 (0.089,0.103) | 0.541 | 0.095 (0.088,0.102) | 0.139 |
| Systolic blood pressure | 0.161 (0.154,0.168) | 0.160 (0.154,0.167) | 0.657 | 0.167 (0.158,0.176) | 0.035 | 0.163 (0.156,0.170) | 0.054 | 0.166 (0.159,0.172) | < 8e-6 ** |
| Waist circumference | 0.261 (0.254,0.267) | 0.261 (0.255,0.268) | 0.521 | 0.264 (0.256,0.272) | 0.266 | 0.260 (0.254,0.267) | 0.742 | 0.263 (0.256,0.269) | 0.004 |
| Waist-hip ratio | 0.187 (0.181,0.193) | 0.187 (0.181,0.193) | 0.908 | 0.189 (0.181,0.197) | 0.436 | 0.186 (0.179,0.192) | 0.176 | 0.190 (0.183,0.196) | 8.8e-5 ** |
| Weight | 0.216 (0.210,0.223) | 0.216 (0.210,0.223) | 0.790 | 0.219 (0.210,0.227) | 0.353 | 0.216 (0.209,0.223) | 0.883 | 0.218 (0.211,0.225) | 0.030 |
| White blood cell count | 0.155 (0.148,0.162) | 0.155 (0.148,0.162) | 0.998 | 0.158 (0.149,0.167) | 0.271 | 0.154 (0.147,0.161) | 0.192 | 0.154 (0.147,0.161) | 0.143 |
| Whole body impedance | 0.135 (0.129,0.142) | 0.136 (0.129,0.143) | 0.432 | 0.128 (0.119,0.136) | 0.006 | 0.137 (0.130,0.144) | 0.159 | 0.137 (0.130,0.144) | 0.054 |
| **Behavioral traits** | | | | | | | | | |

| | | | | | | | | | |
|---|---|---|---|---|---|---|---|---|---|
| Age at first intercourse | 0.417 (0.410,0.424) | 0.416 (0.409,0.423) | 0.744 | 0.416 (0.407,0.425) | 0.792 | 0.427 (0.420,0.434) | < 8e-6 ** | 0.427 (0.420,0.435) | < 8e-6 ** |
| Chronotype | -0.176 (-0.188,-0.164) | -0.175 (-0.187,-0.164) | 0.793 | -0.181 (-0.196,-0.166) | 0.262 | -0.179 (-0.191,-0.167) | 0.051 | -0.180 (-0.192,-0.168) | 0.003 |
| Number of sexual partners | 0.502 (0.495,0.508) | 0.503 (0.497,0.509) | 0.073 | 0.502 (0.494,0.510) | 0.883 | 0.493 (0.486,0.499) | < 8e-6 ** | 0.498 (0.492,0.504) | < 8e-6 ** |
| Risk taking | 0.068 (0.055,0.082) | 0.068 (0.055,0.082) | 0.949 | 0.078 (0.061,0.096) | 0.071 | 0.071 (0.057,0.085) | 0.234 | 0.066 (0.052,0.080) | 0.189 |
| Sleep duration | 0.120 (0.113,0.127) | 0.119 (0.112,0.126) | 0.380 | 0.129 (0.120,0.138) | 0.002 | 0.122 (0.115,0.129) | 0.018 | 0.120 (0.113,0.128) | 0.309 |
| Time spent using computer | 0.101 (0.094,0.107) | 0.101 (0.095,0.108) | 0.467 | 0.097 (0.089,0.106) | 0.220 | 0.098 (0.091,0.105) | 0.003 | 0.101 (0.094,0.107) | 0.765 |
| Time spent watching TV | 0.446 (0.441,0.452) | 0.446 (0.441,0.452) | 0.862 | 0.468 (0.461,0.476) | < 8e-6 ** | 0.448 (0.442,0.454) | 0.046 | 0.447 (0.442,0.453) | 0.114 |
| Water intake | 0.214 (0.208,0.221) | 0.213 (0.207,0.219) | 0.212 | 0.222 (0.213,0.230) | 0.005 | 0.216 (0.209,0.223) | 0.032 | 0.214 (0.207,0.220) | 0.644 |
| **Demographic/Family traits** | | | | | | | | | |
| Age | 0.855 (0.852,0.857) | 0.853 (0.851,0.856) | 4.2e-4 | 0.857 (0.854,0.860) | 0.005 | 0.866 (0.864,0.868) | < 8e-6 ** | 0.911 (0.910,0.912) | < 8e-6 ** |
| Birthplace (east coordinate) | 0.497 (0.490,0.505) | 0.499 (0.491,0.506) | 0.243 | 0.478 (0.468,0.488) | < 8e-6 ** | 0.500 (0.492,0.508) | 0.014 | 0.502 (0.494,0.510) | < 8e-6 ** |
| Birthplace (north coordinate) | 0.650 (0.644,0.656) | 0.652 (0.646,0.658) | 0.023 | 0.630 (0.622,0.638) | < 8e-6 ** | 0.651 (0.644,0.657) | 0.454 | 0.654 (0.648,0.660) | < 8e-6 ** |
| Breastfed | 0.291 (0.276,0.305) | 0.292 (0.278,0.306) | 0.428 | 0.290 (0.272,0.308) | 0.881 | 0.288 (0.273,0.303) | 0.227 | 0.295 (0.280,0.309) | 0.017 |
| Educational attainment | 0.492 (0.487,0.498) | 0.492 (0.487,0.498) | 0.784 | 0.504 (0.497,0.511) | 8.0e-6 ** | 0.496 (0.490,0.501) | < 8e-6 ** | 0.498 (0.492,0.503) | < 8e-6 ** |
| Mother smoked during pregnancy | 0.147 (0.134,0.160) | 0.147 (0.135,0.160) | 0.972 | 0.143 (0.126,0.159) | 0.366 | 0.148 (0.134,0.161) | 0.864 | 0.151 (0.137,0.164) | 0.025 |
| Number of children | 0.721 (0.715,0.726) | 0.717 (0.712,0.722) | < 8e-6 ** | 0.719 (0.712,0.726) | 0.537 | 0.752 (0.747,0.757) | < 8e-6 ** | 0.751 (0.745,0.756) | < 8e-6 ** |
| Number of siblings | 0.099 (0.092,0.105) | 0.099 (0.093,0.106) | 0.422 | 0.091 (0.083,0.100) | 0.009 | 0.101 (0.095,0.108) | 0.006 | 0.100 (0.093,0.106) | 0.244 |
| **Health behaviors** | | | | | | | | | |
| Drinks alcohol | 0.572 (0.556,0.588) | 0.572 (0.557,0.588) | 0.849 | 0.588 (0.566,0.608) | 0.028 | 0.582 (0.565,0.598) | < 8e-6 ** | 0.568 (0.551,0.584) | 0.031 |
| Drinks full-fat or semi-skimmed milk | 0.748 (0.742,0.754) | 0.747 (0.741,0.754) | 0.473 | 0.764 (0.756,0.772) | < 8e-6 ** | 0.749 (0.743,0.756) | 0.198 | 0.749 (0.742,0.755) | 0.482 |
| Eats processed meat >= 1x week | 0.423 (0.414,0.433) | 0.420 (0.411,0.430) | 0.021 | 0.446 (0.433,0.458) | < 8e-6 ** | 0.426 (0.416,0.436) | 0.089 | 0.424 (0.414,0.434) | 0.381 |
| Ever smoked | 0.243 (0.233,0.254) | 0.242 (0.232,0.252) | 0.167 | 0.244 (0.231,0.257) | 0.969 | 0.249 (0.238,0.259) | 5.2e-4 | 0.245 (0.235,0.256) | 0.134 |
| Fruit intake | 0.252 (0.245,0.260) | 0.253 (0.245,0.260) | 0.915 | 0.259 (0.249,0.268) | 0.058 | 0.254 (0.246,0.262) | 0.203 | 0.252 (0.245,0.260) | 0.688 |
| Moderate physical activity (days/week) | 0.115 (0.108,0.122) | 0.116 (0.109,0.123) | 0.739 | 0.131 (0.122,0.139) | < 8e-6 ** | 0.117 (0.110,0.125) | 0.042 | 0.116 (0.109,0.123) | 0.619 |
| Smoked at least 100 times | 0.367 (0.357,0.376) | 0.364 (0.355,0.374) | 0.064 | 0.372 (0.359,0.384) | 0.202 | 0.373 (0.362,0.383) | 1.2e-4 ** | 0.371 (0.361,0.380) | 3.8e-4 |
| Smoking status (Current) | 0.511 (0.495,0.526) | 0.514 (0.499,0.530) | 0.104 | 0.532 (0.511,0.552) | 0.002 | 0.505 (0.488,0.522) | 0.074 | 0.505 (0.488,0.521) | 0.009 |
| Smoking status (Former) | 0.314 (0.303,0.324) | 0.311 (0.301,0.321) | 0.109 | 0.322 (0.309,0.335) | 0.048 | 0.321 (0.311,0.331) | < 8e-6 ** | 0.319 (0.309,0.329) | < 8e-6 ** |
| Smoking status (Never) | 0.368 (0.358,0.377) | 0.366 (0.357,0.375) | 0.130 | 0.372 (0.360,0.384) | 0.303 | 0.374 (0.364,0.384) | 2.4e-5 ** | 0.372 (0.362,0.382) | 4.0e-5 ** |
| Sun protection | 0.243 (0.233,0.254) | 0.242 (0.232,0.253) | 0.439 | 0.237 (0.223,0.250) | 0.132 | 0.246 (0.235,0.257) | 0.081 | 0.242 (0.231,0.253) | 0.224 |
| Vegetable intake | 0.257 (0.250,0.264) | 0.256 (0.249,0.263) | 0.724 | 0.272 (0.263,0.282) | < 8e-6 ** | 0.258 (0.251,0.265) | 0.275 | 0.257 (0.250,0.264) | 0.884 |

| | | | | | | | | |
|---|---|---|---|---|---|---|---|---|
| Walking (minutes/day) | 0.188 (0.180,0.196) | 0.186 (0.178,0.194) | 0.046 | 0.193 (0.183,0.202) | 0.154 | 0.188 (0.180,0.196) | 0.999 | 0.188 (0.180,0.196) | 0.996 |
| **Health traits** | | | | | | | | | |
| Asthma | -0.004 (-0.022,0.015) | 0.003 (-0.015,0.021) | 0.008 | 0.006 (-0.018,0.029) | 0.202 | -0.009 (-0.029,0.010) | 0.054 | -0.005 (-0.024,0.014) | 0.685 |
| Basal metabolic rate | 0.206 (0.199,0.213) | 0.206 (0.200,0.213) | 0.872 | 0.204 (0.196,0.213) | 0.511 | 0.207 (0.200,0.214) | 0.446 | 0.209 (0.203,0.216) | 4.0e-5 ** |
| Dental problems | 0.145 (0.135,0.156) | 0.145 (0.135,0.155) | 0.965 | 0.146 (0.132,0.159) | 0.941 | 0.144 (0.133,0.155) | 0.378 | 0.147 (0.136,0.157) | 0.248 |
| Diabetes | 0.199 (0.173,0.225) | 0.196 (0.170,0.221) | 0.282 | 0.209 (0.174,0.243) | 0.378 | 0.201 (0.174,0.228) | 0.590 | 0.200 (0.172,0.226) | 0.867 |
| Difficulty hearing | -0.069 (-0.081,-0.056) | -0.070 (-0.082,-0.058) | 0.328 | -0.068 (-0.084,-0.052) | 0.921 | -0.070 (-0.083,-0.057) | 0.454 | -0.068 (-0.081,-0.055) | 0.529 |
| Ever had tinnitus | 0.065 (0.045,0.085) | 0.065 (0.046,0.085) | 0.821 | 0.067 (0.042,0.092) | 0.792 | 0.069 (0.049,0.090) | 0.123 | 0.066 (0.046,0.086) | 0.574 |
| Hay fever, allergic rhinitis, or eczema | 0.015 (0.002,0.027) | 0.011 (-0.001,0.024) | 0.045 | 0.009 (-0.008,0.025) | 0.238 | 0.013 (-0.001,0.026) | 0.270 | 0.016 (0.003,0.029) | 0.208 |
| High blood pressure | 0.099 (0.087,0.111) | 0.097 (0.085,0.108) | 0.102 | 0.107 (0.092,0.122) | 0.106 | 0.099 (0.087,0.111) | 0.888 | 0.102 (0.090,0.114) | 0.010 |
| Long-term illness | 0.135 (0.123,0.146) | 0.136 (0.125,0.147) | 0.377 | 0.132 (0.117,0.146) | 0.530 | 0.134 (0.122,0.145) | 0.482 | 0.134 (0.122,0.145) | 0.377 |
| Number of illnesses | 0.160 (0.153,0.166) | 0.159 (0.153,0.166) | 0.635 | 0.157 (0.148,0.165) | 0.248 | 0.160 (0.154,0.167) | 0.494 | 0.163 (0.156,0.170) | < 8e-6 ** |
| Number of medications | 0.218 (0.212,0.224) | 0.219 (0.213,0.225) | 0.180 | 0.217 (0.209,0.225) | 0.729 | 0.218 (0.212,0.225) | 0.844 | 0.222 (0.215,0.228) | < 8e-6 ** |
| Wears Glasses | 0.381 (0.364,0.398) | 0.381 (0.364,0.397) | 0.961 | 0.389 (0.368,0.410) | 0.221 | 0.389 (0.372,0.406) | 2.6e-4 | 0.401 (0.384,0.418) | < 8e-6 ** |
| **Psychological traits** | | | | | | | | | |
| Anxious feelings | -0.101 (-0.112,-0.090) | -0.099 (-0.109,-0.088) | 0.125 | -0.100 (-0.114,-0.086) | 0.872 | -0.104 (-0.116,-0.093) | 0.026 | -0.103 (-0.114,-0.092) | 0.064 |
| Doctor-diagnosed anxiety or depression | 0.152 (0.140,0.163) | 0.152 (0.140,0.163) | 0.997 | 0.158 (0.143,0.172) | 0.182 | 0.152 (0.140,0.164) | 0.790 | 0.151 (0.139,0.162) | 0.632 |
| Ever depressed for at least one week | 0.141 (0.123,0.158) | 0.141 (0.123,0.159) | 0.843 | 0.138 (0.115,0.160) | 0.698 | 0.138 (0.119,0.156) | 0.237 | 0.138 (0.119,0.156) | 0.131 |
| Fed-up feelings | 0.151 (0.140,0.162) | 0.150 (0.140,0.161) | 0.699 | 0.154 (0.141,0.168) | 0.417 | 0.151 (0.139,0.161) | 0.609 | 0.153 (0.142,0.164) | 0.028 |
| Guilt | 0.061 (0.049,0.074) | 0.059 (0.047,0.072) | 0.244 | 0.062 (0.046,0.078) | 0.954 | 0.061 (0.048,0.074) | 0.988 | 0.063 (0.050,0.076) | 0.287 |
| High strung | 0.022 (0.007,0.038) | 0.022 (0.007,0.037) | 0.924 | 0.025 (0.005,0.045) | 0.683 | 0.022 (0.007,0.038) | 0.990 | 0.023 (0.008,0.039) | 0.532 |
| Hurt feelings | 0.043 (0.032,0.054) | 0.045 (0.034,0.056) | 0.159 | 0.038 (0.023,0.052) | 0.246 | 0.043 (0.032,0.055) | 0.761 | 0.043 (0.032,0.054) | 0.918 |
| Irritability | -0.019 (-0.031,-0.007) | -0.019 (-0.031,-0.007) | 0.943 | -0.016 (-0.032,-0.001) | 0.560 | -0.019 (-0.031,-0.006) | 0.855 | -0.019 (-0.032,-0.007) | 0.792 |
| Loneliness | 0.157 (0.140,0.174) | 0.160 (0.143,0.177) | 0.197 | 0.157 (0.134,0.179) | 0.994 | 0.160 (0.142,0.178) | 0.275 | 0.157 (0.139,0.174) | 0.807 |
| Miserableness | 0.109 (0.098,0.120) | 0.108 (0.098,0.119) | 0.502 | 0.108 (0.094,0.122) | 0.782 | 0.107 (0.095,0.118) | 0.113 | 0.108 (0.097,0.119) | 0.513 |
| Mood swings | 0.059 (0.048,0.070) | 0.058 (0.048,0.069) | 0.676 | 0.055 (0.042,0.069) | 0.414 | 0.055 (0.044,0.066) | 0.016 | 0.058 (0.047,0.069) | 0.480 |
| Nerves | 0.079 (0.065,0.092) | 0.077 (0.063,0.090) | 0.218 | 0.078 (0.060,0.095) | 0.843 | 0.076 (0.062,0.090) | 0.173 | 0.076 (0.062,0.090) | 0.072 |
| Nervous feelings | 0.036 (0.023,0.049) | 0.033 (0.020,0.045) | 0.068 | 0.037 (0.020,0.054) | 0.777 | 0.037 (0.023,0.050) | 0.523 | 0.033 (0.020,0.046) | 0.068 |
| Neuroticism | 0.042 (0.033,0.050) | 0.042 (0.034,0.050) | 0.701 | 0.045 (0.035,0.055) | 0.316 | 0.038 (0.030,0.047) | 0.003 | 0.042 (0.034,0.050) | 0.572 |
| Worry too long after embarrassment | 0.021 (0.010,0.032) | 0.022 (0.011,0.032) | 0.810 | 0.024 (0.010,0.038) | 0.575 | 0.021 (0.009,0.032) | 0.708 | 0.021 (0.010,0.033) | 0.930 |

### Supplemental Table 19: Trait missing value counts for the UKB-colocated pairs

| | Neither missing data | One partner missing data | | | | Both missing data |
|---|---|---|---|---|---|---|
| | | Female missing | Male missing | Total | Proportion | |
| **Anthropometric traits** | | | | | | |
| Body mass index | 88690 | 232 | 274 | 506 | 0.006 | 82 |
| C-reactive protein (log10) | 79388 | 4702 | 4019 | 8721 | 0.098 | 1169 |
| Cholesterol | 79705 | 4596 | 3859 | 8455 | 0.095 | 1118 |
| Diastolic blood pressure | 82558 | 2226 | 2183 | 4409 | 0.049 | 2311 |
| Fluid IQ | 29150 | 851 | 826 | 1677 | 0.019 | 58451 |
| Forced expiratory volume in 1 second | 50455 | 16631 | 14220 | 30851 | 0.346 | 7972 |
| Forced vital capacity | 50455 | 16631 | 14220 | 30851 | 0.346 | 7972 |
| Hand grip strength | 88855 | 162 | 164 | 326 | 0.004 | 97 |
| Handedness | 86432 | 1043 | 1769 | 2812 | 0.031 | 34 |
| Heel bone mineral density | 48295 | 2265 | 2276 | 4541 | 0.051 | 36442 |
| Height | 88828 | 158 | 211 | 369 | 0.004 | 81 |
| Hemoglobin A1c | 79980 | 3972 | 3385 | 7357 | 0.082 | 1941 |
| High-density lipoprotein (HDL) | 68182 | 9615 | 7757 | 17372 | 0.195 | 3724 |
| Hip circumference | 88891 | 151 | 157 | 308 | 0.003 | 79 |
| Low-density lipoprotein (LDL) | 79464 | 4672 | 3959 | 8631 | 0.097 | 1183 |
| Peak expiratory flow | 74820 | 5958 | 7110 | 13068 | 0.146 | 1390 |
| Pulse | 82558 | 2226 | 2183 | 4409 | 0.049 | 2311 |
| Red blood cell count | 82911 | 3170 | 2314 | 5484 | 0.061 | 883 |
| Systolic blood pressure | 82554 | 2229 | 2184 | 4413 | 0.049 | 2311 |
| Waist circumference | 88911 | 146 | 142 | 288 | 0.003 | 79 |
| Waist-hip ratio | 88876 | 160 | 163 | 323 | 0.004 | 79 |
| Weight | 88771 | 209 | 218 | 427 | 0.005 | 80 |
| White blood cell count | 82910 | 3170 | 2315 | 5485 | 0.061 | 883 |
| Whole body impedance | 86414 | 1255 | 1399 | 2654 | 0.030 | 210 |
| **Behavioral traits** | | | | | | |
| Age at first intercourse | 71629 | 8043 | 6614 | 14657 | 0.164 | 2992 |
| Chronotype | 71187 | 6174 | 10107 | 16281 | 0.182 | 1810 |
| Number of sexual partners | 64840 | 10074 | 10360 | 20434 | 0.229 | 4004 |
| Risk taking | 83873 | 2838 | 2394 | 5232 | 0.059 | 173 |
| Sleep duration | 88539 | 492 | 230 | 722 | 0.008 | 17 |
| Time spent using computer | 87778 | 690 | 477 | 1167 | 0.013 | 333 |
| Time spent watching TV | 88053 | 811 | 378 | 1189 | 0.013 | 36 |
| Water intake | 88221 | 526 | 516 | 1042 | 0.012 | 15 |
| **Demographic/Family traits** | | | | | | |
| Age | 89278 | 0 | 0 | 0 | 0.000 | 0 |
| Birthplace (east coordinate) | 77487 | 5158 | 4027 | 9185 | 0.103 | 2606 |
| Birthplace (north coordinate) | 77487 | 5158 | 4027 | 9185 | 0.103 | 2606 |
| Breastfed | 53054 | 9836 | 18325 | 28161 | 0.315 | 8063 |
| Educational attainment | 87175 | 883 | 873 | 1756 | 0.020 | 347 |

| | | | | | | |
|---|---|---|---|---|---|---|
| Mother smoked during pregnancy | 66895 | 9820 | 10579 | 20399 | 0.228 | 1984 |
| Number of children | 88554 | 59 | 645 | 704 | 0.008 | 20 |
| Number of siblings | 89278 | 0 | 0 | 0 | 0.000 | 0 |
| **Health behaviors** | | | | | | |
| Drinks alcohol | 89156 | 80 | 39 | 119 | 0.001 | 3 |
| Drinks full-fat or semi-skimmed milk | 89188 | 32 | 52 | 84 | 0.001 | 6 |
| Eats processed meat >= 1x week | 89033 | 138 | 100 | 238 | 0.003 | 7 |
| Ever smoked | 88656 | 303 | 306 | 609 | 0.007 | 13 |
| Fruit intake | 74251 | 6796 | 7196 | 13992 | 0.157 | 1035 |
| Moderate physical activity (days/week) | 80788 | 5285 | 2672 | 7957 | 0.089 | 533 |
| Smoked at least 100 times | 86610 | 1563 | 1039 | 2602 | 0.029 | 66 |
| Smoking status (Current) | 88634 | 311 | 320 | 631 | 0.007 | 13 |
| Smoking status (Former) | 88634 | 311 | 320 | 631 | 0.007 | 13 |
| Smoking status (Never) | 88634 | 311 | 320 | 631 | 0.007 | 13 |
| Sun protection | 88181 | 475 | 296 | 771 | 0.009 | 326 |
| Vegetable intake | 86536 | 1094 | 1511 | 2605 | 0.029 | 137 |
| Walking (minutes/day) | 65831 | 12590 | 7610 | 20200 | 0.226 | 3247 |
| **Health traits** | | | | | | |
| Asthma | 89051 | 117 | 90 | 207 | 0.002 | 20 |
| Basal metabolic rate | 86426 | 1242 | 1400 | 2642 | 0.030 | 210 |
| Dental problems | 88574 | 233 | 166 | 399 | 0.004 | 305 |
| Diabetes | 88892 | 181 | 187 | 368 | 0.004 | 18 |
| Difficulty hearing | 82514 | 3660 | 2636 | 6296 | 0.071 | 468 |
| Ever had tinnitus | 29722 | 651 | 667 | 1318 | 0.015 | 58238 |
| Hay fever, allergic rhinitis, or eczema | 89051 | 117 | 90 | 207 | 0.002 | 20 |
| High blood pressure | 88972 | 177 | 113 | 290 | 0.003 | 16 |
| Long-term illness | 85600 | 2058 | 1561 | 3619 | 0.041 | 59 |
| Number of illnesses | 89202 | 24 | 27 | 51 | 0.001 | 25 |
| Number of medications | 89202 | 24 | 27 | 51 | 0.001 | 25 |
| Wears Glasses | 89129 | 70 | 68 | 138 | 0.002 | 11 |
| **Psychological traits** | | | | | | |
| Anxious feelings | 85034 | 1955 | 2198 | 4153 | 0.047 | 91 |
| Doctor-diagnosed anxiety or depression | 88188 | 675 | 394 | 1069 | 0.012 | 21 |
| Ever depressed for at least one week | 29564 | 736 | 751 | 1487 | 0.017 | 58227 |
| Fed-up feelings | 85660 | 1877 | 1657 | 3534 | 0.040 | 84 |
| Guilt | 84899 | 2402 | 1866 | 4268 | 0.048 | 111 |
| High strung | 83987 | 3046 | 2059 | 5105 | 0.057 | 186 |
| Hurt feelings | 84369 | 2327 | 2468 | 4795 | 0.054 | 114 |
| Irritability | 81555 | 4032 | 3387 | 7419 | 0.083 | 304 |
| Loneliness | 86964 | 1325 | 944 | 2269 | 0.025 | 45 |
| Miserableness | 86237 | 1535 | 1430 | 2965 | 0.033 | 76 |
| Mood swings | 84983 | 2114 | 2056 | 4170 | 0.047 | 125 |
| Nerves | 83239 | 3539 | 2337 | 5876 | 0.066 | 163 |
| Nervous feelings | 84952 | 2713 | 1534 | 4247 | 0.048 | 79 |

| | | | | | | |
|---|---|---|---|---|---|---|
| Neuroticism | 59442 | 14672 | 11366 | 26038 | 0.292 | 3798 |
| Worry too long after embarrassment | 82222 | 3796 | 3039 | 6835 | 0.077 | 221 |

**Supplemental Table 20: Comparison of trait means for people whose partners are or are not missing data for that trait**

| | Females | | | Males | | |
|---|---|---|---|---|---|---|
| | Partner missing | Partner not missing | P | Partner missing | Partner not missing | P |
| **Anthropometric traits** | | | | | | |
| C-reactive protein (log10(mg/L)) | 0.162 (0.007) | 0.154 (0.002) | 0.258 | 0.129 (0.006) | 0.113 (0.001) | 0.015 |
| Cholesterol (mmol/L) | 5.895 (0.017) | 5.928 (0.004) | 0.060 | 5.483 (0.016) | 5.475 (0.004) | 0.616 |
| Diastolic blood pressure (mmHg) | 81.338 (0.200) | 81.083 (0.034) | 0.209 | 84.710 (0.192) | 84.003 (0.034) | 2.94e-04 ** |
| Fluid IQ (score) | 5.259 (0.071) | 5.950 (0.012) | 6.44e-21 ** | 5.535 (0.077) | 6.288 (0.013) | 6.16e-21 ** |
| Forced expiratory volume in 1 second (L) | 2.387 (0.004) | 2.428 (0.002) | 5.46e-19 ** | 3.278 (0.005) | 3.324 (0.003) | 9.46e-14 ** |
| Forced vital capacity (L) | 3.142 (0.005) | 3.180 (0.003) | 6.14e-12 ** | 4.370 (0.006) | 4.427 (0.004) | 6.44e-15 ** |
| Handedness (proportion left-handed) | 0.083 (0.007) | 0.086 (0.001) | 0.554 | 0.080 (0.008) | 0.104 (0.001) | 0.004 |
| Heel bone mineral density (standard deviations) | -0.579 (0.018) | -0.589 (0.005) | 0.572 | -0.095 (0.022) | -0.111 (0.005) | 0.473 |
| Hemoglobin A1c (mmol/mol) | 35.383 (0.059) | 35.215 (0.014) | 0.005 | 35.917 (0.070) | 35.660 (0.017) | 3.26e-04 ** |
| High-density lipoprotein (HDL) (mmol/L) | 1.607 (0.004) | 1.594 (0.001) | 0.002 | 1.275 (0.003) | 1.271 (0.001) | 0.274 |
| Low-density lipoprotein (LDL) (mmol/L) | 3.634 (0.013) | 3.659 (0.003) | 0.058 | 3.474 (0.012) | 3.472 (0.003) | 0.828 |
| Peak expiratory flow (L/min) | 300.989 (1.000) | 309.011 (0.305) | 1.87e-14 ** | 433.726 (1.580) | 447.215 (0.451) | 2.65e-16 ** |
| Pulse (bpm) | 70.764 (0.196) | 69.540 (0.035) | 8.99e-10 ** | 67.958 (0.206) | 66.776 (0.038) | 1.99e-08 ** |
| Red blood cell count (10^12 cells/L) | 4.342 (0.006) | 4.335 (0.001) | 0.239 | 4.750 (0.006) | 4.740 (0.001) | 0.113 |
| Systolic blood pressure (mmHg) | 137.814 (0.374) | 136.793 (0.065) | 0.007 | 143.681 (0.338) | 141.678 (0.059) | 5.69e-09 ** |
| White blood cell count (10^9 cells/L) | 6.783 (0.032) | 6.770 (0.006) | 0.704 | 6.912 (0.028) | 6.847 (0.006) | 0.021 |
| Whole body impedance (ohms) | 656.084 (1.705) | 656.822 (0.241) | 0.668 | 533.699 (1.446) | 533.732 (0.186) | 0.982 |
| **Behavioral traits** | | | | | | |
| Age at first intercourse | 19.433 (0.037) | 18.976 (0.011) | 2.13e-32 ** | 19.610 (0.042) | 19.151 (0.013) | 5.92e-26 ** |
| Chronotype (proportion morning) | 0.329 (0.005) | 0.359 (0.002) | 2.49e-09 ** | 0.333 (0.006) | 0.362 (0.002) | 2.35e-06 ** |
| Number of sexual partners | 3.099 (0.034) | 3.138 (0.014) | 0.302 | 5.913 (0.078) | 5.080 (0.027) | 9.99e-24 ** |
| Risk taking (proportion) | 0.155 (0.007) | 0.139 (0.001) | 0.036 | 0.346 (0.009) | 0.316 (0.002) | 9.03e-04 |
| Time spent using computer (hours/day) | 0.817 (0.048) | 0.691 (0.004) | 0.008 | 1.105 (0.047) | 1.146 (0.005) | 0.388 |
| Time spent watching TV (hours/day) | 3.652 (0.074) | 2.824 (0.005) | 1.92e-26 ** | 3.489 (0.054) | 2.840 (0.005) | 3.15e-31 ** |
| Water intake (glasses/day) | 2.926 (0.081) | 2.758 (0.007) | 0.041 | 2.261 (0.078) | 2.156 (0.006) | 0.179 |
| **Demographic/Family traits** | | | | | | |
| Birthplace (east coordinate) | 418712.036 (1279.588) | 413379.880 (269.526) | 4.63e-05 ** | 425638.403 (1181.171) | 414122.805 (270.039) | 2.86e-21 ** |
| Birthplace (north coordinate) | 347338.192 (2570.829) | 361233.949 (557.224) | 1.33e-07 ** | 327622.060 (2284.715) | 359054.615 (556.747) | 3.56e-40 ** |
| Breastfed (proportion) | 0.675 (0.003) | 0.709 (0.002) | 6.17e-18 ** | 0.774 (0.004) | 0.774 (0.002) | 0.982 |
| Educational attainment (estimated years) | 11.270 (0.142) | 13.458 (0.016) | 3.12e-47 ** | 12.078 (0.155) | 14.193 (0.017) | 3.71e-38 ** |
| Mother smoked during pregnancy (proportion) | 0.300 (0.004) | 0.286 (0.002) | 0.005 | 0.325 (0.005) | 0.305 (0.002) | 8.77e-05 ** |
| **Health behaviors** | | | | | | |
| Fruit intake (pieces/day) | 3.301 (0.024) | 3.250 (0.008) | 0.040 | 2.574 (0.022) | 2.673 (0.007) | 2.66e-05 ** |
| Moderate physical activity (days/week) | 3.534 (0.046) | 3.611 (0.008) | 0.101 | 3.552 (0.032) | 3.621 (0.008) | 0.037 |
| Smoked at least 100 times (proportion) | 0.344 (0.015) | 0.349 (0.002) | 0.717 | 0.569 (0.013) | 0.520 (0.002) | 1.21e-04 ** |
| Vegetable intake (Tbsp/day) | 5.164 (0.053) | 4.932 (0.008) | 1.38e-05 ** | 5.000 (0.066) | 4.535 (0.008) | 2.56e-12 ** |
| Walking (minutes/day) | 57.017 (0.784) | 57.163 (0.250) | 0.859 | 64.513 (0.701) | 61.270 (0.281) | 1.77e-05 ** |

| | | | | | | |
|---|---|---|---|---|---|---|
| **Health traits** | | | | | | |
| Basal metabolic rate (kJ) | 5723.170 (14.545) | 5585.400 (2.030) | 1.61e-20 ** | 7941.625 (22.378) | 7736.630 (3.230) | 2.58e-19 ** |
| Difficulty hearing (proportion) | 0.200 (0.008) | 0.211 (0.001) | 0.159 | 0.330 (0.008) | 0.341 (0.002) | 0.153 |
| Ever had tinnitus (proportion) | 0.276 (0.017) | 0.260 (0.003) | 0.372 | 0.293 (0.018) | 0.323 (0.003) | 0.106 |
| Long-term illness (proportion) | 0.307 (0.012) | 0.283 (0.002) | 0.035 | 0.375 (0.011) | 0.344 (0.002) | 0.005 |
| **Psychological traits** | | | | | | |
| Anxious feelings (proportion) | 0.671 (0.010) | 0.672 (0.002) | 0.867 | 0.483 (0.011) | 0.467 (0.002) | 0.153 |
| Doctor-diagnosed anxiety or depression (proportion) | 0.411 (0.025) | 0.378 (0.002) | 0.179 | 0.230 (0.016) | 0.234 (0.001) | 0.770 |
| Ever depressed for at least one week (proportion) | 0.533 (0.018) | 0.539 (0.003) | 0.730 | 0.398 (0.018) | 0.413 (0.003) | 0.411 |
| Fed-up feelings (proportion) | 0.428 (0.012) | 0.412 (0.002) | 0.184 | 0.373 (0.011) | 0.339 (0.002) | 0.003 |
| Guilt (proportion) | 0.387 (0.011) | 0.339 (0.002) | 2.45e-05 ** | 0.185 (0.008) | 0.191 (0.001) | 0.430 |
| High strung (proportion) | 0.231 (0.009) | 0.198 (0.001) | 3.47e-04 ** | 0.148 (0.006) | 0.133 (0.001) | 0.020 |
| Hurt feelings (proportion) | 0.659 (0.010) | 0.663 (0.002) | 0.698 | 0.430 (0.010) | 0.438 (0.002) | 0.454 |
| Irritability (proportion) | 0.248 (0.007) | 0.269 (0.002) | 0.006 | 0.312 (0.007) | 0.308 (0.002) | 0.622 |
| Loneliness (proportion) | 0.199 (0.013) | 0.146 (0.001) | 5.76e-05 ** | 0.098 (0.008) | 0.083 (0.001) | 0.075 |
| Miserableness (proportion) | 0.499 (0.013) | 0.488 (0.002) | 0.406 | 0.324 (0.012) | 0.328 (0.002) | 0.763 |
| Mood swings (proportion) | 0.490 (0.011) | 0.472 (0.002) | 0.109 | 0.416 (0.011) | 0.403 (0.002) | 0.228 |
| Nerves (proportion) | 0.227 (0.009) | 0.214 (0.001) | 0.152 | 0.204 (0.007) | 0.191 (0.001) | 0.074 |
| Nervous feelings (proportion) | 0.300 (0.012) | 0.299 (0.002) | 0.968 | 0.173 (0.007) | 0.172 (0.001) | 0.898 |
| Neuroticism (score) | 4.650 (0.030) | 4.602 (0.013) | 0.150 | 3.379 (0.025) | 3.316 (0.012) | 0.025 |
| Worry too long after embarrassment (proportion) | 0.575 (0.009) | 0.564 (0.002) | 0.232 | 0.380 (0.008) | 0.383 (0.002) | 0.638 |
